# Effects of Exercise Combined With Green Tea in Non-Aging on Anthropometric and Blood Lipids in Non-Aging Overweight/Obesity Adults-A Systematic Review and Network Meta-Analysis

**DOI:** 10.1101/2025.09.07.25334837

**Authors:** Chuang Liu, Yu Wang, Yuxin Shen, Yang Gao

## Abstract

Increasing physical activity and drinking green tea are alternate non-surgical or pharmaceutical complementary treatments that many individuals opt to employ to manage overweight/obesity, and a number of recent clinical randomized controlled studies (RCTs) have shown interest in combining the two. The effectiveness of these three therapies in treating overweight/obesity has not been summarized, compared, or interpreted. A reticulated meta-analysis is superior than a meta-analysis for comparing and rating the effectiveness of different therapies for a particular condition. Through a reticulated meta-analysis, we wanted to examine the efficacy of three alternative supplementary treatments for overweight/obesity, namely green tea, physical activity, and green tea coupled with physical activity, and to provide recommendations. A total of 44 RCTs assessing the effects of nine alternative supplementary treatments on body weight, BMI, body fat percentage, waist circumference, total cholesterol, triglycerides, low-density lipoprotein, and high-density lipoprotein were included. Green tea combined with aerobic exercise may be optimal for reducing body weight, BMI, TC, and LDL; green tea combined with resistance training may be optimal for reducing body fat percentage and waist circumference; aerobic exercise may be optimal for reducing TG; and resistance combined with aerobic exercise may be optimal for increasing HDL. Nevertheless, based on the NMA results for the aforementioned eight indicators, we believe that the combination of green tea and increased physical activity is preferable to green tea or increased physical activity alone for non-elderly adult populations seeking non-surgical or pharmacological treatment or management of overweight/obesity.

## 1. Introduction

According to the World Health Organization (WHO), overweight/obesity is a non-communicable chronic disease that affects more than 1.6 billion people worldwide[1]. Despite repeated calls from relevant health organizations to pay greater attention to overweight/obesity, this number continues to rise as society becomes more convenient and materially wealthy[2]. Overweight/obesity serves as a “breeding environment” for several ailments, including diabetes, fatty liver, and atherosclerosis[3].

In the clinical management of overweight/obesity, clinicians use a variety of measures, including medication, surgery, and energy-restricted diets[4,5], but the efficacy and safety of medication (e.g. sibutramine or orlistat) and surgery (e.g. liposuction) are still highly variable due to the different body types of patients and the presence of potential risk factors[6,7]. The good news is that individuals are experimenting with non-pharmaceutical and non-surgical complementary treatments to prevent and cure overweight/obesity, such as nutritional supplements, energy-restricted diets, and increased physical activity.

The core of exercise as a prescription therapy for overweight/obesity is increasing physical activity, boosting calorie intake, and mobilizing more adipose tissue to participate in the functioning of the energy system in order to burn excess fat[8].

In addition, a globally popular beverage, tea, may be a potentially beneficial dietary supplement for the treatment of overweight/obesity. As an unfermented Camellia sinensis product, green tea is rich in catechins that have significant health benefits, particularly epigallocatechin gallate EGCG, the main component of catechins (catechins make up 30-42% of the dried tea leaves, and EGCG accounts for 60-65% of the catechin content and is therefore regarded as the substance with the most significant health role)[9]. Yang has established that green tea and its catechins control hunger and limit energy absorption in his study[10], Boschmann’s research has demonstrated that green tea regulates fat metabolism and thermogenesis[11], and Chatree’s research has demonstrated that green tea inhibits fat formation and accumulation[12]. Therefore, physicians and researchers are increasingly using green tea as an effective and easily available dietary supplement for the treatment of overweight/obesity, and this is becoming a trend[13,14].

The use of green tea or increased physical activity as a supplementary treatment for overweight/obesity has been acknowledged for a long time. Additionally, a portion of recent research has begun to focus on combining the two for the treatment of overweight/obesity, with multiple animal models demonstrating that exercise combined with green tea has a positive effect on overweight/obesity control in high-fat-fed rats and is more effective than exercise or green tea alone[15,16]. This seems to indicate a novel supplemental alternative medicine for the treatment of overweight/obesity.

Numerous randomized controlled trials (RCTs) have investigated the efficacy of the three modalities described above (exercise, green tea, and green tea during exercise) as an alternative complementary treatment for overweight/obesity; however, the results of individual RCTs on these interventions for overweight/obesity are not uniform and there are differences in evidence-based meta-analyses, indicating that these interventions may have different effects on the treatment of overweight/obesity. To our knowledge, there is no research compared the efficacy and consequences of these therapies for overweight/obesity. We feel it is necessary to do a pooled analysis to assess and identify the best therapies using a net meta-analysis in order to give physicians and the obese population with recommendations.

## 2. Materials and Methods

### 2.1. Search Strategy

We consequently checked five internet databases, including Web of Science, Pubmed, Cochrane, Sport Discus, and EMBASE. The search term is roughly “green tea OR tea OR green tea extract OR EGCG OR epigallocatechin gallate OR catechins AND exercise OR physical activity OR exercise, physical OR isometric exercise OR aerobic exercise OR exercise training OR training OR resistance training AND overweight/obesity OR obese OR overweight”. Detailed search strategies can be found in the **Supplementary Table 1**.

### 2.2. Eligibility criteria for inclusion

According to PICOS principles, the original research were evaluated and incorporated[17]. (P) individuals (>18 years) having a BMI more than or equal to 25 kg/m2, excluding secondary overweight/obesity or overweight/obesity in conjunction with other conditions; (I) treatments of increased exercise or green tea or increased exercise in conjunction with green tea; (C) controls were given a conventional diet, placebo or similar calorie food as controls, and no extra physical activity or health education beyond everyday routines. (O) Body weight (BM), body mass index (BMI), waist circumference (WC), body fat percentage (BF%), plasma total cholesterol (TC), triglycerides (TG), high-density lipoprotein levels (HDL), and low-density lipoprotein levels (LDL) were among the outcomes measured after the intervention; (S) randomised controlled trials were utilized (RCTs). In order to ensure the integrity of the search, we also excluded conference abstracts, correspondence, reviews of works, pre-experimental protocols, literature reviews, crossover trial designs, and literature with unpublished data (but not literature with author permission), but we did not restrict the language.

### 2.3. Classification of interventions

Notably, because in several RCTs the intervention in the control group included some kind of exercise mixed with a placebo, we have neither categorized this as a “exercise alone group” nor have we simply merged it with the control group; rather, we have regarded it as a distinct categorization. This is discussed as a new category. The precise sorts of interventions are detailed in the **Table 1**[18].

**Table 1.** The characteristics of the different interventions.

| Name | Characteristics of the intervention |
| --- | --- |
| High intensity interval training (HIIT) | F : 3-5 times/week , 30min-60min/time<br>I: >65% VO<sub>2</sub>max or >65% HRR or >75% HRmax<br>T : ≥8weeks<br>The intensity of the exercises is high, and the intensity between sessions fluctuates: shorter intervals of greater intensity training (working groups) are alternated with longer periods of lower intensity resting groups, as opposed to total rest for the body. |
| Aerobic training (AEM) | F: 3-5 times/week. 30min-60min/time<br>I: 45%-65%VO<sub>2</sub>max or >50%-60% HRR or 65%-75% HRmax<br>T : ≥4 weeks<br>Less strenuous workouts are performed, with no respite between sessions and continuous training until the conclusion. |
| Resistance training (RT) | F : 3-5 times/week , 30min-60min/time<br>I : Average <80% 1RM<br>T : ≥4 weeks |
| Aerobic training combined with resistance training (COM) | Aerobic and resistance training in the same session, with no sequencing constraints. |
| Aerobic training combined with green tea intake (AEM+GTA) | Daily supplementation of decocted green tea or green tea extract in capsules/tablets in conjunction with aerobic training |
| Resistance training combined with green tea intake (RT+GTA) | Daily supplementation of decocted green tea or green tea extract in capsules/tablets in conjunction with resistance training |
| Green tea intake (GTA) | Daily supplementation of decocted green tea or green tea extract in capsules/tablets |
| Resistance training combined with placebo intake (RT+PLA) | Daily placebo supplements such as fibre or starch in conjunction with resistance training |
| Aerobic training combined with placebo intake (AEM+PLA) | Daily placebo supplements such as fibre or starch in conjunction with aerobic training |

Table notes: HRmax: maximum heart rate; HRR: heart rate reserve; VO<sub>2</sub>max: maximum oxygen uptake; RM : repetition maximum; F:frequency; I:intensity; T:times

### 2.4. Literature Screening

The two researchers used the program Zotero 5.0 (Corporation for Digital Scholarship, Vienna, VA, USA) to do a three-round screening of the whole body of literature. The procedure was the following: (1) Eliminate duplicate publications from various databases. (2) After reading the abstracts and titles, those that did not correspond to the sort of literature were discarded. (3) Searching for the complete text of the remaining literature and reading it, removing material once again, and then selecting the literature to be included[19].

### 2.5. Data extraction and quality assessment

A word table including seven parts was used for data extraction: (1) author’s name and year of publication; (2) gender characteristics of individuals (male and female, male only, female only) and sample size; (3) body mass index; (4) age; (5) interventions; and (7) sample size (experimental and control groups) (6) experiment length (7) Outcomes.

The Cochrane Handbook version 5.1.0 (Cochrane, London, UK) was used by two researchers to evaluate the quality of each chosen piece of literature[20]. The instrument featured seven potential risks: (1) generation of a random sequence, (2) concealment of treatment allocation, (3) participant and (4) staff blinding, (4) insufficient outcome data, (5) selective reporting, and (7) other types of bias. Based on the number of components for which high ROB was potentially present, trials were categorized into three degrees of ROB: high risk (five or more components), moderate risk (three or four components), and low risk (two or fewer components).

The selection of the literature, the extraction of data, and the evaluation of quality were independently performed by two writers, and any contentious areas of the procedure were evaluated and finalized by a third author[21].

### 2.6. Data analysis

By definition, we retrieved the mean (Mean) and standard deviation (SD) of the number of individuals participated in each RCT before and after the intervention. As the post-intervention impact, the mean and standard deviation at baseline were deducted from the mean and standard deviation after the intervention[22]. In addition, the missing standard deviations in the report were extracted using the formula[23].

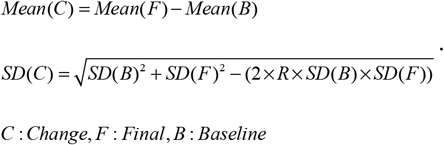

According to the definition of outcome indicators, a negative mean indicates a beneficial impact of treating overweight/obesity for WB, BMI, WC, BF%, TG, TC, and LDL, while a positive mean indicates a negative effect for HDL. In addition, since BM (in kg), BF% (in %), WC (in cm), and BMI (in kg/m2) have consistent units of measurement, MD (mean difference) was used for computation, however SMD (standardized mean difference) was utilized for TG, TC, LDL, HDL, as their units are not statistical[24].

An essential assumption of the NMA is the consistency and transferability of interventions; thus, before calculating the NMA, we conducted consistency and transferability tests on the combined data. I^2^ was used to evaluate inconsistency between studies, with I^2^ > 50% indicating high heterogeneity between studies and I^2^ <50% indicating low heterogeneity between studies. P values were used to evaluate transferability between studies, with P > 0.05 indicating good transferability between studies and P 0.05 indicating poor transferability between studies[25]. A horizontal line parallel to the horizontal axis in a funnel plot indicates a decreased probability of publishing bias, and vice versa[26].

The ‘network map’ diagram in STATA is used to visualize the connection between all studies, where each dot represents an intervention and each line indicates a direct comparison between two locations, or an indirect comparison through transmission if no line exists[27].

The creation of SUCRA (surface under the cumulative ranking curve) plots and P-values ranging from 0 to 1 is an essential feature of the NMA. In a comparison of various treatments, a P-value closer to 0 indicates that an intervention is less successful, while a P-value closer to 1 indicates that the intervention is more effective[28].

On the basis of the NMA calculations, a league table (network meta-analysis matrix) was constructed between the various treatments, which gives a visual depiction of the differences between the two comparisons of the various interventions. The diagonal lines across the table represent the different interventions, with the best intervention in the upper left corner and the worst intervention in the lower right corner. Below the diagonal lines are the estimates from the pairwise meta-analysis, where a 95% confidence interval (95% CI) in the estimate that does not contain ‘0’ indicates a statistically significant difference between the pairwise comparisons[29].

## 3. Results

### 3.1. Literature selection

The search approach described above yielded a total of 8899 original documents in the database, plus three more documents discovered by manual search. After deleting duplicates, 5734 documents remained; for this section of the literature, the abstract and title were read; after eliminating 5564 documents, 174 documents remained; for this section of the literature, the entire text was read; after eliminating 130 documents, 44 documents remained. The extraction of literature flowchart is shown in **Figure 1**.

**Figure 1.**
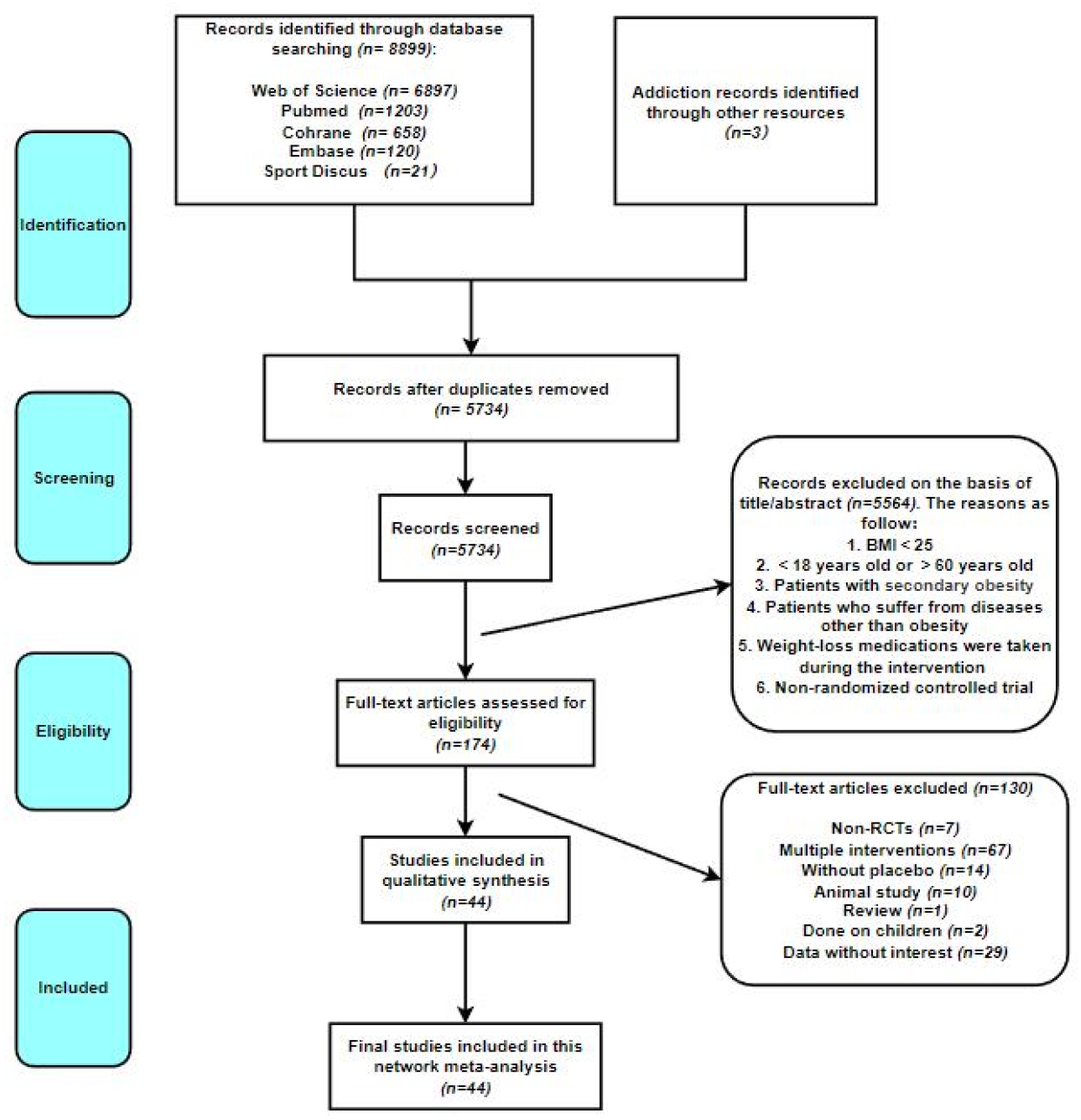
The full process of literature screening.

### 3.2. Characteristics of the included literature

There were 44 original studies included[30–73], with a total of 2001 individuals, of whom 62.3% were female. 11 studies reported aerobic exercise with green tea, 2 studies reported resistance training with green tea, 10 studies reported green tea alone, 9 studies reported low to moderate intensity aerobic exercise, 9 studies reported high intensity interval training, 13 studies reported resistance training, 8 studies reported combined exercise, 8 studies reported aerobic training with placebo, and 1 study reported resistance training with placebo. The **Supplementary Table 2** will present the details.

### 3.3. Quality assessment

18% (8 studies) of the studies were deemed high risk, 31% (14 studies) were deemed low risk, and some (51%) were deemed intermediate risk. We consider that despite the fact that moderate risk accounts for fifty percent of the total, it is acceptable since the difficulties of adopting a double-blind approach to participants for exercise as an intervention is intrinsic to it and not due to the experimental design. The **Supplementary Table 3** will present the details.

### 3.4. Network meta-analysis

Following NMA calculations, the corresponding network evidence maps were generated, with the size of the nodes proportional to the number of subjects who participated in the intervention and the thickness of the line segments proportional to the number of studies that compared the two interventions head-to-head. The **Figure 2** depicts network evidence maps for four anthropometric indicators (BM, BMI, BF%, WC), whereas **Figure 3** depicts network evidence maps for four lipid profile indicators (TG, TC, HDL, LDL). The SUCRA diagram depicts the effectiveness rating of each intervention for a particular outcome metric (**Supplementary Figure 2 to 9 and 26 to 33**). **Supplementary Figure 18 to 25** also depicts a forest patch between interventions. For each of the eight outcome measures, the inconsistency tests passed, and there was a high degree of transferability across treatments, as shown in the **Supplementary Figure 1 and Supplementary Table 4**. The effects of two-by-two comparisons of treatments will be given in a meta-analysis matrix (**Figure 4 and 5**) (mean difference/standardized mean difference + 95% confidence interval), with 95% CI excluding 0 representing statistically significant differences.

**Figure 2.**
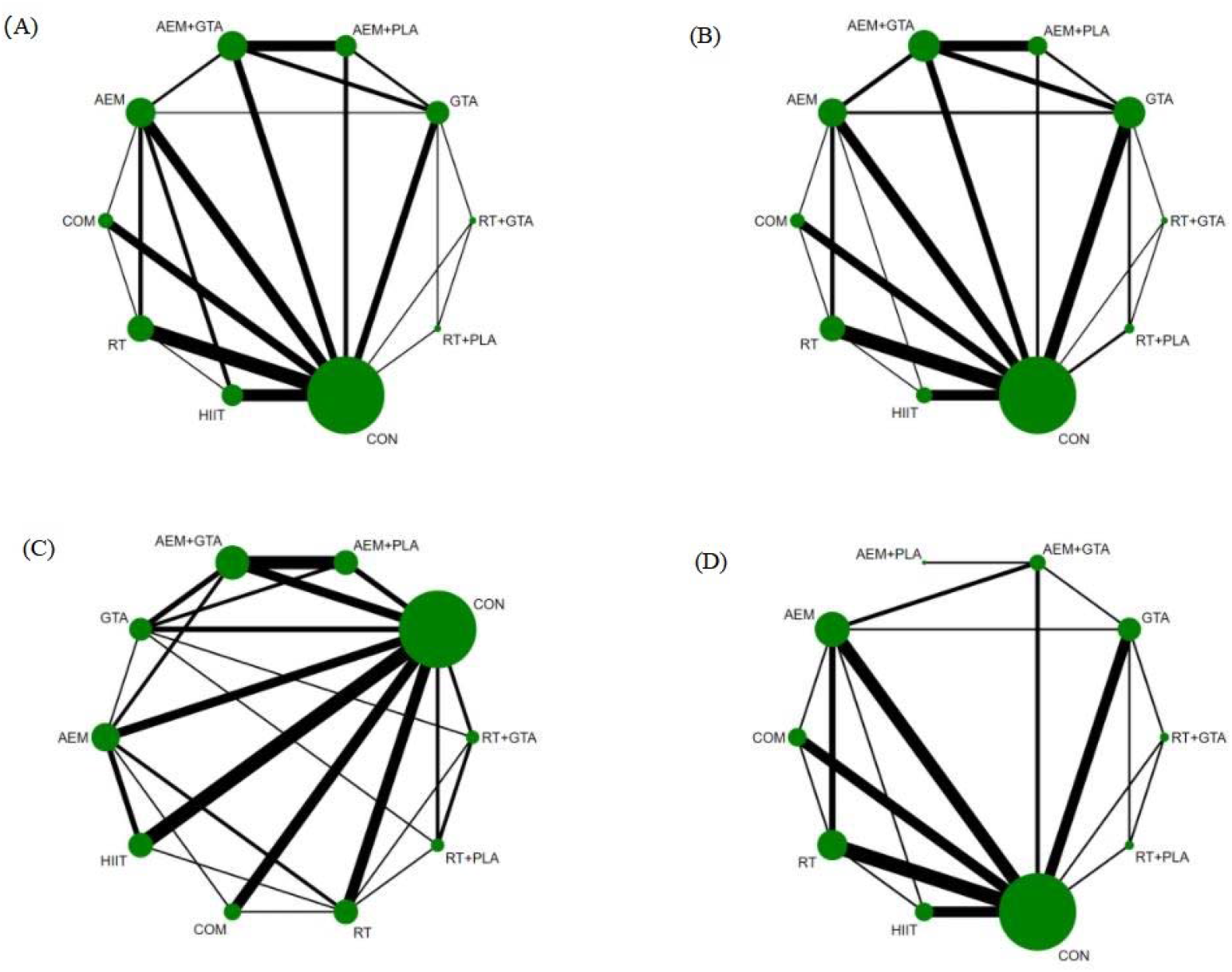
The network map of anthropometric indicators. (A): BW; (B): BMI; (C): BF%; (D): WC. AEM:aerobic exercise; RT: resistance training; HIIT: high-intensity interval training; GTA: green tea or green tea extract; AEM+GTA: aerobic training combined with green tea; RT+GTA: resistance training combined with green tea; AEM+PLA: aerobic training combined with placebo; RT+PLA: resistance training combined with placebo; COM: aerobic training combined with resistance training; CON: control group

**Figure 3.**
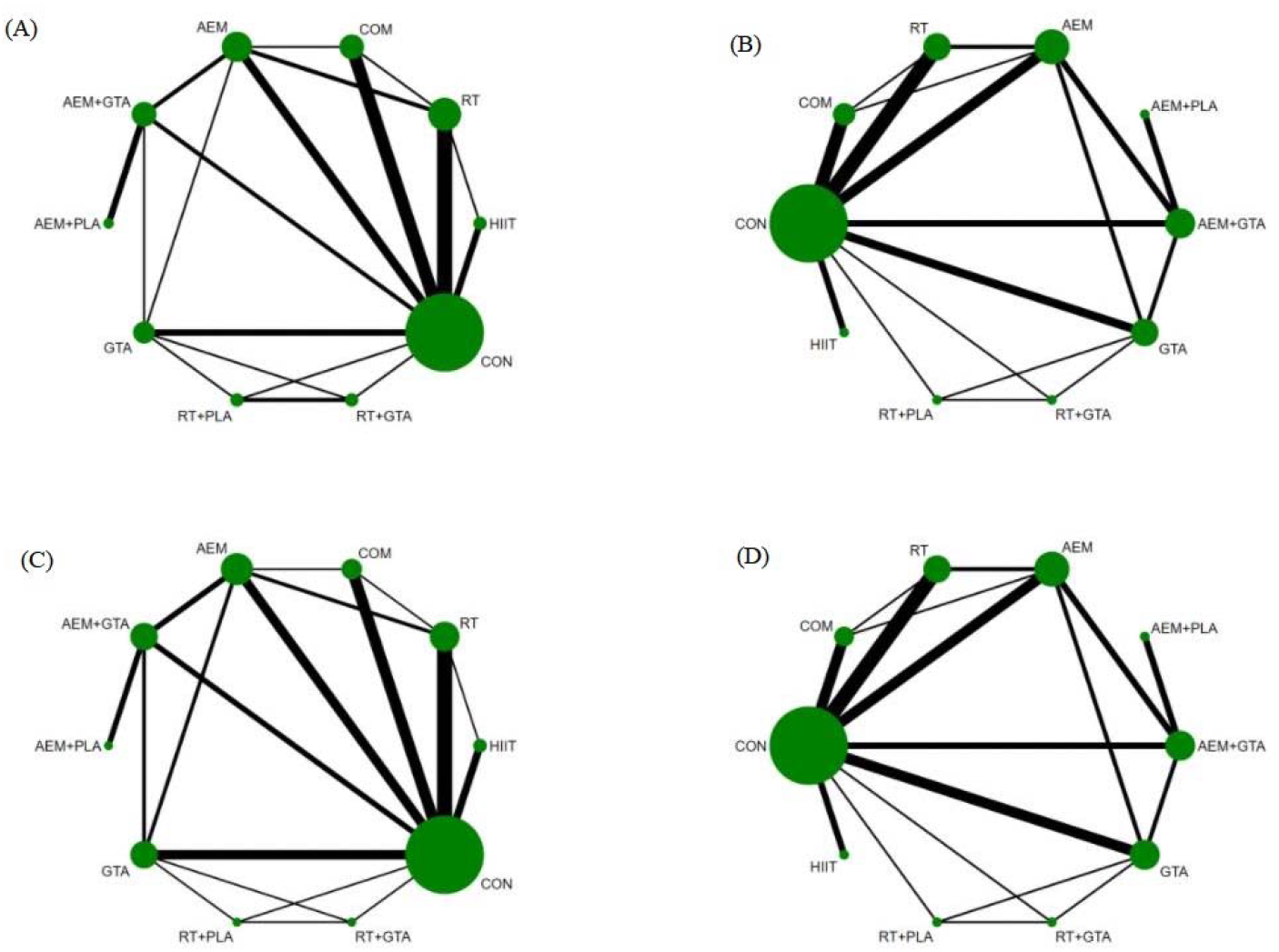
The network map of lipid profile indicators. (A): TG; (B): TC; (C): HDL; (D): LDL. AEM:aerobic exercise; RT: resistance training; HIIT: high-intensity interval training; GTA: green tea or green tea extract; AEM+GTA: aerobic training combined with green tea; RT+GTA: resistance training combined with green tea; AEM+PLA: aerobic training combined with placebo; RT+PLA: resistance training combined with placebo; COM: aerobic training combined with resistance training; CON: control group

**Figure 4.**
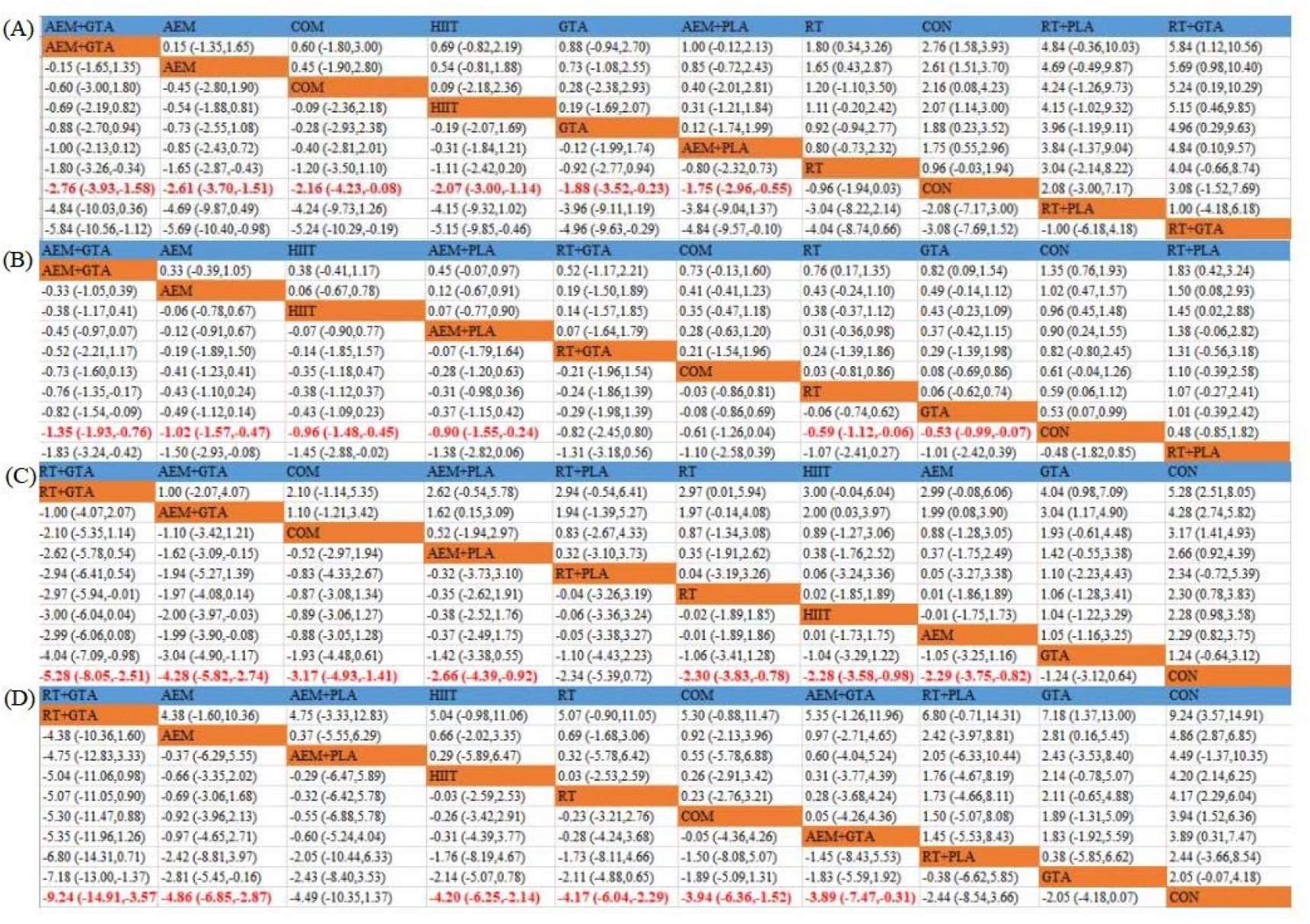
The network meta-analysis matrix of anthropometric indicators. (A): BW; (B): BMI; (C): BF%; (D): WC.

**Figure 5.**
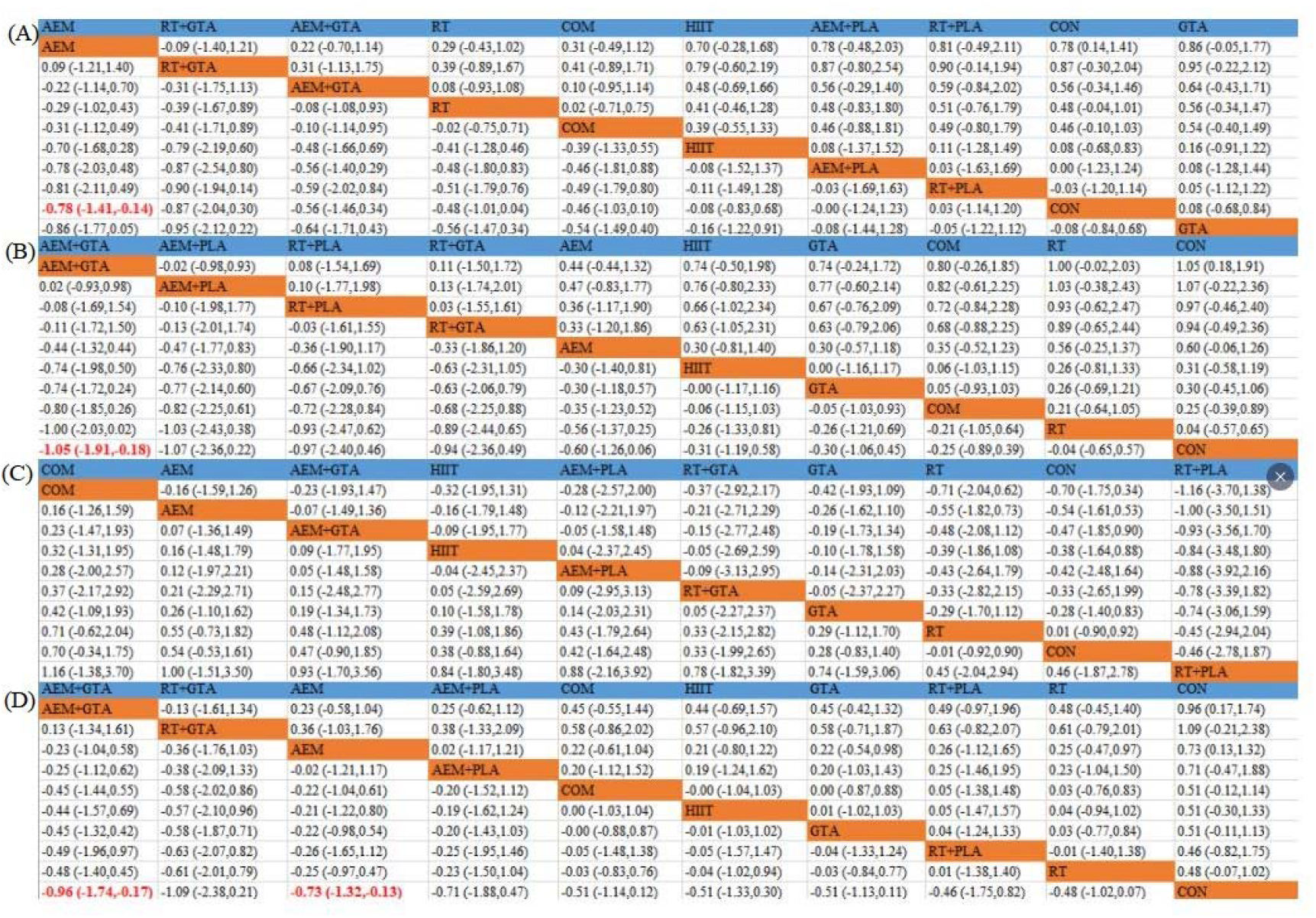
The network meta-analysis matrix of lipid profile indicators. (A): TG; (B): TC; (C): HDL; (D): LDL.

#### 3.4.1. BW

Interventions that included an aerobic component were at the top of the list for BW reduction, with AEM+GTA: MD=-2.76, 95%CI= (-3.93, -1.58); AEM: MD=-2.61, 95%CI= (-3.7. -1.51); COM: MD=-2.16, 95%CI= (-4.23, -0.08); HIIT: MD=-2.07, 95%CI= (-3.00, -1.14); GTA: MD=-1.88, 95%CI= (-3.52, -0.23); AEM+PLA: MD=-1.75, 95%CI= (-2.96, -0.55) efficacy was significant compared to the control group. Based on the SUCRA and meta-analysis matrix, AEM+GTA was probably the best intervention for weight reduction with SUCRA=86.9%. The SUCRA for each intervention can be found in the **supplementary table**.

#### 3.4.2. BMI

Interventions that included an aerobic exercise component were at the top of the list for BMI reduction, with AEM+GTA: MD=-1.35, 95%CI= (-1.93, -0.76); AEM: MD=-1.02, 95%CI= (-1.57, -0.47); HIIT: MD=-0.96, 95%CI= (-1.48, - 0.45); AEM+PLA: MD=-0.9, 95%CI= (-1.55, -0.24); RT: MD=-0.59, 95%CI= (-1.12, -0.06) efficacy was significant compared to the control group. Based on the SUCRA and meta-analysis matrix, AEM + GTA may be the best intervention to reduce BMI, SUCRA = 91.5%.

#### 3.4.3. BF%

All interventions were more effective than the control group in reducing the percentage of body fat, with RT+GTA: MD=-5.28, 95%CI= (-8.05, -2.51); AEM+GTA: MD=-4.28, 95%CI= (-5.82, -2.74); COM: MD=-3.17, 95%CI= (-4.93, -1.41); AEM+PLA: MD=-2.66, 95%CI= (-4.39, -0.92); RT: MD=-2.3, 95%CI= (-3.83, -0.78); HIIT: MD=-2.29, 95%CI= (-3.58, -0.98); AEM: MD=-2.29, 95%CI= (-3.75, -0.82); AEM: MD=-2.29, 95%CI= (-3.75, -0.82) 3.75, -0.82) were significant for efficacy. Based on the SUCRA and meta-analysis matrix, RT+GTA was probably the best intervention for reducing BF% with SUCRA= 94.1%.

#### 3.4.4. WC

All interventions were more effective than controls in reducing WC, with RT+GTA: MD=-9.24, 95%CI= (-14.91, -3.57); AEM: MD=-4.86, 95%CI= (-6.85, -2.87); HIIT: MD=-4.20, 95%CI= (-6.25, -2.14); RT: MD=-4.17, 95%CI= (-6.04, -2.29); COM: MD=-3.94, 95%CI= (-6.36, -1.52); AEM+GTA: MD=-3.89, 95%CI= (-7.47, -0.31) efficacy was significant compared to the control group. Based on the SUCRA and meta-analysis matrix, RT+GTA was probably the best intervention for reducing WC, SUCRA=95.1%.

#### 3.4.5.TG

In terms of TG reduction, only AEM was significant compared to the control group: SMD=-0.78, 95% CI= (-1.41, -0.14). The remaining interventions, although not significant, still showed an effect that could reduce triglyceride levels, with exercise combined with green tea at the top level. Based on the SUCRA and meta-analysis matrix, AEM was probably the best intervention for lowering TG with a SUCRA=81.6%.

#### 3.4.6. TC

In terms of TC reduction, only AEM+GTA was significant compared to the control group: SMD=-1.05, 95% CI=(-1.91,-0.18). The remaining interventions, although not statistically significantly different, still showed an effect in reducing TC, with exercise combined with green tea at the top level. Based on the SUCRA and meta-analysis matrix, AEM+GTA was probably the best intervention for reducing TG with SUCRA=79.2%.

#### 3.4.7. LDL

All interventions were more effective than controls in reducing LDL, with AEM+GTA: SMD=-0.96, 95%CI= (-1.74, -0.17); AEM: SMD=-0.73, 95%CI= (-1.32, -0.13) being significantly more effective compared to controls. Based on the SUCRA and meta-analysis matrix, AEM + GTA was probably the best intervention for LDL reduction with SUCRA = 76.6%.

#### 3.4.8. HDL

In terms of increasing HDL, all interventions did not show a significant increase compared to the control group. Based on the SUCRA and meta-analysis matrix, COM was probably the best intervention for reducing TG with a SUCRA = 70.5%.

### 3.5. Publication bias test

Visual inspection of the horizontal line in the middle of the funnel plot revealed no significant publication bias. The funnel plots of eight outcomes can find in the **Supplementary Figure 10 to 17**.

## 4. Discussion

Numerous prior research and meta-analyses have evaluated the benefits of GTA and exercise on decreasing overweight/obesity[74–77], but they have overlooked a key intervention, namely the combination of exercise and green tea drinking. This intervention was evaluated with other treatments for the first time in this NMA. The interventions that included an aerobic component resulted in significant weight loss; however, resistance training combined with green tea did not outperform the control group. We hypothesize that the effect of resistance training on increasing lean body mass counteracted the majority of the weight loss caused by green tea, consistent with a recent Grainne study comparing the effects of multiple exercise modalities for overweight/obesity. This is similar with the findings of Grainne’s recent NMA research assessing the impact of various exercise methods on overweight/obesity[23]. However, we do not consider resistance training paired with green tea to be ineffective or inefficient, since resistance training combined with green tea came out on top in terms of P scores for body fat percentage decrease and waist circumference reduction.

Several therapies that incorporated an aerobic component topped the SUCRA rankings for BMI reduction, although it is important to note that while the difference from the control group was statistically significant, the range of means was narrower, from -0.05 to -1.00.

According to a recent research, BF% is more sensitive than BMI to changes in body fat, the most harmful body tissue, and BF% is more responsive to exercise[78]. Previous meta-analyses have also demonstrated that green tea consumption reduces BF% to some degree[79], and our NMA demonstrated that exercise combined with green tea achieved the best P-score for reducing BF%; therefore, we propose that regular green tea consumption combined with increased physical activity may be more effective than green tea alone or increased physical activity alone.

In addition, WC has been reported to be a better predictor than BMI, since abdominal fat is a significant independent correlate of all-cause mortality excluding total body fat[80], and Lin reported in his study that green tea consumption did not show a significant reduction in WC, which is consistent with our NMA, but Lin also concluded that there was a benefit of green tea consumption in reducing visceral fat[77], and our NMA supports this conclusion. The NMA reveals that the combination of resistance exercise and green tea drinking led to a considerable decrease in waist circumference. Considering that abdominal fat (produced by the deposition of subcutaneous and visceral fat) is more strongly connected with stroke and deep vein thrombosis in obese individuals, resistance exercise coupled with green tea administration is recommended for the management of abdominal fat. Exercise causes an increase in serum levels of the prototypical muscle factor IL-6, which helps to suppress central feeding mechanisms and reduces appetite to a certain extent[81]; and green tea’s highest content of catechins, which are flavanol polyphenolic compounds, are largely not absorbed by the small intestine and mostly enter the colon, where they combine with lipids and lipase, inhibiting lipid absorption and exercise-induced red blood cell synthesis[82]. This is supplemented by the anti-absorption activity of green tea catechins, which together aid in the treatment of excessive fat buildup in the body.

In addition to the more visible anthropometric characteristics mentioned previously, recent studies indicate that overweight/obesity is frequently associated with abnormal lipid metabolism, which not only increases the risk of cardiovascular disease but is also an independent risk factor for coronary artery disease and ischaemic stroke[83]. In light of this, the effects of various treatments on the human lipid profile were also examined in our NMA. According to a previous big study, green tea intake was inversely related to cardiovascular risk variables, a conclusion that was partly supported by the current research[84,85]. Obese patients are more likely to have abnormally high plasma total cholesterol levels[86], and Xu concluded in his meta-analysis that green tea consumption was associated with a significant reduction in TC levels[87]. However, in our NMA, green tea consumption, while reducing TC levels to some extent, did not reduce them as much as exercise or exercise combined with green tea consumption. High LDL levels may cause the cholesterol it transports to accumulate on artery walls and create atherosclerosis[88]. Aerobic exercise mixed with green tea or aerobic training significantly reduced LDL levels in this NMA. According to previous research, both exercise and green tea catechins (particularly EGCG) stimulate AMPK activity[89,90], decrease plasma and liver fat concentrations by upregulating glucose carrier (GLUT4) expression, stimulate hepatic fat oxidation, and decrease lipid production in adipose tissue and lipid droplet size in the liver. However, in terms of boosting HDL levels, we did not notice a substantial advantage from the green tea-related intervention; rather, aerobic and resistance training had the greatest P values.

Moreover, according to some researches, overweight/obesity may be an inflammatory disease owing to the overproduction of TNF-α cytokines, and both exercise and EGCG have a partial inhibitory impact on TNF-α, i.e. both have anti-overweight/obesity inflammatory effects[36,39,91]. Due to the limited number of relevant research, it is desired that future studies would build on this review in order to consolidate and investigate the impact size of TNF-α modification in this NMA.

At last, it is important to note that many prior studies have revealed that HIIT is useful as an exercise intervention for the treatment of overweight/obesity, and our NMA confirms this, but it is not as beneficial as exercise coupled with green tea intake for the treatment of overweight/obesity[92]. Moreover, according to Grainne ‘s NMA, a combination of aerobic and resistance training appears to have the greatest effect on reducing abdominal fat and improving lean body mass, and we actively sought RCTs on this combination of exercise and green tea in our literature. However, we were unable to locate any RCTs that reported on COM+GTA, so we hope that future research will focus on this intervention. More research will concentrate on this intervention.

## 5. Strength and limitation

The NMA has a higher sample size (n=44) and number of individuals (n=2001), as well as a greater number of meaningful intervention effects for people to examine. The NMA also addresses a gap in the literature by comparing the intervention of exercise paired with green tea drinking to similar therapies. In addition, we have modified the subject characteristics to eliminate minors, the old population, and those with overweight/obesity in combination with other diseases, focusing on non-elderly adults with overweight/obesity. This makes the NMA’s evidence-based medical recommendations to professionals and patients more precise. However, and this may be a disadvantage, we did not include the entire population for NMA, and the number of RCTs reporting RT+NMA was small compared to other interventions. Furthermore, interventions that include an exercise component are difficult to conduct double-blind due to the need for participants to sign an informed consent form prior to the experiment, which is a limitation of exercise interventions. This NMA does not account for cardiorespiratory fitness or cytokine markers (TNF-α, IL-1, IL-6, etc.) since the sample size was insufficient. The NMA focuses more on the comparative efficacy of green tea supplementation, exercise supplementation, and exercise supplementation combined with green tea as interventions, and does not explore interventions such as energy-restricted diets and supplementation with other dietary supplements that may be equally effective, although they have been reported; therefore, readers should approach this NMA with caution.

## 6. Conclusion

Based on the NMA calculations and SUCRA probability rankings, we consider that for non-elderly individuals seeking non-pharmaceutical or surgical therapies for overweight/obesity, consuming green tea in conjunction with increased physical activity is preferable than taking green tea or exercising alone. In addition, combining green tea with exercise is an alternative complementary treatment that should be promoted and tested in clinical settings.

## Data Availability

All data produced in the present study are available upon reasonable request to the authors

## References

1. Arroyo-Johnson, C.; Mincey, K.D. Obesity Epidemiology Worldwide. Gastroenterol. Clin. North Am. 2016, 45, 571-+, doi:10.1016/j.gtc.2016.07.012.

2. Bray, G.A.; Heisel, W.E.; Afshin, A.; Jensen, M.D.; Dietz, W.H.; Long, M.; Kushner, R.F.; Daniels, S.R.; Wadden, T.A.; Tsai, A.G.; et al. The Science of Obesity Management: An Endocrine Society Scientific Statement. Endocr. Rev. 2018, 39, 79–132, doi:10.1210/er.2017-00253.

3. Ortega, F.B.; Lavie, C.J.; Blair, S.N. Obesity and Cardiovascular Disease. Circ. Res. 2016, 118, 1752–1770, doi:10.1161/CIRCRESAHA.115.306883.

4. Yumuk, V.; Tsigos, C.; Fried, M.; Schindler, K.; Busetto, L.; Micic, D.; Toplak, H. European Guidelines for Obesity Management in Adults. Obes. Facts 2015, 8, 402–424, doi:10.1159/000442721.

5. Hall, K.D.; Kahan, S. Maintenance of Lost Weight and Long-Term Management of Obesity. Med. Clin. North Am. 2018, 102, 183-+, doi:10.1016/j.mcna.2017.08.012.

6. Powell-Wiley, T.M.; Poirier, P.; Burke, L.E.; Despres, J.-P.; Gordon-Larsen, P.; Lavie, C.J.; Lear, S.A.; Ndumele, C.E.; Neeland, I.J.; Sanders, P.; et al. Obesity and Cardiovascular Disease: A Scientific Statement From the American Heart Association. Circulation 2021, 143, E984–E1010, doi:10.1161/CIR.0000000000000973.

7. Bessesen, D.H.; Van Gaal, L.F. Progress and Challenges in Anti-Obesity Pharmacotherapy. Lancet Diabetes Endocrinol. 2018, 6, 237–248, doi:10.1016/S2213-8587(17)30236-X.

8. Hill, J.O.; Wyatt, H.R.; Peters, J.C. Energy Balance and Obesity. Circulation 2012, 126, 126–132, doi:10.1161/CIRCULATIONAHA.111.087213.

9. Legeay, S.; Rodier, M.; Fillon, L.; Faure, S.; Clere, N. Epigallocatechin Gallate: A Review of Its Beneficial Properties to Prevent Metabolic Syndrome. Nutrients 2015, 7, 5443–5468, doi:10.3390/nu7075230.

10. Yang, C.S.; Zhang, J.; Zhang, L.; Huang, J.; Wang, Y. Mechanisms of Body Weight Reduction and Metabolic Syndrome Alleviation by Tea. Mol. Nutr. Food Res. 2016, 60, 160–174, doi:10.1002/mnfr.201500428.

11. Boschmann, M.; Thielecke, F. The Effects of Epigallocatechin-3-Gallate on Thermogenesis and Fat Oxidation in Obese Men: A Pilot Study. J. Am. Coll. Nutr. 2007, 26, 389S–395S, doi:10.1080/07315724.2007.10719627.

12. Chatree, S.; Sitticharoon, C.; Maikaew, P.; Pongwattanapakin, K.; Keadkraichaiwat, I.; Churintaraphan, M.; Sripong, C.; Sririwichitchai, R.; Tapechum, S. Epigallocatechin Gallate Decreases Plasma Triglyceride, Blood Pressure, and Serum Kisspeptin in Obese Human Subjects. Exp. Biol. Med. 2021, 246, 163–176, doi:10.1177/1535370220962708.

13. Dludla, P.V.; Nkambule, B.B.; Jack, B.; Mkandla, Z.; Mutize, T.; Silvestri, S.; Orlando, P.; Tiano, L.; Louw, J.; Mazibuko-Mbeje, S.E. Inflammation and Oxidative Stress in an Obese State and the Protective Effects of Gallic Acid. Nutrients 2019, 11, 23, doi:10.3390/nu11010023.

14. Rodriguez-Perez, C.; Segura-Carretero, A.; del Mar Contreras, M. Phenolic Compounds as Natural and Multifunctional Anti-Obesity Agents: A Review. Crit. Rev. Food Sci. Nutr. 2019, 59, 1212–1229, doi:10.1080/10408398.2017.1399859.

15. Khoo, W.Y.; Chrisfield, B.J.; Sae-tan, S.; Lambert, J.D. Mitigation of Nonalcoholic Fatty Liver Disease in High-Fat-Fed Mice by the Combination of Decaffeinated Green Tea Extract and Voluntary Exercise. J. Nutr. Biochem. 2020, 76, 108262, doi:10.1016/j.jnutbio.2019.108262.

16. Sun, Y.; Wang, Y.; Song, P.; Wang, H.; Xu, N.; Wang, Y.; Zhang, Z.; Yue, P.; Gao, X. Anti-Obesity Effects of Instant Fermented Teas in Vitro and in Mice with High-Fat-Diet-Induced Obesity. Food Funct. 2019, 10, 3502–3513, doi:10.1039/c9fo00162j.

17. Methley, A.M.; Campbell, S.; Chew-Graham, C.; McNally, R.; Cheraghi-Sohi, S. PICO, PICOS and SPIDER: A Comparison Study of Specificity and Sensitivity in Three Search Tools for Qualitative Systematic Reviews. Bmc Health Serv. Res. 2014, 14, 579, doi:10.1186/s12913-014-0579-0.

18. Thompson, P.D.; Arena, R.; Riebe, D.; Pescatello, L.S. ACSM’s New Preparticipation Health Screening Recommendations from ACSM’s Guidelines for Exercise Testing and Prescription, Ninth Edition. Curr. Sports Med. Rep. 2013, 12, 215–217, doi:10.1249/JSR.0b013e31829a68cf.

19. Panic, N.; Leoncini, E.; de Belvis, G.; Ricciardi, W.; Boccia, S. Evaluation of the Endorsement of the Preferred Reporting Items for Systematic Reviews and Meta-Analysis (PRISMA) Statement on the Quality of Published Systematic Review and Meta-Analyses. Plos One 2013, 8, e83138, doi:10.1371/journal.pone.0083138.

20. Armijo-Olivo, S.; Stiles, C.R.; Hagen, N.A.; Biondo, P.D.; Cummings, G.G. Assessment of Study Quality for Systematic Reviews: A Comparison of the Cochrane Collaboration Risk of Bias Tool and the Effective Public Health Practice Project Quality Assessment Tool: Methodological Research. J. Eval. Clin. Pract. 2012, 18, 12–18, doi:10.1111/j.1365-2753.2010.01516.x.

21. Page, M.J.; McKenzie, J.E.; Bossuyt, P.M.; Boutron, I.; Hoffmann, T.C.; Mulrow, C.D.; Shamseer, L.; Tetzlaff, J.M.; Akl, E.A.; Brennan, S.E.; et al. The PRISMA 2020 Statement: An Updated Guideline for Reporting Systematic Reviews. Int. J. Surg. 2021, 88, 105906, doi:10.1016/j.ijsu.2021.105906.

22. White, I.R. Network Meta-Analysis. Stata J. 2015, 15, 951–985, doi:10.1177/1536867X1501500403.

23. O’Donoghue, G.; Blake, C.; Cunningham, C.; Lennon, O.; Perrotta, C. What Exercise Prescription Is Optimal to Improve Body Composition and Cardiorespiratory Fitness in Adults Living with Obesity? A Network Meta-Analysis. Obes. Rev. 2021, 22, e13137, doi:10.1111/obr.13137.

24. Huhn, M.; Nikolakopoulou, A.; Schneider-Thoma, J.; Krause, M.; Samara, M.; Peter, N.; Arndt, T.; Backers, L.; Rothe, P.; Cipriani, A.; et al. Comparative Efficacy and Tolerability of 32 Oral Antipsychotics for the Acute Treatment of Adults with Multi-Episode Schizophrenia: A Systematic Review and Network Meta-Analysis. Lancet 2019, 394, 939–951, doi:10.1016/S0140-6736(19)31135-3.

25. Rouse, B.; Chaimani, A.; Li, T. Network Meta-Analysis: An Introduction for Clinicians. Intern. Emerg. Med. 2017, 12, 103–111, doi:10.1007/s11739-016-1583-7.

26. Lin, L. Graphical Augmentations to Sample-Size-Based Funnel Plot in Meta-Analysis. Res. Synth. Methods 2019, 10, 376–388, doi:10.1002/jrsm.1340.

27. Chaimani, A.; Salanti, G. Visualizing Assumptions and Results in Network Meta-Analysis: The Network Graphs Package. Stata J. 2015, 15, 905–950, doi:10.1177/1536867X1501500402.

28. Ruecker, G.; Schwarzer, G. Ranking Treatments in Frequentist Network Meta-Analysis Works without Resampling Methods. Bmc Med. Res. Methodol. 2015, 15, 58, doi:10.1186/s12874-015-0060-8.

29. Ma, C.; Avenell, A.; Bolland, M.; Hudson, J.; Stewart, F.; Robertson, C.; Sharma, P.; Fraser, C.; MacLennan, G. Effects of Weight Loss Interventions for Adults Who Are Obese on Mortality, Cardiovascular Disease, and Cancer: Systematic Review and Meta-Analysis. Bmj-Br. Med. J. 2017, 359, j4849, doi:10.1136/bmj.j4849.

30. Venkatakrishnan, K.; Chiu, H.-F.; Cheng, J.-C.; Chang, Y.-H.; Lu, Y.-Y.; Han, Y.-C.; Shen, Y.-C.; Tsai, K.-S.; Wang, C.-K. Comparative Studies on the Hypolipidemic, Antioxidant and Hepatoprotective Activities of Catechin-Enriched Green and Oolong Tea in a Double-Blind Clinical Trial. Food Funct. 2018, 9, 1205–1213, doi:10.1039/C7FO01449J.

31. Afzalpour, M.E.; Ghasemi, E.; Zarban, A. Effects of 10 Weeks of High Intensity Interval Training and Green Tea Supplementation on Serum Levels of Sirtuin-1 and Peroxisome Proliferator-Activated Receptor Gamma Co-Activator 1-Alpha in Overweight Women. Sci. Sports 2017, 32, 82–90, doi:10.1016/j.scispo.2016.09.004.

32. Abdulghani, M. Effects of Green Tea on the Body Weight of Malaysian Young Obese Females: Single Blind Clinical Trial Study.

33. Suliburska, J.; Bogdanski, P.; Szulinska, M.; Stepien, M.; Pupek-Musialik, D.; Jablecka, A. Effects of Green Tea Supplementation on Elements, Total Antioxidants, Lipids, and Glucose Values in the Serum of Obese Patients. Biol. Trace Elem. Res. 2012, 149, 315–322, doi:10.1007/s12011-012-9448-z.

34. Nabi, B.N.; Sedighinejad, A.; Haghighi, M.; Farzi, F.; Rimaz, S.; Atrkarroushan, Z.; Biazar, G. The Anti-Obesity Effects of Green Tea: A Controlled, Randomized, Clinical Trial. Iran. Red Crescent Med. J. 2018, 20, e55950, doi:10.5812/ircmj.55950.

35. Ghasemi, E.; Afzalpour, M.E.; Nayebifar, S. Combined High-Intensity Interval Training and Green Tea Supplementation Enhance Metabolic and Antioxidant Status in Response to Acute Exercise in Overweight Women. J. Physiol. Sci. 2020, 70, 31, doi:10.1186/s12576-020-00756-z.

36. Bagheri, R.; Rashidlamir, A.; Ashtary-Larky, D.; Wong, A.; Alipour, M.; Motevalli, M.S.; Chebbi, A.; Laher, I.; Zouhal, H. Does Green Tea Extract Enhance the Anti-Inflammatory Effects of Exercise on Fat Loss? Br. J. Clin. Pharmacol. 2020, 86, 753–762, doi:10.1111/bcp.14176.

37. Zhang, T.; Li, N.; Chen, S.; Hou, Z.; Saito, A. Effects of Green Tea Extract Combined with Brisk Walking on Lipid Profiles and the Liver Function in Overweight and Obese Men: A Randomized, Double-Blinded, Placebo-Control Trial. An. Acad. Bras. Ciênc. 2020, 92, doi:10.1590/0001-3765202020191594.

38. Bagheri, R.; Rashidlamir, A.; Ashtary-Larky, D.; Wong, A.; Grubbs, B.; Motevalli, M.S.; Baker, J.S.; Laher, I.; Zouhal, H. Effects of Green Tea Extract Supplementation and Endurance Training on Irisin, pro-Inflammatory Cytokines, and Adiponectin Concentrations in Overweight Middle-Aged Men. Eur. J. Appl. Physiol. 2020, 120, 915–923, doi:10.1007/s00421-020-04332-6.

39. Alikhani, S.; Etemad, Z.; Azizbeigi, K. Effects of Spinning Workout and Green Tea Consumption on the Anti-Inflammatory and Inflammatory Markers and Body Composition of Overweight Women. J. Kermanshah Univ. Med. Sci. 2021, 25, doi:10.5812/jkums.110116.

40. Ors, E.D.; Goktas, Z. Herbal Dietary Interventions for Weight Loss among Regularly Exercising Women in Turkey. Nutr. Food Sci. 2021, 51, 1272–1281, doi:10.1108/NFS-02-2021-0064.

41. Ghadami, A.; Abedi, B.; Abarghoee, J.P.; Rarani, S.A. The Combined Effect of Resistance Training and Green Tea Supplements on the Lipid Profile and Anthropometric Indices of Overweight and Obese Males. Zahedan J. Res. Med. Sci. 2018, 20, doi:10.5812/zjrms.10698.

42. Amozadeh, H.; Shabani, R.; Nazari, M. The Effect of Aerobic Training and Green Tea Supplementation on Cardio Metabolic Risk Factors in Overweight and Obese Females: A Randomized Trial. Int. J. Endocrinol. Metab. 2018, 16, e60738, doi:10.5812/ijem.60738.

43. Gahreman, D.; Heydari, M.; Boutcher, Y.; Freund, J.; Boutcher, S. The Effect of Green Tea Ingestion and Interval Sprinting Exercise on the Body Composition of Overweight Males: A Randomized Trial. Nutrients 2016, 8, 510, doi:10.3390/nu8080510.

44. Hematinezhad Touli, M.; Elmieh, A.; Hosseinpour, A. The Effect of Six-Week Aerobic Exercise Combined with Green Tea Consumption on PON1 and VO2max Increase and Apelin, Blood Pressure, and Blood Lipids Reduction in Young Obese Men. Arch. Razi Inst. 2022, 77, 2103–2111, doi:10.22092/ari.2022.357847.2109.

45. Cardoso, G.A.; Salgado, J.M.; Cesar, M. de C.; Donado-Pestana, C.M. The Effects of Green Tea Consumption and Resistance Training on Body Composition and Resting Metabolic Rate in Overweight or Obese Women. J. Med. Food 2013, 16, 120–127, doi:10.1089/jmf.2012.0062.

46. Roberts, J.D.; Willmott, A.G.B.; Beasley, L.; Boal, M.; Davies, R.; Martin, L.; Chichger, H.; Gautam, L.; Del Coso, J. The Impact of Decaffeinated Green Tea Extract on Fat Oxidation, Body Composition and Cardio-Metabolic Health in Overweight, Recreationally Active Individuals. Nutrients 2021, 13, 764, doi:10.3390/nu13030764.

47. Rostamian, M.; Bijeh, N. The The Effect of Short-Term Aerobic Exercise and Green Tea Consumption on MFO, Fatmax, Body Composition and Lipid Profile in Sedentary Postmenopausal women. Int. J. Appl. Exerc. Physiol. 2017, 6, 21–31, doi:10.22631/ijaep.v6i1.100.

48. Said, M.; Abdelmoneim, A.; Alibrahim, M.S.; Kotb, A.A.H. Aerobic Training, Resistance Training, or Their Combination as a Means to Fight against Excess Weight and Metabolic Syndrome in Obese Students — Which Is the Most Effective Modality? A Randomized Controlled Trial. Appl. Physiol. Nutr. Metab. 2021, 46, 952–963, doi:10.1139/apnm-2020-0972.

49. Beer, N.J.; Jackson, B.; Dimmock, J.A.; Guelfi, K.J. Attenuation of Post-Exercise Energy Intake Following 12 Weeks of Sprint Interval Training in Men and Women with Overweight. Nutrients 2022, 14, 1362, doi:10.3390/nu14071362.

50. Atashak, S.; Stannard, S.R.; Azizbeigi, K. Cardiovascular Risk Factors Adaptation to Concurrent Training in Overweight Sedentary Middle-Aged Men. J. Sports Med. Phys. Fitness 2016, 56, 624–630.

51. Franklin, N.C.; Robinson, A.T.; Bian, J.-T.; Ali, M.M.; Norkeviciute, E.; McGinty, P.; Phillips, S.A. Circuit Resistance Training Attenuates Acute Exertion-Induced Reductions in Arterial Function but Not Inflammation in Obese Women. Metab. Syndr. Relat. Disord. 2015, 13, 227–234, doi:10.1089/met.2014.0135.

52. Bonfante, I.L.P.; Chacon-Mikahil, M.P.T.; Brunelli, D.T.; Gáspari, A.F.; Duft, R.G.; Lopes, W.A.; Bonganha, V.; Libardi, C.A.; Cavaglieri, C.R. Combined Training, FNDC5/Irisin Levels and Metabolic Markers in Obese Men: A Randomised Controlled Trial. Eur. J. Sport Sci. 2017, 17, 629–637, doi:10.1080/17461391.2017.1296025.

53. Christensen, R.H.; Wedell-Neergaard, A.-S.; Lehrskov, L.L.; Legaard, G.E.; Dorph, E.; Larsen, M.K.; Launbo, N.; Fagerlind, S.R.; Seide, S.K.; Nymand, S.; et al. Effect of Aerobic and Resistance Exercise on Cardiac Adipose Tissues: Secondary Analyses From a Randomized Clinical Trial. JAMA Cardiol. 2019, 4, 778–787, doi:10.1001/jamacardio.2019.2074.

54. Dupuit, M.; Rance, M.; Morel, C.; Bouillon, P.; Boscaro, A.; Martin, V.; Vazeille, E.; Barnich, N.; Chassaing, B.; Boisseau, N. Effect of Concurrent Training on Body Composition and Gut Microbiota in Postmenopausal Women with Overweight or Obesity. Med. Sci. Sports Exerc. 2022, 54, 517–529, doi:10.1249/MSS.0000000000002809.

55. Keating, S.E.; Hackett, D.A.; Parker, H.M.; Way, K.L.; O’Connor, H.T.; Sainsbury, A.; Baker, M.K.; Chuter, V.H.; Caterson, I.D.; George, J.; et al. Effect of Resistance Training on Liver Fat and Visceral Adiposity in Adults with Obesity: A Randomized Controlled Trial. Hepatol. Res. Off. J. Jpn. Soc. Hepatol. 2017, 47, 622–631, doi:10.1111/hepr.12781.

56. Jang, S.-H.; Paik, I.-Y.; Ryu, J.-H.; Lee, T.-H.; Kim, D.-E. Effects of Aerobic and Resistance Exercises on Circulating Apelin-12 and Apelin-36 Concentrations in Obese Middle-Aged Women: A Randomized Controlled Trial. BMC Womens Health 2019, 19, 23, doi:10.1186/s12905-019-0722-5.

57. Oh, M.; Kim, S.; An, K.-Y.; Min, J.; Yang, H.I.; Lee, J.; Lee, M.K.; Kim, D.-I.; Lee, H.-S.; Lee, J.-W.; et al. Effects of Alternate Day Calorie Restriction and Exercise on Cardio-Metabolic Risk Factors in Overweight and Obese Adults: An Exploratory Randomized Controlled Study. BMC Public Health 2018, 18, 1124, doi:10.1186/s12889-018-6009-1.

58. Hu, J.; Liu, M.; Yang, R.; Wang, L.; Liang, L.; Yang, Y.; Jia, S.; Chen, R.; Liu, Q.; Ren, Y.; et al. Effects of High-Intensity Interval Training on Improving Arterial Stiffness in Chinese Female University Students with Normal Weight Obese: A Pilot Randomized Controlled Trial. J. Transl. Med. 2022, 20, 60, doi:10.1186/s12967-022-03250-9.

59. Nikseresht, M.; Agha-Alinejad, H.; Azarbayjani, M.A.; Ebrahim, K. Effects of Nonlinear Resistance and Aerobic Interval Training on Cytokines and Insulin Resistance in Sedentary Men Who Are Obese. J. Strength Cond. Res. 2014, 28, 2560–2568, doi:10.1519/JSC.0000000000000441.

60. Streb, A.R.; da Silva, R.P.; Leonel, L. dos S.; Possamai, L.T.; Gerage, A.M.; Turnes, T.; Del Duca, G.F. Effects of Nonperiodized and Linear Periodized Combined Training on Health-Related Physical Fitness in Adults With Obesity: A Randomized Controlled Trial. J. Strength Cond. Res. 2022, 36, 2628–2634, doi:10.1519/JSC.0000000000003859.

61. Donges, C.E.; Duffield, R.; Drinkwater, E.J. Effects of Resistance or Aerobic Exercise Training on Interleukin-6, C-Reactive Protein, and Body Composition. Med. Sci. Sports Exerc. 2010, 42, 304–313, doi:10.1249/MSS.0b013e3181b117ca.

62. Croymans, D.M.; Krell, S.L.; Oh, C.S.; Katiraie, M.; Lam, C.Y.; Harris, R.A.; Roberts, C.K. Effects of Resistance Training on Central Blood Pressure in Obese Young Men. J. Hum. Hypertens. 2014, 28, 157–164, doi:10.1038/jhh.2013.81.

63. Ahmadizad, S.; Ghorbani, S.; Ghasemikaram, M.; Bahmanzadeh, M. Effects of Short-Term Nonperiodized, Linear Periodized and Daily Undulating Periodized Resistance Training on Plasma Adiponectin, Leptin and Insulin Resistance. Clin. Biochem. 2014, 47, 417–422, doi:10.1016/j.clinbiochem.2013.12.019.

64. Winn, N.C.; Liu, Y.; Rector, R.S.; Parks, E.J.; Ibdah, J.A.; Kanaley, J.A. Energy-Matched Moderate and High Intensity Exercise Training Improves Nonalcoholic Fatty Liver Disease Risk Independent of Changes in Body Mass or Abdominal Adiposity - A Randomized Trial. Metabolism. 2018, 78, 128–140, doi:10.1016/j.metabol.2017.08.012.

65. Arad, A.D.; DiMenna, F.J.; Thomas, N.; Tamis-Holland, J.; Weil, R.; Geliebter, A.; Albu, J.B. High-Intensity Interval Training without Weight Loss Improves Exercise but Not Basal or Insulin-Induced Metabolism in Overweight/Obese African American Women. J. Appl. Physiol. Bethesda Md 1985 2015, 119, 352–362, doi:10.1152/japplphysiol.00306.2015.

66. Sun, J.; Cheng, W.; Fan, Z.; Zhang, X. Influence of High-Intensity Intermittent Training on Glycolipid Metabolism in Obese Male College Students. Ann. Palliat. Med. 2020, 9, 2013–2019, doi:10.21037/apm-20-1105.

67. Ataeinosrat, A.; Saeidi, A.; Abednatanzi, H.; Rahmani, H.; Daloii, A.A.; Pashaei, Z.; Hojati, V.; Basati, G.; Mossayebi, A.; Laher, I.; et al. Intensity Dependent Effects of Interval Resistance Training on Myokines and Cardiovascular Risk Factors in Males With Obesity. Front. Endocrinol. 2022, 13.

68. Duft, R.G.; Castro, A.; Bonfante, I.L.P.; Brunelli, D.T.; Chacon-Mikahil, M.P.T.; Cavaglieri, C.R. Metabolomics Approach in the Investigation of Metabolic Changes in Obese Men after 24 Weeks of Combined Training. J. Proteome Res. 2017, 16, 2151–2159, doi:10.1021/acs.jproteome.6b00967.

69. Kolahdouzi, S.; Baghadam, M.; Kani-Golzar, F.A.; Saeidi, A.; Jabbour, G.; Ayadi, A.; De Sousa, M.; Zouita, A.; Abderrahmane, A.B.; Zouhal, H. Progressive Circuit Resistance Training Improves Inflammatory Biomarkers and Insulin Resistance in Obese Men. Physiol. Behav. 2019, 205, 15–21, doi:10.1016/j.physbeh.2018.11.033.

70. Ho, S.S.; Dhaliwal, S.S.; Hills, A.P.; Pal, S. The Effect of 12 Weeks of Aerobic, Resistance or Combination Exercise Training on Cardiovascular Risk Factors in the Overweight and Obese in a Randomized Trial. BMC Public Health 2012, 12, 704, doi:10.1186/1471-2458-12-704.

71. Heydari, M.; Freund, J.; Boutcher, S.H. The Effect of High-Intensity Intermittent Exercise on Body Composition of Overweight Young Males. J. Obes. 2012, 2012, 480467, doi:10.1155/2012/480467.

72. Boutcher, Y.N.; Boutcher, S.H.; Yoo, H.Y.; Meerkin, J.D. The Effect of Sprint Interval Training on Body Composition of Postmenopausal Women. Med. Sci. Sports Exerc. 2019, 51, 1413–1419, doi:10.1249/MSS.0000000000001919.

73. Ahmadizad, S.; Avansar, A.S.; Ebrahim, K.; Avandi, M.; Ghasemikaram, M. The Effects of Short-Term High-Intensity Interval Training vs. Moderate-Intensity Continuous Training on Plasma Levels of Nesfatin-1 and Inflammatory Markers. Horm. Mol. Biol. Clin. Investig. 2015, 21, 165–173, doi:10.1515/hmbci-2014-0038.

74. Zhang, T.; Chen, S.; Saito, A. A META-ANALYSIS OF THE EFFECTS OF GREEN TEA COMBINED WITH PHYSICAL ACTIVITY ON BLOOD LIPIDS IN HUMANS. Rev. Bras. Med. Esporte 2020, 26, 454–460, doi:10.1590/1517-869220202605212295.

75. Asbaghi, O.; Fouladvand, F.; Gonzalez, M.J.; Aghamohammadi, V.; Choghakhori, R.; Abbasnezhad, A. Effect of Green Tea on Anthropometric Indices and Body Composition in Patients with Type 2 Diabetes Mellitus: A Systematic Review and Meta-Analysis. Complement. Med. Res. 2021, 28, 244–251, doi:10.1159/000511665.

76. Kapoor, M.P.; Sugita, M.; Fukuzawa, Y.; Okubo, T. Physiological Effects of Epigallocatechin-3-Gallate (EGCG) on Energy Expenditure for Prospective Fat Oxidation in Humans: A Systematic Review and Meta-Analysis. J. Nutr. Biochem. 2017, 43, 1–10, doi:10.1016/j.jnutbio.2016.10.013.

77. Lin, Y.; Shi, D.; Su, B.; Wei, J.; Gaman, M.-A.; Sedanur Macit, M.; Borges do Nascimento, I.J.; Guimaraes, N.S. The Effect of Green Tea Supplementation on Obesity: A Systematic Review and Dose-Response Meta-Analysis of Randomized Controlled Trials. Phytother. Res. 2020, 34, 2459–2470, doi:10.1002/ptr.6697.

78. Gomez-Ambrosi, J.; Silva, C.; Galofre, J.C.; Escalada, J.; Santos, S.; Millan, D.; Vila, N.; Ibanez, P.; Gil, M.J.; Valenti, V.; et al. Body Mass Index Classification Misses Subjects with Increased Cardiometabolic Risk Factors Related to Elevated Adiposity. Int. J. Obes. 2012, 36, 286–294, doi:10.1038/ijo.2011.100.

79. Lee, W.; Lee, D.; Han, E.; Choi, J. Intake of Green Tea Products and Obesity in Nondiabetic Overweight and Obese Females: A Systematic Review and Meta-Analysis. J. Funct. Foods 2019, 58, 330–337, doi:10.1016/j.jff.2019.05.010.

80. Ross, R.; Neeland, I.J.; Yamashita, S.; Shai, I.; Seidell, J.; Magni, P.; Santos, R.D.; Arsenault, B.; Cuevas, A.; Hu, F.B.; et al. Waist Circumference as a Vital Sign in Clinical Practice: A Consensus Statement from the IAS and ICCR Working Group on Visceral Obesity. Nat. Rev. Endocrinol. 2020, 16, 177–189, doi:10.1038/s41574-019-0310-7.

81. Covarrubias, A.J.; Horng, T. IL-6 Strikes a Balance in Metabolic Inflammation. Cell Metab. 2014, 19, 898–899, doi:10.1016/j.cmet.2014.05.009.

82. Gondoin, A.; Grussu, D.; Stewart, D.; McDougall, G.J. White and Green Tea Polyphenols Inhibit Pancreatic Lipase in Vitro. Food Res. Int. 2010, 43, 1537–1544, doi:10.1016/j.foodres.2010.04.029.

83. Vekic, J.; Zeljkovic, A.; Stefanovic, A.; Jelic-Ivanovic, Z.; Spasojevic-Kalimanovska, V. Obesity and Dyslipidemia. Metab.-Clin. Exp. 2019, 92, 71–81, doi:10.1016/j.metabol.2018.11.005.

84. Kuriyama, S. The Relation between Green Tea Consumption and Cardiovascular Disease as Evidenced by Epidemiological Studies. J. Nutr. 2008, 138, 1548S–1553S.

85. Murray, M.; Walchuk, C.; Suh, M.; Jones, P.J. Green Tea Catechins and Cardiovascular Disease Risk Factors: Should a Health Claim Be Made by the United States Food and Drug Administration? Trends Food Sci. Technol. 2015, 41, 188–197, doi:10.1016/j.tifs.2014.10.004.

86. Pirillo, A.; Casula, M.; Olmastroni, E.; Norata, G.D.; Catapano, A.L. Global Epidemiology of Dyslipidaemias. Nat. Rev. Cardiol. 2021, 18, 689–700, doi:10.1038/s41569-021-00541-4.

87. Xu, R.; Yang, K.; Li, S.; Dai, M.; Chen, G. Effect of Green Tea Consumption on Blood Lipids: A Systematic Review and Meta-Analysis of Randomized Controlled Trials. Nutr. J. 2020, 19, 48, doi:10.1186/s12937-020-00557-5.

88. Ference, B.A.; Ginsberg, H.N.; Graham, I.; Ray, K.K.; Packard, C.J.; Bruckert, E.; Hegele, R.A.; Krauss, R.M.; Raal, F.J.; Schunkert, H.; et al. Low-Density Lipoproteins Cause Atherosclerotic Cardiovascular Disease. 1. Evidence from Genetic, Epidemiologic, and Clinical Studies. A Consensus Statement from the European Atherosclerosis Society Consensus Panel. Eur. Heart J. 2017, 38, 2459–2472, doi:10.1093/eurheartj/ehx144.

89. Chen, R.; Lai, X.; Xiang, L.; Li, Q.; Sun, L.; Lai, Z.; Li, Z.; Zhang, W.; Wen, S.; Cao, J.; et al. Aged Green Tea Reduces High-Fat Diet-Induced Fat Accumulation and Inflammation via Activating the AMP-Activated Protein Kinase Signaling Pathway. Food Nutr. Res. 2022, 66, 7923, doi:10.29219/fnr.v66.7923.

90. Wonchung, L. Study on the role of AMPK on the obesity by inflammation. Asian J. Phys. Educ. Sport Sci. 2020, 8, 187–196, doi:10.24007/ajpess.2020.8.3.016.

91. Akhter, F.; Rahman, M.S.; Amin, G.M.A.; Miah, M.I.; Koh, Y.S. Beneficial Therapy with Natural Anti-Inflammatory Agents and Supplements. J. Bacteriol. Virol. 2021, 51, 149–162.

92. Khalafi, M.; Symonds, M.E. The Impact of High Intensity Interval Training on Liver Fat Content in Overweight or Obese Adults: A Meta-Analysis. Physiol. Behav. 2021, 236, 113416, doi:10.1016/j.physbeh.2021.113416.

