## supplementary tables for "Effects of Exercise Combined With Green Tea in Non-Aging on Anthropometric and Blood Lipids in Non-Aging Overweight/Obesity Adults-A Systematic Review and Network Meta-Analysis"

**Figure 1.** The full process of literature screening.

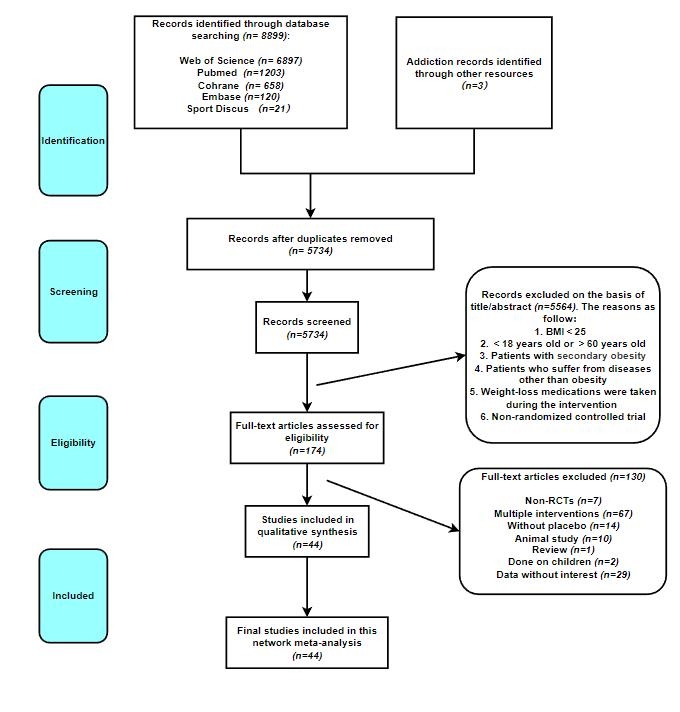

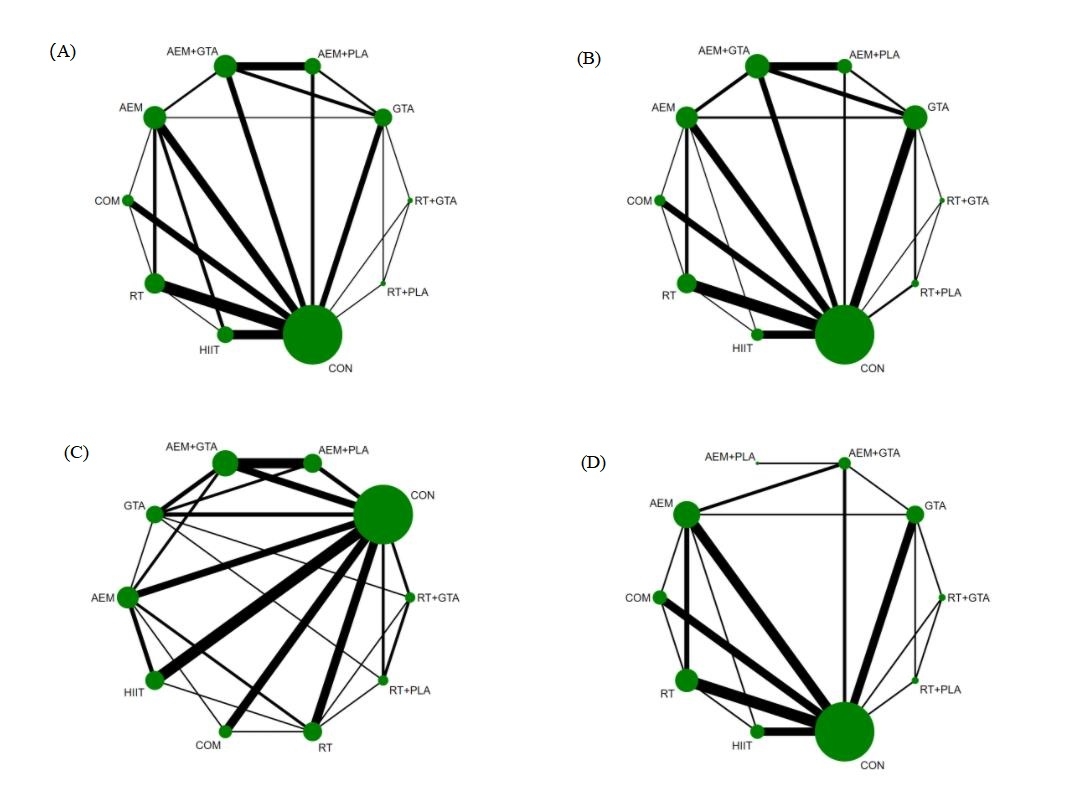

**Figure 2.** The network map of anthropometric indicators. (A): BW; (B): BMI; (C): BF%; (D): WC. AEM：aerobic exercise; RT: resistance training; HIIT: high-intensity interval training; GTA: green tea or green tea extract; AEM+GTA: aerobic training combined with green tea; RT+GTA: resistance training combined with green tea; AEM+PLA: aerobic training combined with placebo; RT+PLA: resistance training combined with placebo; COM: aerobic training combined with resistance training; CON: control group

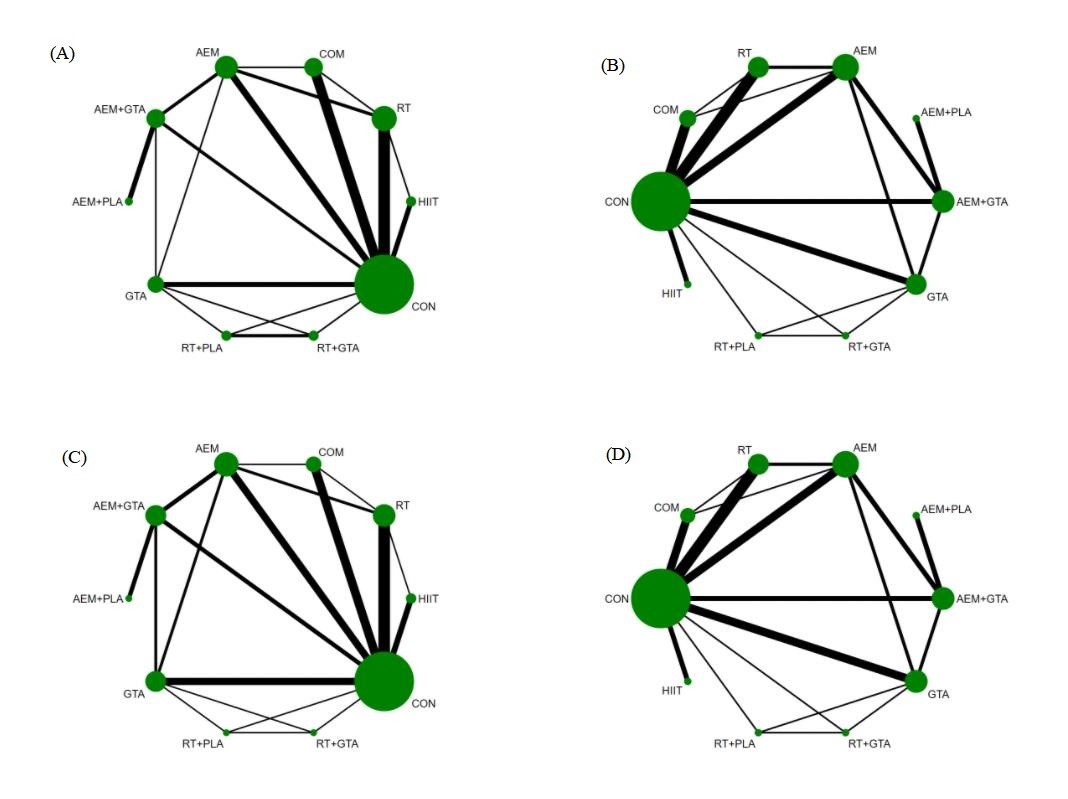

**Figure 3.** The network map of lipid profile indicators. (A): TG; (B): TC; (C): HDL; (D): LDL. AEM：aerobic exercise; RT: resistance training; HIIT: high-intensity interval training; GTA: green tea or green tea extract; AEM+GTA: aerobic training combined with green tea; RT+GTA: resistance training combined with green tea; AEM+PLA: aerobic training combined with placebo; RT+PLA: resistance training combined with placebo; COM: aerobic training combined with resistance training; CON: control group

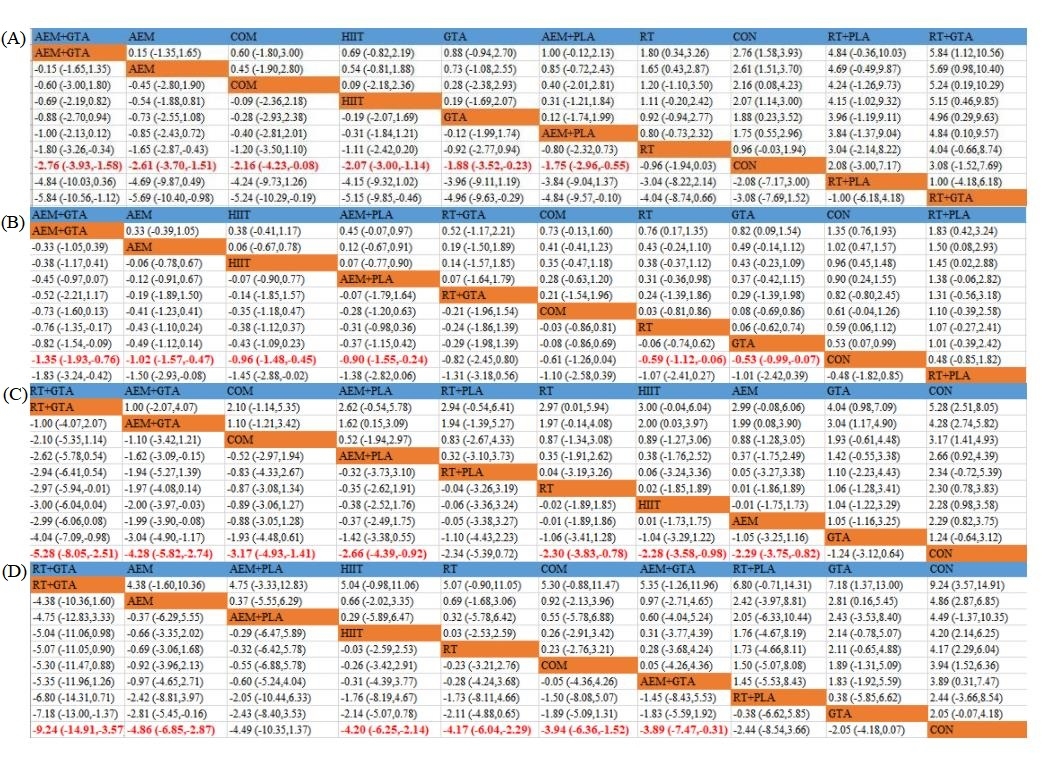

**Figure 4.** The network meta-analysis matrix of anthropometric indicators. (A): BW; (B): BMI; (C): BF%; (D): WC.

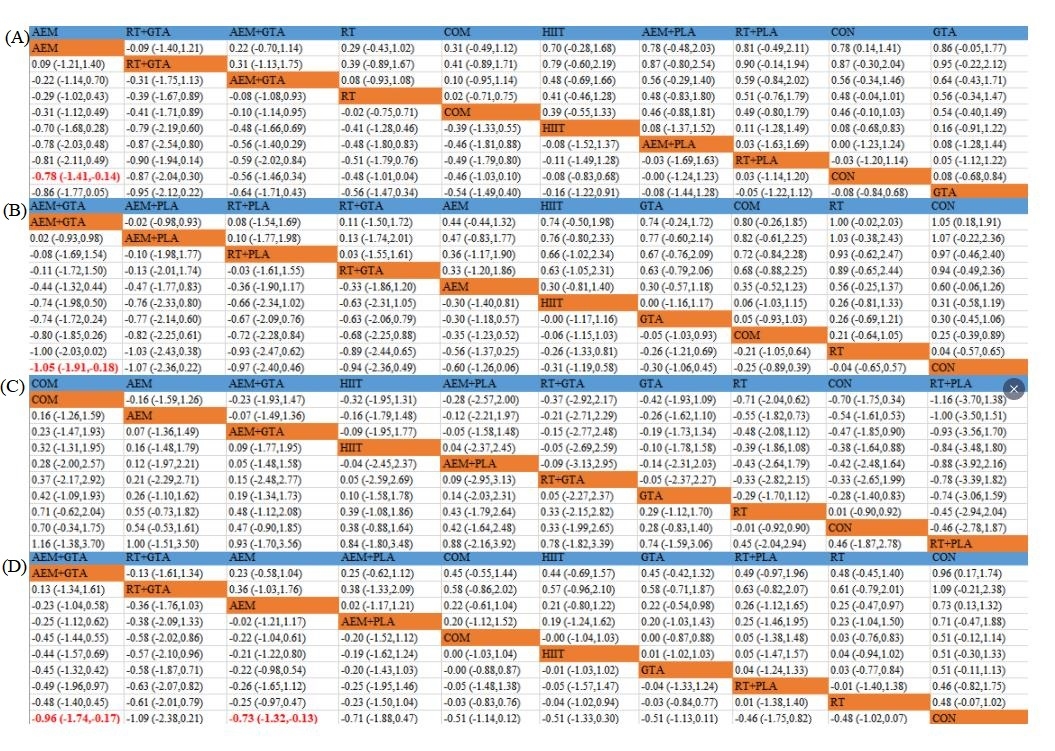

**Figure 5.** The network meta-analysis matrix of lipid profile indicators. (A): TG; (B): TC; (C): HDL; (D): LDL.
