## supplementary figures for "Effects of Exercise Combined With Green Tea in Non-Aging on Anthropometric and Blood Lipids in Non-Aging Overweight/Obesity Adults-A Systematic Review and Network Meta-Analysis"

**Supplementary Materials**

**Supplementary Table 1. Search strategy**

**Supplementary Table 2.** Specific information between studies.

**Supplementary Table 3**. Risk of bias.

**Supplementary table 4.** Inconsistency test

**Supplementary Figure 1.** Transferability check.

**Supplementary Figure 2-9.** SUCRA for outcomes.

**Supplementary Figure 10-17. Funnel plot for outcomes.**

**Supplementary Figure 18-25. Network forest figure for outcomes.**

**Supplementary Figure 26-33. SUCRA figure for outcomes.**

**PRISMA check list**

**Systematic Review Registration**

**Data**

**Supplementary Table 1. Search strategy**

| **Database** |  |
| --- | --- |
| **Pubmed** | (“green tea” [MeSH Terms] OR “tea” [All Fields] OR “green tea extract” [All Fields] OR “EGCG” [All Fields] OR “epigallocatechin gallate” [All Fields] OR “catechins” [All Fields]) AND (“exercise” [MeSH Terms] OR “physical activity” [All Fields] OR “exercise, physical” [All Fields] OR “isometric exercise” [All Fields] OR “aerobic exercise” [All Fields] OR “exercise training” [All Fields] OR “training“[All Fields]” OR “resistance training” [All Fields]) AND (“obesity” [MeSH Terms] OR “obese” [All Fields]OR “overweight” [All Fields] ) |
| **WOS** | **TOPIC: (**green tea OR tea OR green tea extract OR EGCG OR epigallocatechin gallate OR catechins) AND TOPIC: (exercise OR physical activity OR exercise, physical OR isometric exercise OR aerobic exercise OR exercise training OR training OR resistance training) AND TOPIC: (obesity OR obese OR overweight) Timespan: All years. Indexes: ALL |
| **Cochrane** | (green tea OR tea OR green tea extract OR EGCG OR epigallocatechin gallate OR catechins) in All Text AND (exercise OR physical activity OR exercise, physical OR isometric exercise OR aerobic exercise OR exercise training OR training OR resistance training) in All Text AND (obesity OR obese OR overweight) in All Text |

**Supplementary Table 2.** Specific information between studies.

| Study  (Author) | Year | The groups included in each original study (interventions, sample size, gender, average age, BMI) | | Whether to take supplements | Whether to do extra exercise | Duration | Outcomes |
| --- | --- | --- | --- | --- | --- | --- | --- |
| Touli | 2022 | GTA+AEM（n=10，M） | | 1000 mg/d GTA | AEM, 3d/week, 50%-70%HRR, 45min/d | 6 weeks | ②④⑤⑥⑦ |
|  |  | AEM (n=10, M) | | / | AEM, 3d/week, 50%-70%HRR, 45min/d |  |  |
|  |  | GTA (n=10, M) | | 1000 mg/d GTA | / |  |  |
|  |  | CON (n=10, M) | | / | / |  |  |
| Roberts | 2021 | GTA+AEM（n=9，F/M）: 46±5, BMI: 26.8±2.4 | | 1160 mg/d GTE (800mg/d EGCG) | AEM, 3-5/ week and 75%-80% HRmax，150min/week | 8 weeks | ①②③④⑤⑥⑦⑧ |
|  |  | CON+AEM (n=9, F/M): 35±7, BMI: 28.2±4.1 | | Cellulose |  |  |  |
| Alikhani | 2021 | GTA+AEM（n=12，F）: 25.4±3.5, BMI: 26.7±1.7 | | 1350 mg/d GTE | AEM, cycling（45-60min/d, 3d/week） | 8 weeks | ①②④ |
|  |  | CON+AEM(n=12, F): 24.8±3.7, BMI: 26.8±1.4 | | Maltodextrin | AEM, cycling（45-60min/d, 3d/week） |  |  |
|  |  | CON (n=10, F): 24.3±3.8, BMI: 26.4±1.4 | | / | / |  |  |
| Bagheri A | 2020 | GTA+AEM（n=10，F）: 39.5±4.17, BMI: 26.76±2.07 | | 500 mg/d GTE (225mg/d ) | AEM（40%-60% HRR，3d/week） | 8 weeks | ①②④ |
|  |  | CON+AEM(n=10, F): 37.6±1.71, BMI: 27.49±2.33 | | Chickpea flour | AEM（40%-60% HRR，3d/week） |  |  |
|  |  | CON (n=10, F): 38.0±3.12, BMI: 27.56±1.40 | | / | / |  |  |
| Bagheri B | 2020 | GTA+AEM（n=15，M）: 44.6±3, BMI: 27.3±0.4 | | 500 mg/d GTE (225mg/d ) | AEM（40%-60% HRR，3d/week） | 8 weeks | ①②④ |
|  |  | CON+AEM(n=15, M): 43.8±3.4, BMI: 27.2±0.5 | | Chickpea flour | AEM（40%-60% HRR，3d/week） |  |  |
|  |  | CON (n=15, M): 44±3.5, BMI: 27.8±0.3 | | / | / |  |  |
| Ghasemi | 2020 | GTA+AEM（n=10，F）: 22.47±3.32, BMI: 34.12±1.8 | | 1500 mg/d GTA (294 mg EGCG) | AEM，40 meters shuttle run（80% HRmax，3d/week） | 10 weeks | ①②④ |
|  |  | CON+AEM(n=10, F): 23.58±2.23, BMI: 27.32±1.27 | | Maltodextrin | AEM，40 meters shuttle run（80% HRmax，3d/week） |  |  |
|  |  | GTA（n=10，F）: 21.06±2.65, BMI: 34.28±1.38 | | 1500 mg/d GTA (294 mg EGCG) | / |  |  |
| Amozadeh | 2018 | GTA+AEM（n=13，F/M）: 28.14±7.48, BMI: 33.44±3.78 | | 100mg/d GTE | AEM（80-90min/d 3d/weeks, 70%-80% HRR） | 8 weeks | ①②③④⑤⑥⑦⑧ |
|  |  | AEM(n=13, F/M): 27.12±6.5, BMI: 32.77±5.42 | | / | AEM（80-90min/d 3d/weeks, 70%-80% HRR） |  |  |
|  |  | CON (n=13, F/M): 28.13±6.54, BMI: 33.87±4.25 | | / | / |  |  |
| Ghadami | 2018 | GTA+RT (n=15, M) | 34±8, BMI: 29.88±2.08 | 1500 mg/d GTA | RT（90min/d: 15min warming up+60min working group +10min resting， 3d/week） | 8 weeks | ②③⑤⑥⑦⑧ |
|  |  | RT (n=15, M) |  | Cellulose | RT（90min/d: 15min warming up+60min working group +10min resting， 3d/week） |  |  |
|  |  | GTA (n=15, M) |  | 1500 mg/d GTA | / |  |  |
|  |  | CON (n=15, M) |  | / | / |  |  |
| Gahreman | 2016 | GTA+AEM (n=12, M) | 26.1±0.7, BMI: ＞25 | 500 mg/d GTA (125mg EGCG) | AEM（5min warming up+20min  Working group 65%-80% HRR）3d/week | 12 weeks | ①②④⑤⑥⑦⑧ |
|  |  | AEM (n=12, M) |  | / | AEM（5min warming up+20min  Working group 65%-80% HRR）3d/week |  |  |
|  |  | GTA (n=12, M) |  | 500 mg/d GTA (125mg EGCG) | / |  |  |
|  |  | CON (n=12, M) |  | / | / |  |  |
| Marjan | 2017 | GTA+AEM（n=12，F） | 54±2.7  BMI: 28.8±3.2 | 1200 mg/d GTE (210mg/d EGCG) | AEM（40-50分钟，4/ week） | 2 weeks | ①②③⑤⑥⑦⑧ |
|  |  | CON+AEM（n=12，F） |  | Cellulose | AEM（40-50分钟，4/ week） |  |  |
| Cardoso | 2013 | GTA+RT (n=9, F) | 20-40, BMI: 25-35 | 1200 mg/d GTE (210mg/d EGCG) | RT（3d/week 8 group/d，60min/d） | 8 weeks | ①②③④⑤ |
|  |  | CON+RT (n=9, F) |  | Cellulose | RT（3d/week 8 group/d，60min/d） |  |  |
|  |  | GTA (n=9, F) |  | 1200 mg/d GTE (210mg/d EGCG) | / |  |  |
|  |  | CON (n=9, F) |  | / | / |  |  |
| Zhang | 2020 | GTA+AEM (n=12, M): 42.5±9.82, BMI: 28.4±2.24 | | 300mg/d GTA | AEM, Walking 60min/d, 4d/week 50%-65% HRmax | 12 weeks | ⑤⑥⑦⑧ |
|  |  | CON+AEM (n=12, M) 37.2±7.25, BMI: 27.7±2.26 | | Cellulose | AEM, Walking 60min/d, 4d/week 50%-65% HRmax |  |  |
| Nabi | 2018 | GTA (n=41, F/M): 46.2±6.4, BMI: 29.95 ± 1.79 | | 300mg/d GTA | / | 12 weeks | ①②③⑤⑥⑦⑧ |
|  |  | CON (n=43, F/M): 47.68 ± 7.58, BMI: 29.69 ± 2.01 | | Maltodextrin | / |  |  |
| Kamesh | 2018 | GTA (n=20, F/M) | 35-45, BMI＞25 | 780 mg/d GTA | / | 12 weeks | ①②③④⑤⑥⑦⑧ |
|  |  | CON (n=20, F/M) |  | Cellulose | / |  |  |
| Afzalpour | 2017 | GTA+AEM (n=10, F): 20.37±1.5, BMI: 27.15±1.47 | | 1500 mg/d GTA | AEM（4-8times/ week） | 10 weeks | ①②③ |
|  |  | CON±AEM (n=10, F): 21.00±1.82, BMI: 27.32±1.27 | | Cellulose | AEM（4-8times/ week） |  |  |
|  |  | GTA (n=10, F): 21.85±1.34, BMI: 28.03±1.04 | | 1500 mg/d GTA | / |  |  |
| Redhwan | 2013 | GTA (n=15, F) | 20-25, BMI＞25 | 1500 mg/d GTA | / | 4 weeks | ①②④ |
|  |  | CON (n=15, F) |  | Cellulose | / |  |  |
| Suliburska | 2012 | GTA (n=23, F/M) | 40-60, BMI：27-31 | 500 mg/d GTA | / | 12 weeks | ②④⑦⑧ |
|  |  | CON (n=23, F/M) |  | Cellulose | / |  |  |
| Atashak | 2016 | COM (n=15, M): 42.0±3.3, BMI: 28.29±1.62 | | / | COM（3d/week,＜80%HRmax walking/running 20min and ＜80%1RM RT, 70min/d） | 8 weeks | ①②③④⑤⑥⑦⑧ |
|  |  | CON (n=15, M) 43.0±3.0, BMI: 27.75±1.67 | | / | / |  |  |
| Bonfante | 2017 | COM (n=12, M) | 49.13±5.75, BMI: 30.86±1.63 | / | COM（3d/week,55%- 80%VO2peak walking/running and ＜80%1RM RT, 60min/d） | 26 weeks | ①②③④⑤⑥⑦⑧ |
|  |  | CON (n=15, M) |  | / | / |  |  |
| Boutcher | 2019 | HIIT (n=20, F): 54.1±3.6, BMI: 28.3±3.7 | | / | AEM（3d/week 80%-85% HRmax, cycling） | 8 weeks | ①②③④⑥⑦⑧ |
|  |  | CON (n=20, F) 53.3±3.4, BMI: 27.3±4.1 | | / | / |  |  |
| Croymans | 2014 | RT (n=28, M) | 20-25, BMI:29-32 | / | RT（3d/week 8 group/d，60min/d） | 8 weeks | ①②③④⑤⑥⑦⑧ |
|  |  | CON (n=8, M) |  | / | / |  |  |
| Donges | 2010 | AEM (n=41, F/M) | 18-60, BMI: 26-30 | / | AEM（cycling，3d/week 70%-75% HRmax，30-50min） | 10 weeks | ①②③⑤⑥⑦⑧ |
|  |  | RT (n=35, F/M) |  | / | RT (7 groups/d, 30-50min/d, 3d/week) |  |  |
|  |  | CON (n=26, F/M) |  | / | / |  |  |
| Franklin | 2016 | RT (n=10, F): 30.3±5.4, BMI: 34.2±3 | |  | RT( 2/d week,80% 1RM,8-10 groups×2-3 cycle) | 8 weeks | ①②③④⑥⑦⑧ |
|  |  | CON (n=8, F) 30.8±9.0, BMI: 32.2±6.9 | | / | / |  |  |
| Ahmadizad A | 2014 | RT (n=8, M) | 23.4±0.6,BMI＞25 | / | RT (3d/week, 70%1RM,50min/d,8groups) | 8 weeks |  |
|  |  | CON (n=8, M) |  | / | / |  |  |
| Ahmadizad B | 2015 | AEM (n=10, M) | 25±1, BMI＞25 | / | AEM, walking (3d/week,50%-60%VO2max,30-70min） | 6 weeks | ①④ |
|  |  | HIIT (n=10, M) |  | / | HIIT (3d/week, 90%VO2max,30min） |  |  |
|  |  | CON (n=10, M) |  | / | / |  |  |
| Arad | 2015 | HIIT (n=11, F), 30±7 | | / | HIIT（4 groups at 75%-90%HRR and 3 groups at 50%HRR cross，24min） | 14 weeks | ①②④ |
|  |  | CON (n=9, F)，29±4 | | / | / |  |  |
| Christensen | 2019 | HIIT (n=14, F/M) | 30-50 | / | HIIT cycling（3d/week，8min 40%-60% VO2max warm up then 75%-85%VO2max,total 45min） | 12 weeks | ⑤⑦ |
|  |  | RT (n=12, F/M) |  | / | RT (60%-80%1RM,45min) |  |  |
|  |  | CON (n=13, F/M) |  | / | / |  |  |
| Duft | 2017 | COM (n=11, M) | 30-50 | / | COM (3d/week,30min RT and 50-85%VO2peak walking/running 30min) | 12 weeks | ①②③④ |
|  |  | CON (n=11, M) |  | / | / |  |  |
| Heydari | 2013 | HIIT (n=20, M) | 18-30 | / | HIIT(3d/week,80 %-90%HRmax, 20min) | 12 weeks | ①②③④⑤⑥⑦⑧ |
|  |  | CON (n=20, M) |  | / | / |  |  |
| Ho | 2013 | AEM (n=14, F/M) | 55±2 | / | AEM（5d/week, 60%HRR, 30min） | 8 weeks | ①②③④⑤⑥⑦⑧ |
|  |  | RT (n=17, F/M) |  | / | RT (5d/week, 75%1RM, 30min) |  |  |
|  |  | COM (n=15, F/M) |  | / | COM（5d/week, 60%HRR, 15min+75%1RM,15min） |  |  |
|  |  | CON (n=16, F/M) |  | / | / |  |  |
| Jang | 2019 | AEM (n=8, F) | 45-60 | / | AEM（4d/week 60%-70%VO2peak，50min） | 8 weeks | ①②③④ |
|  |  | RT (n=8, F) |  | / | RT（4d/week 60%-70%1RM，50min） |  |  |
|  |  | CON (n=8, F) |  | / | / |  |  |
| Keating | 2018 | RT (n=12, F/M) | 25-50,BMI＞25 | / | RT (3d/week，80%-85% 1RM, 45min) | 8 weeks | ①②④⑤⑥⑦⑧ |
|  |  | CON (n=14, F/M) |  | / | / |  |  |
| Kolahdouzi | 2019 | RT (n=13, M) | 23±3.2，BMI:30.67±3.06 | / | RT (3d/week，65%-85% 1RM, 35min) | 8 weeks | ①②⑤⑥⑦⑧ |
|  |  | CON (n=13, M) |  | / |  |  |  |
| Nikseresht | 2014 | HIIT (n=12, M): 38.9±4.1 | | / | HIIT（3d/week，80%-90%HRmax working group and 55%-65% HRmax resting group，35min，running） | 12 weeks | ①②③④ |
|  |  | RT (n=13, M)：39.6±3.7 | | / | RT (3d/week,45%-90%1RM,10 groups,35min) |  |  |
|  |  | CON (n=10, M)：40.4±5.2 | | / | / |  |  |
| Oh | 2018 | COM (n=8, F/M) | 32-40 | / | COM (3d/week, 70%1RM 40min+60%-85%HRmax running, walking,20min) | 8 weeks | ①②③④⑤⑥⑦ |
|  |  | CON (n=8, F/M) |  | / | / |  |  |
| Winn | 2019 | AEM (n=8): 46±9 | BMI:＞25 | / | AEM（4d/week，55%VO2peak，58min cycling） | 4 weeks | ①②③④ |
|  |  | HIIT （n=8）：41±14 |  | / | HIIT（4d/week，80%VO2peak working group+50% VO2peak resting group，56min） |  |  |
|  |  | CON（n=5）：51±13 |  | / | / |  |  |

| Ataeinosrat | 2022 | RT (n=11，M) | 27.5±9.4 | / | RT (3d/week 60%1RM, 13×3, 10 groups，70min) | 12 weeks | ①②③⑤⑥⑦⑧ |
| --- | --- | --- | --- | --- | --- | --- | --- |
|  |  | CON (n=11，M) |  | / | / |  |  |
| Sun | 2020 | HIIT (n=150, F/M)  21.78±1.47 | | / | HIIT (5d/week，85%VO2max，30min) | 12 weeks | ①②③④⑤⑥⑦⑧ |
|  |  | CON (n=150, F/M)  21.63±1.39 | | / | / |  |  |
| Streb | 2022 | COM (n=18, F/M) | 37±1，BMI: 33±0.4 | / | COM (3d/week, 30min 50%-60% HRmax walking+ 30min 10-12/group) | 16 weeks | ⑥⑦⑧ |
|  |  | CON (n=18, F/M) |  | / | / |  |  |
| Beer | 2022 | AEM (n=18, F/M) | 18-40,BMI:＞25 | / | AEM （3d/week，45min，60%VO2peak, cycling） | 12 weeks | ①②③⑦⑧ |
|  |  | HIIT (n=18, F/M) |  | / | HIIT (3d/week，35-45min) |  |  |
| Hu | 2022 | HIIT (n=17, F) | 18-25, BMI:25-30 | / | HIIT (5d/week, 30min,90%HRmax) | 4 weeks | ①②③ |
|  |  | CON (n=13, F) |  | / | / |  |  |
| Dupuit | 2022 | COM (n=8, F) | 45-60,BMI＞25 | / | COM （＜85% HRmax 20min+12 1RM 30min） | 12 weeks | ②③④⑤⑦⑥ |
|  |  | CON (n=9, F) |  | / | / |  |  |
| Said | 2021 | AEM (n=13, F/M) | 21.74±1.42，BMI: 36.21±2.43 | / | AEM (4d/week,50%-70%HRmax，60min) | 12 weeks | ①②④⑤⑥⑦⑧ |
|  |  | RT (n=16, F/M) |  | / | RT (4d/week,50%-55% 1RM，60min) |  |  |
|  |  | COM (n=13, F/M) |  | / | RT+AEM (4d/week，60min) |  |  |
|  |  | CON (n=13, F/M) |  | / | / |  |  |

Table notes：AEM：aerobic exercise; RT: resistance training; HIIT: high-intensity interval training; GTA: green tea or green tea extract; AEM+GTA: aerobic training combined with green tea; RT+GTA: resistance training combined with green tea; AEM+PLA: aerobic training combined with placebo; RT+PLA: resistance training combined with placebo; COM: aerobic training combined with resistance training; CON: control group; F:female; M:male; F/M: both male and female; BMI: body mass index; HRmax: maximum heart rate; HRR: heart rate reserve; VO_2max_: maximum oxygen uptake; VO_2peak_: peak oxygen uptake; d:day; min: minutes; RM：repetition maximum; ①: body weight; ②: body mass index; ③：waist circumference; ④: body fat percentage; ⑤: total cholesterol; ⑥: triglycerides; ⑦: high-density lipoprotein; ⑧: low-density lipoprotein

**Supplementary Table 3**. Risk of bias.

| Study | Random sequence generation | Allocation concealment | Blinding of participants and personnel | Blinding of outcome assessment | Incomplete outcome data | Selective reporting | Other bias |
| --- | --- | --- | --- | --- | --- | --- | --- |
| Touli | U | L | H | L | H | U | L |
| Roberts | L | H | H | U | H | L | L |
| Alikhani | U | U | H | U | L | L | U |
| Bagheri A | U | L | H | U | L | H | L |
| Bagheri B | L | L | H | L | L | H | L |
| Ghasemi | U | L | H | L | H | H | L |
| Amozadeh | L | L | H | L | L | U | L |
| Ghadami | L | U | H | U | L | L | L |
| Gahreman | L | U | H | L | L | H | L |
| Marjan | L | U | H | U | L | L | L |
| Cardoso | L | U | H | L | L | U | L |
| Zhang | L | L | H | H | L | U | L |
| Nabi | U | U | L | U | L | U | U |
| Kamesh | L | L | L | L | L | U | L |
| Afzalpour | L | L | H | U | U | U | L |
| Redhwan | L | L | H | L | L | U | L |
| Suliburska | U | U | H | L | U | U | L |
| Atashak | L | L | H | U | H | U | L |
| Bonfante | L | L | H | U | U | U | L |
| Boutcher | L | L | H | L | H | U | L |
| Croymans | L | L | H | L | H | H | L |
| Donges | L | L | H | L | L | U | L |
| Franklin | L | L | H | L | H | U | L |
| Ahmadizad A | L | L | H | L | H | U | L |
| Ahmadizad B | L | L | H | U | L | U | L |
| Arad | L | H | H | U | L | U | L |
| Christensen | H | L | H | L | L | L | L |
| Duft | L | U | H | L | L | L | L |
| Heydari | L | L | H | L | L | U | L |
| Ho | L | U | H | L | L | U | L |
| Jang | L | L | H | U | L | U | L |
| Keating | L | L | H | L | L | U | L |
| Kolahdouzi | L | L | H | L | L | U | L |
| Nikseresht | L | U | H | L | L | H | L |
| Oh | L | L | H | H | L | U | L |
| Winn | L | L | H | H | L | U | L |
| Ataeinosrat | L | L | H | H | L | L | L |
| Sun | L | L | H | L | U | L | L |
| Streb | L | L | H | L | L | U | L |
| Beer | L | L | H | U | L | U | L |
| Hu | L | L | H | L | L | U | L |
| Dupuit | L | L | H | L | L | U | L |
| Said | L | U | H | L | L | H | L |
| Venkatakrishnan | L | L | H | H | L | U | L |

**Supplementary table 4.** Inconsistency test

| Outcomes | Chi^2^ | Prob > chi2 |
| --- | --- | --- |
| BW | 7.13 | 0.9890 |
| BMI | 10.46 | 0.9407 |
| BF% | 24.55 | 0.2192 |
| WC | 19.42 | 0.1107 |
| TC | 2.18 | 0.9883 |
| TG | 3.23 | 0.9936 |
| LDL | 2.65 | 0.9766 |
| HDL | 1.06 | 0.9999 |

**Supplementary Figure 1.** Transferability check.

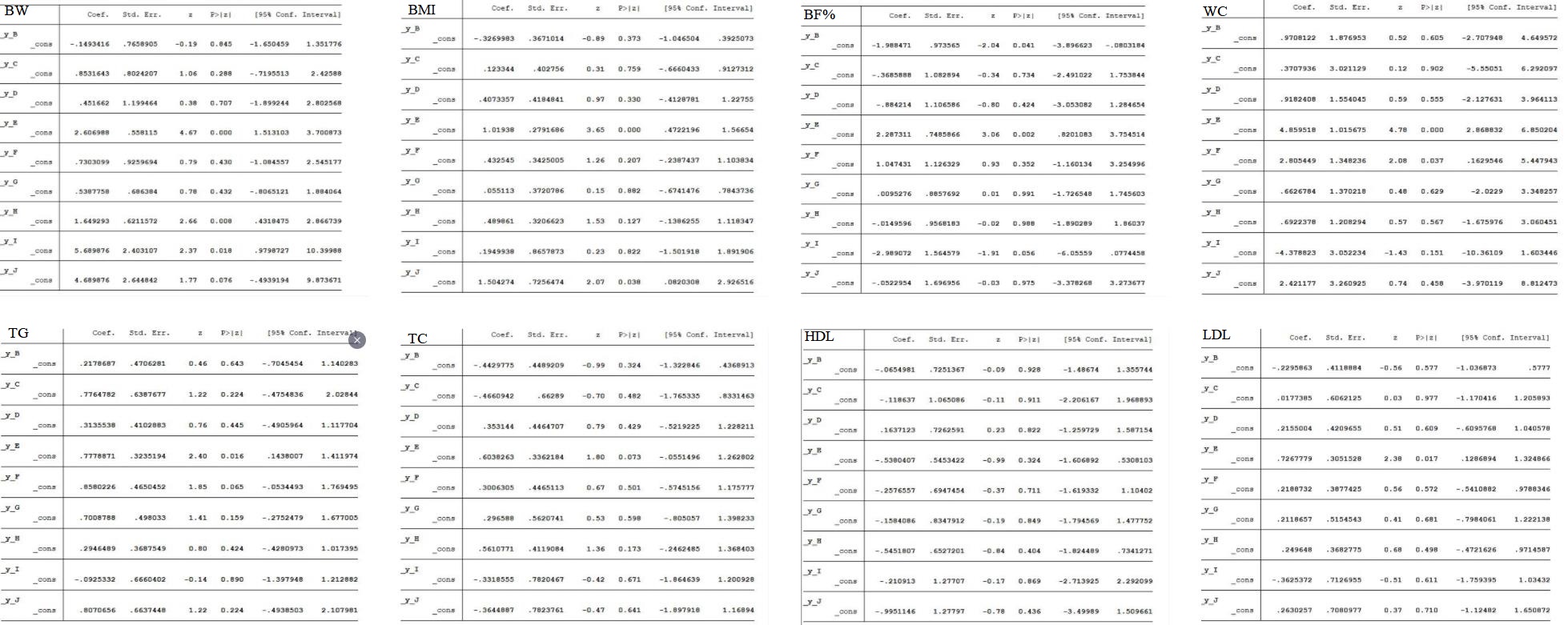

**Supplementary Figure 2. SUCRA for BW.**

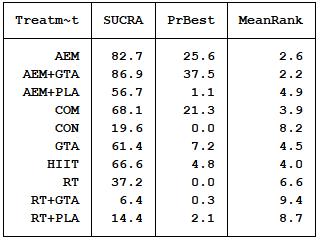

**Supplementary Figure 3. SUCRA for BMI.**

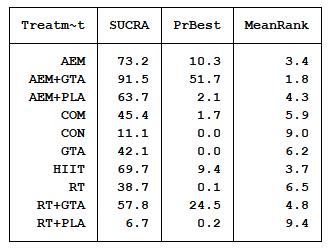

**Supplementary Figure 4. SUCRA for BF%.**

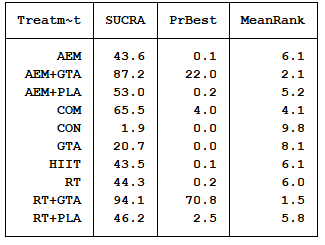

**Supplementary Figure 5. SUCRA for WC.**

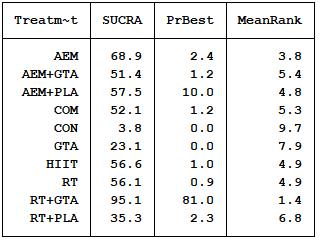

**Supplementary Figure 6. SUCRA for TG.**

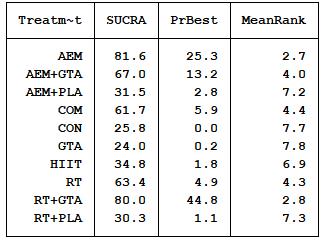

**Supplementary Figure 7. SUCRA for TC.**

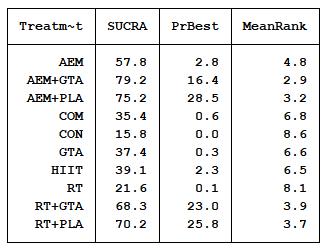

**Supplementary Figure 8. SUCRA for LDL.**

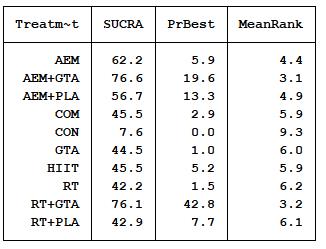

**Supplementary Figure 9. SUCRA for HDL.**

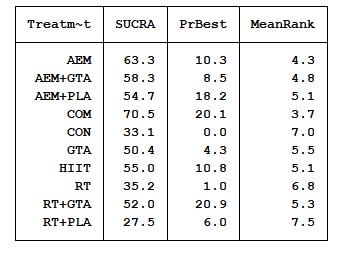

**Supplementary Figure 10. Funnel plot for BW.**

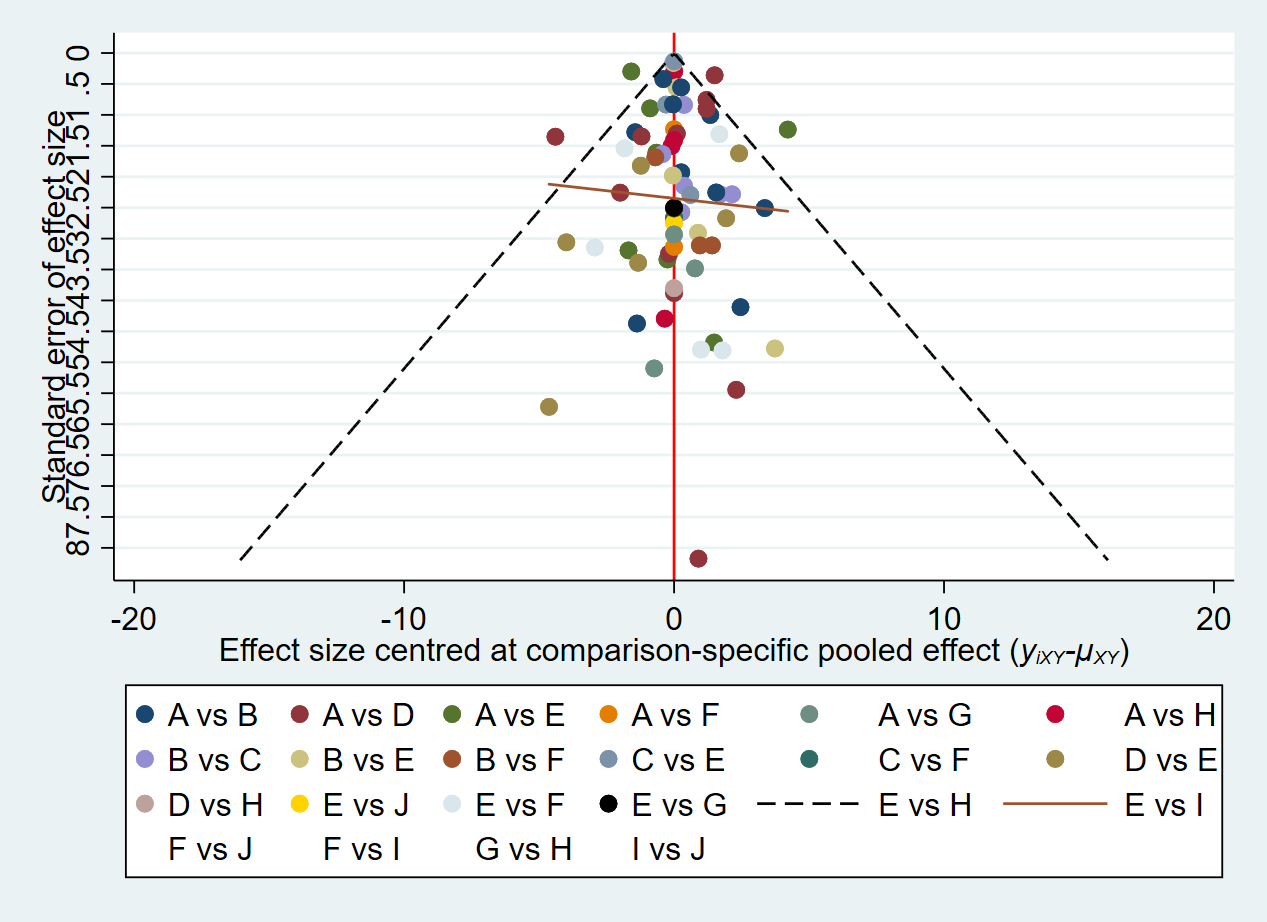

**Supplementary Figure 11. Funnel plot for BMI.**

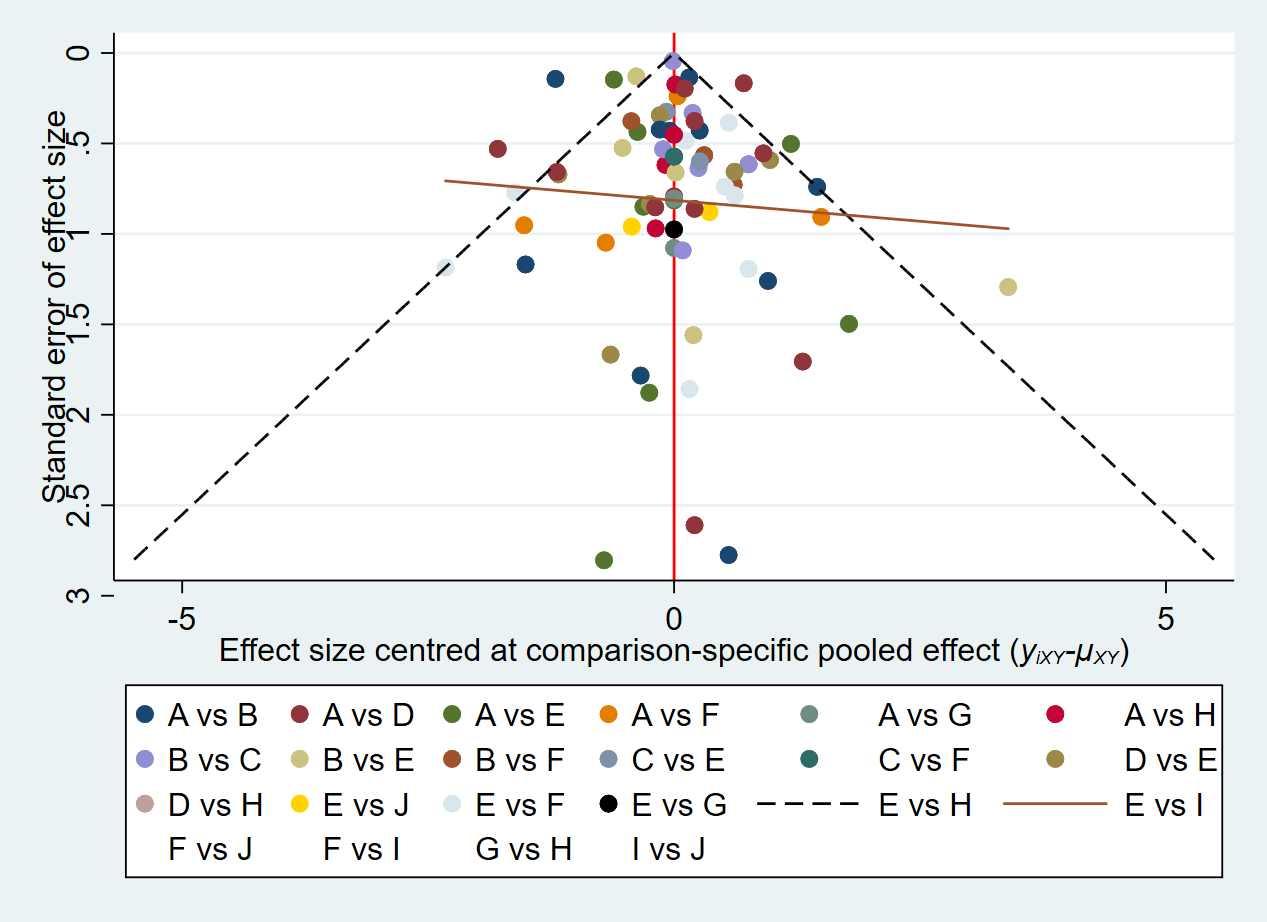

**Supplementary Figure 12. Funnel plot for BF%.**

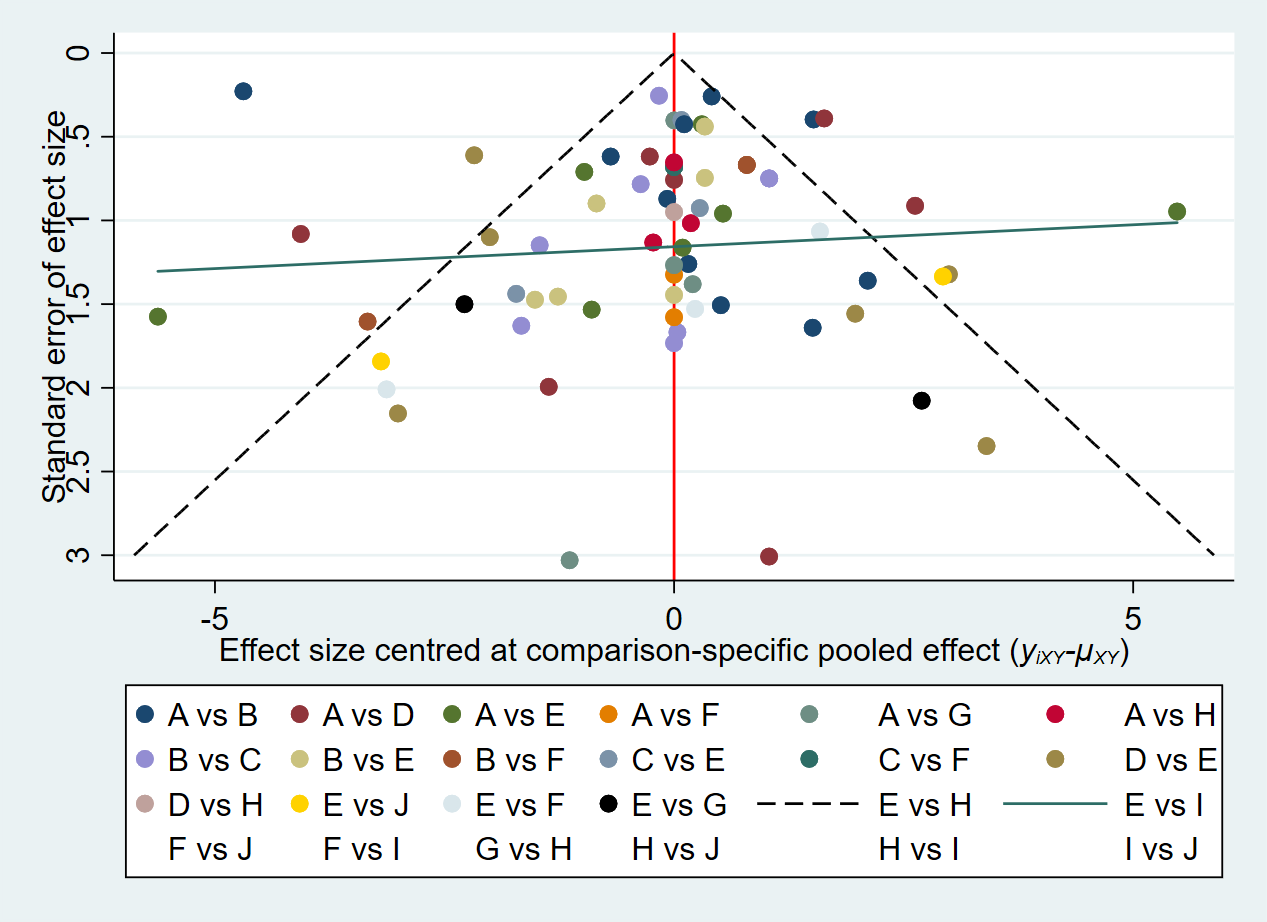

**Supplementary Figure 13. Funnel plot for WC.**

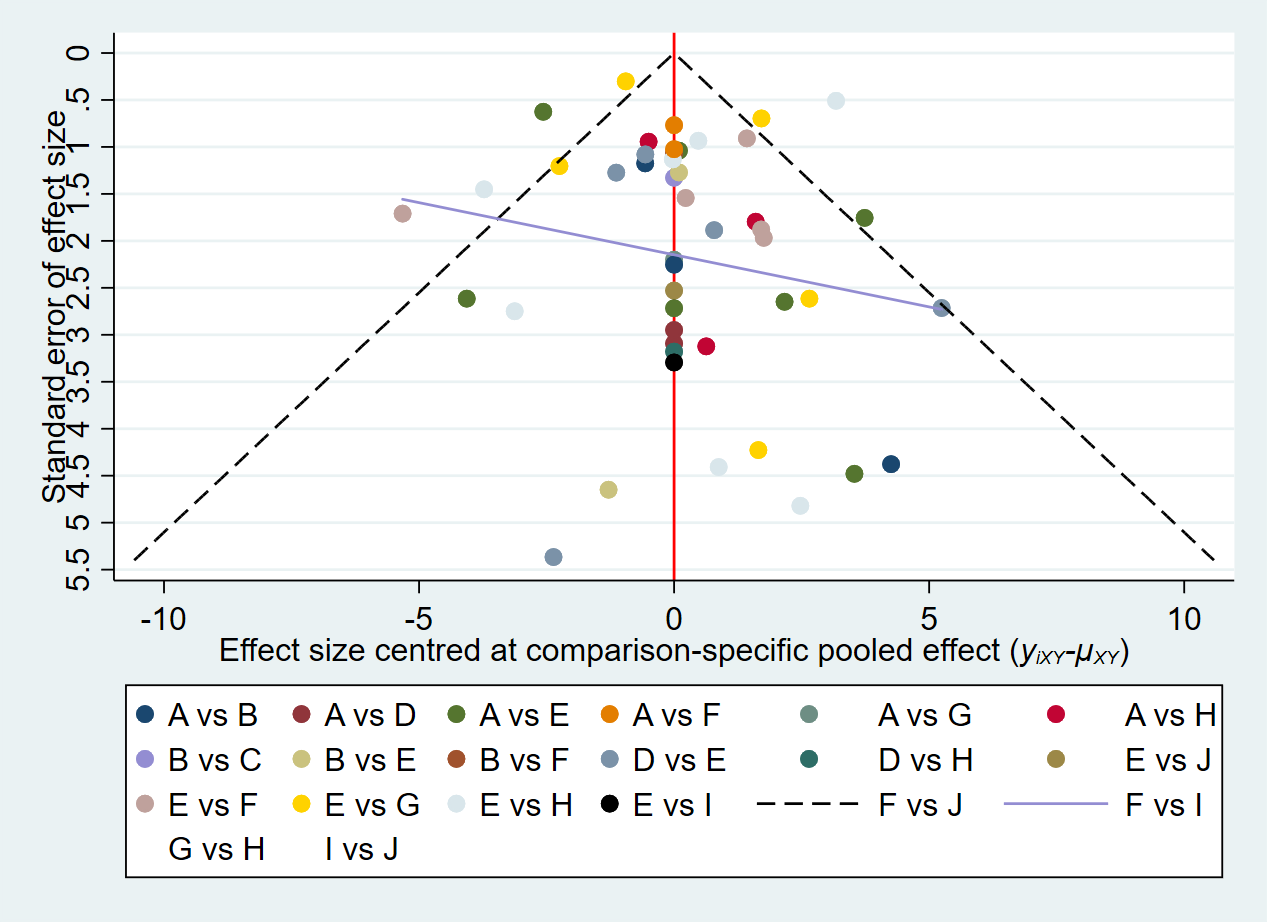

**Supplementary Figure 14. Funnel plot for TG.**

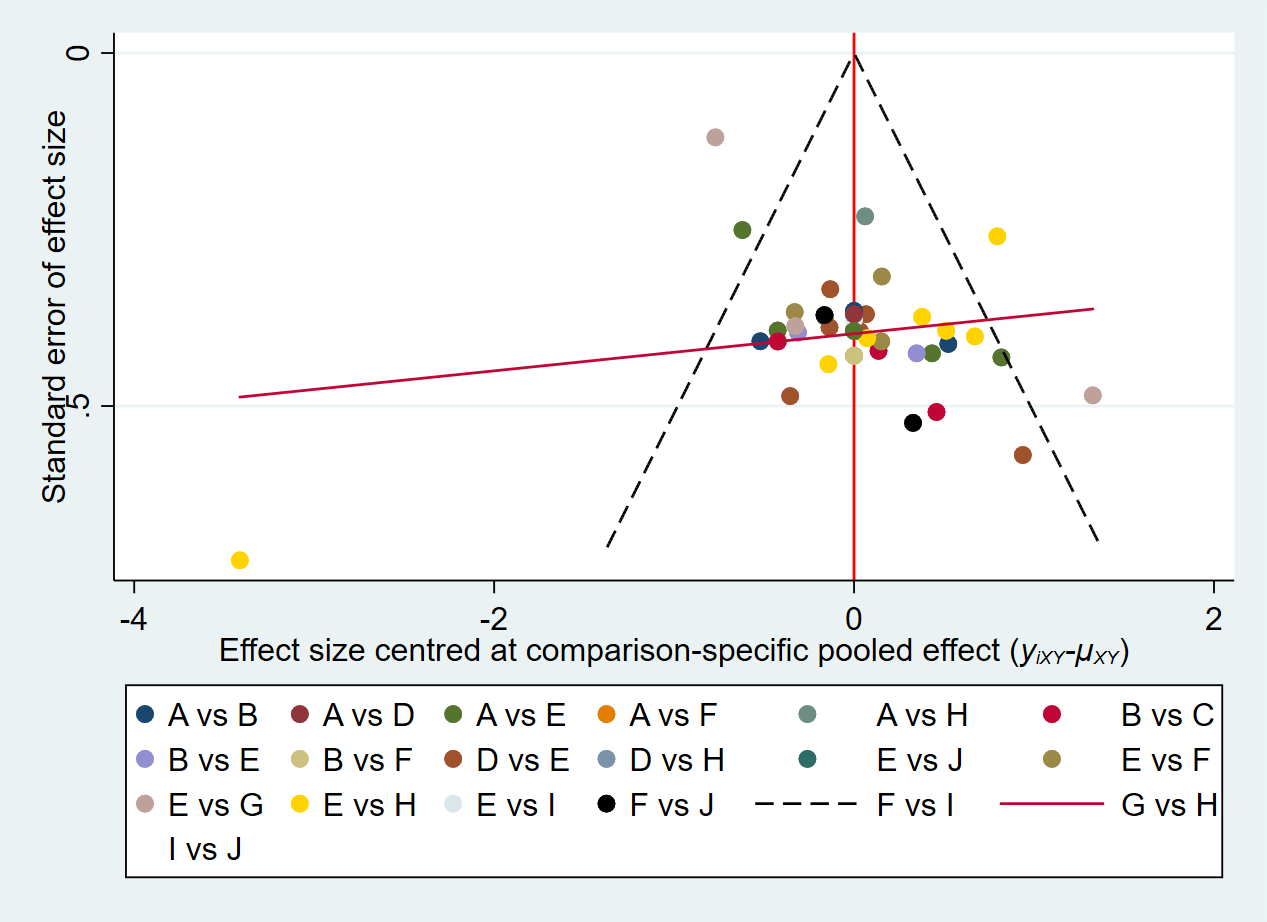

**Supplementary Figure 15. Funnel plot for TC.**

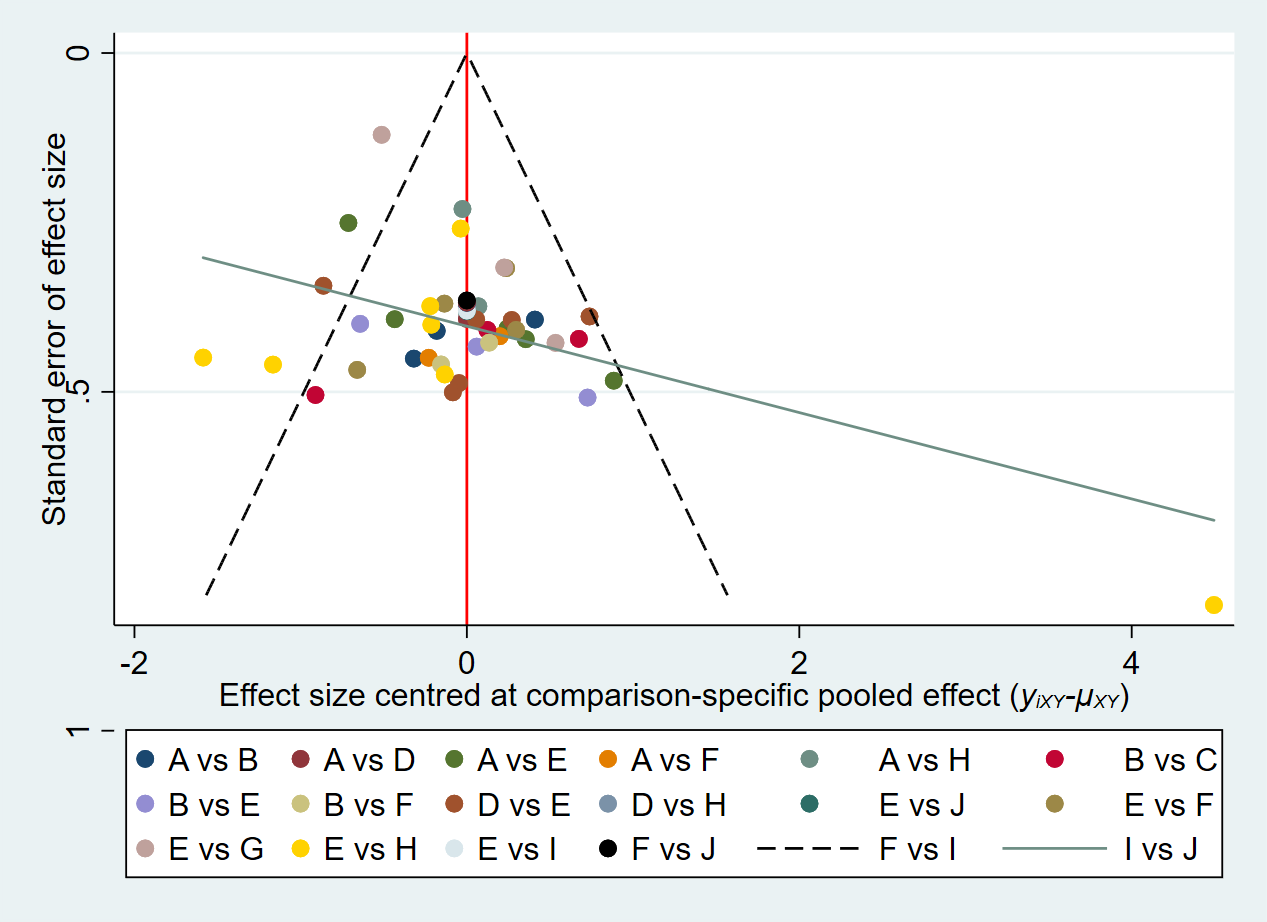

**Supplementary Figure 16. Funnel plot for LDL.**

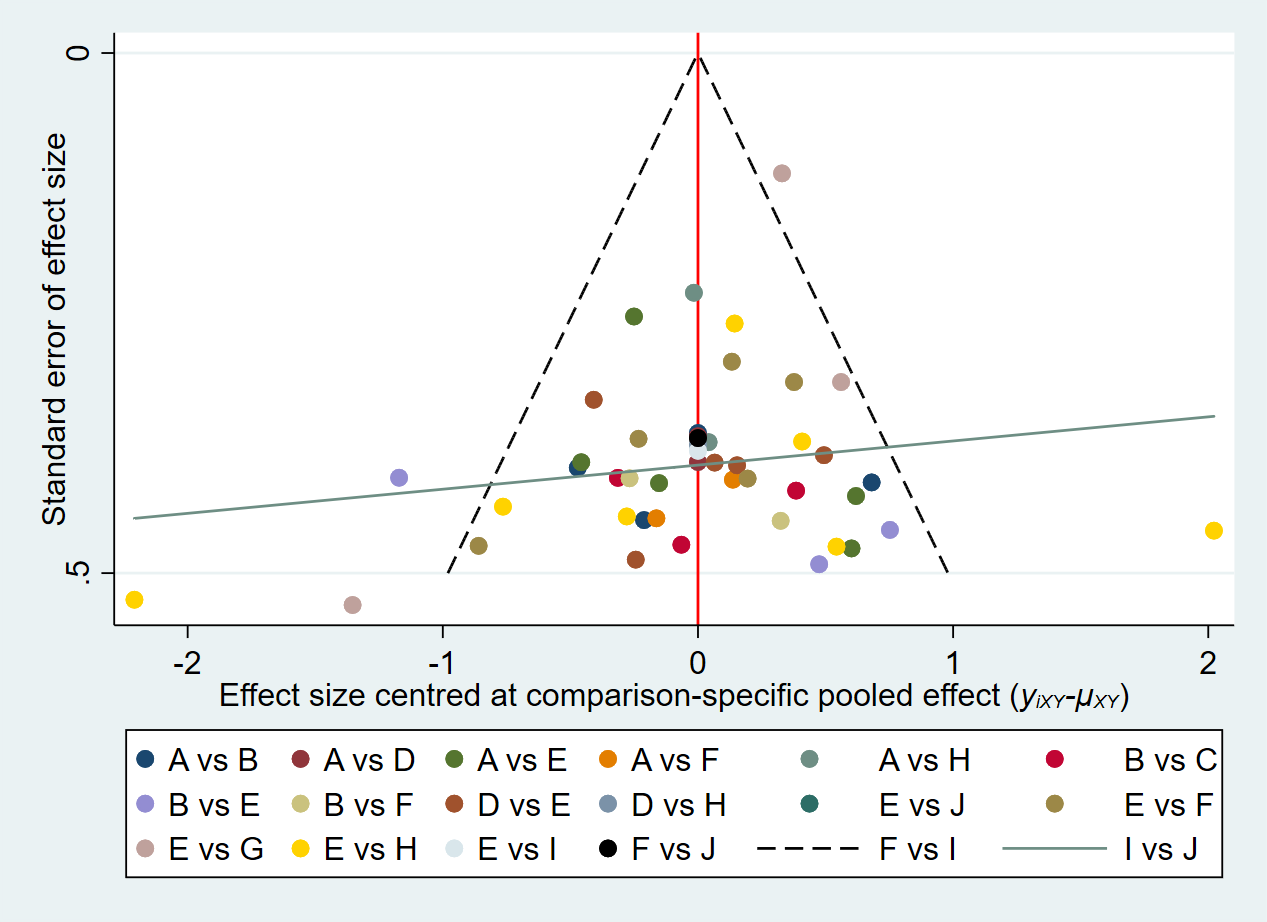

**Supplementary Figure 17. Funnel plot for HDL.**

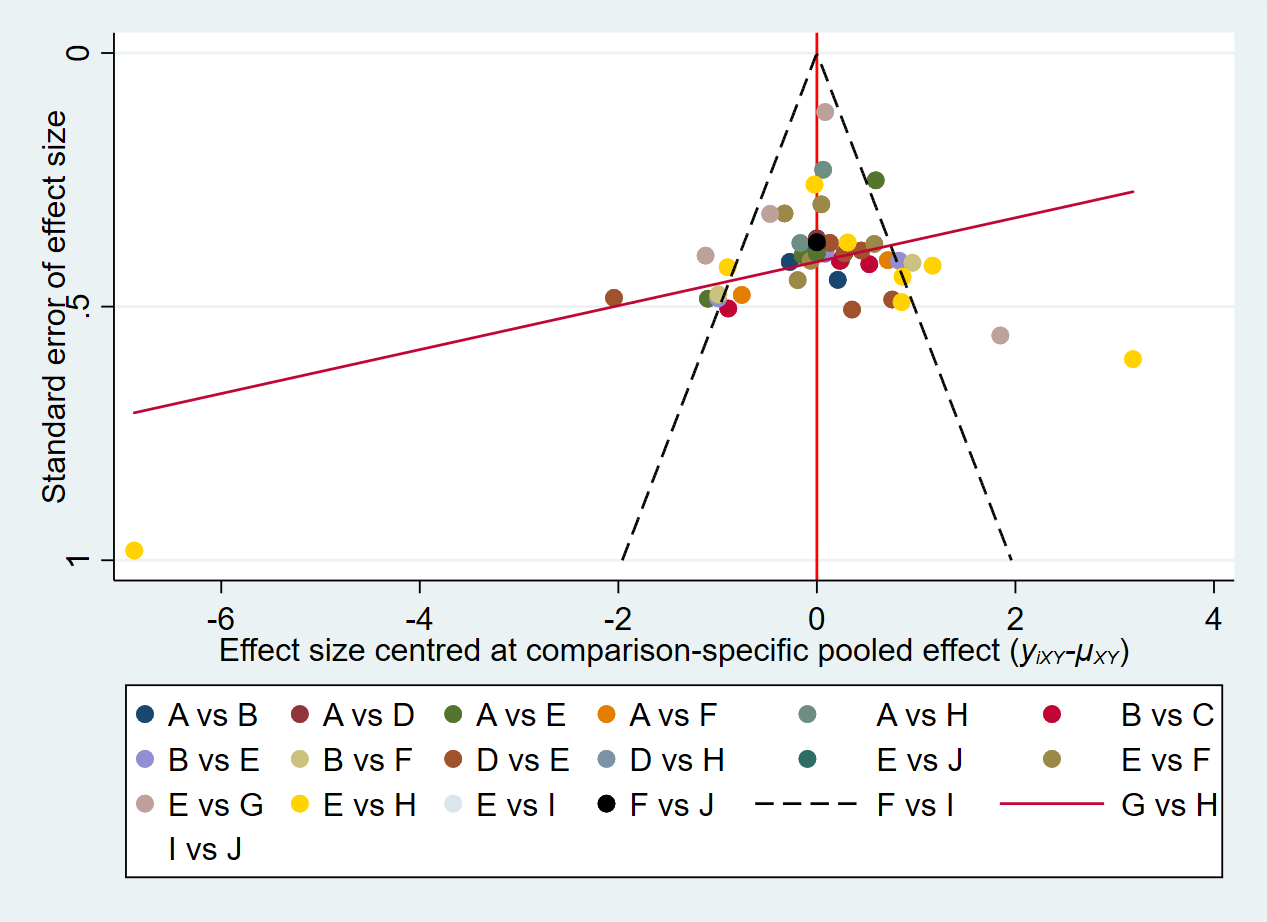

**Supplementary Figure 18. Network forest figure for BW.**

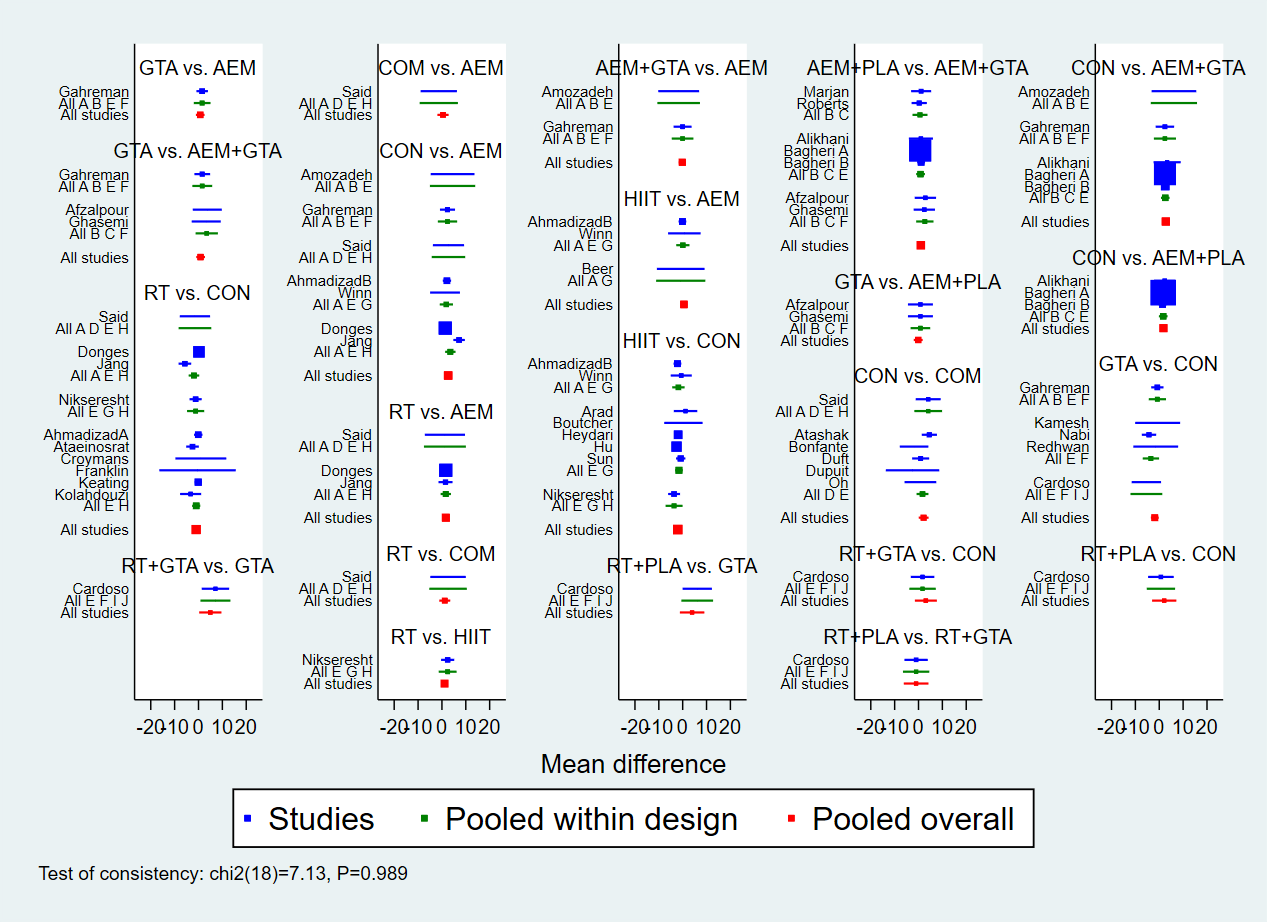

**Supplementary Figure 19. Network forest figure for BMI.**

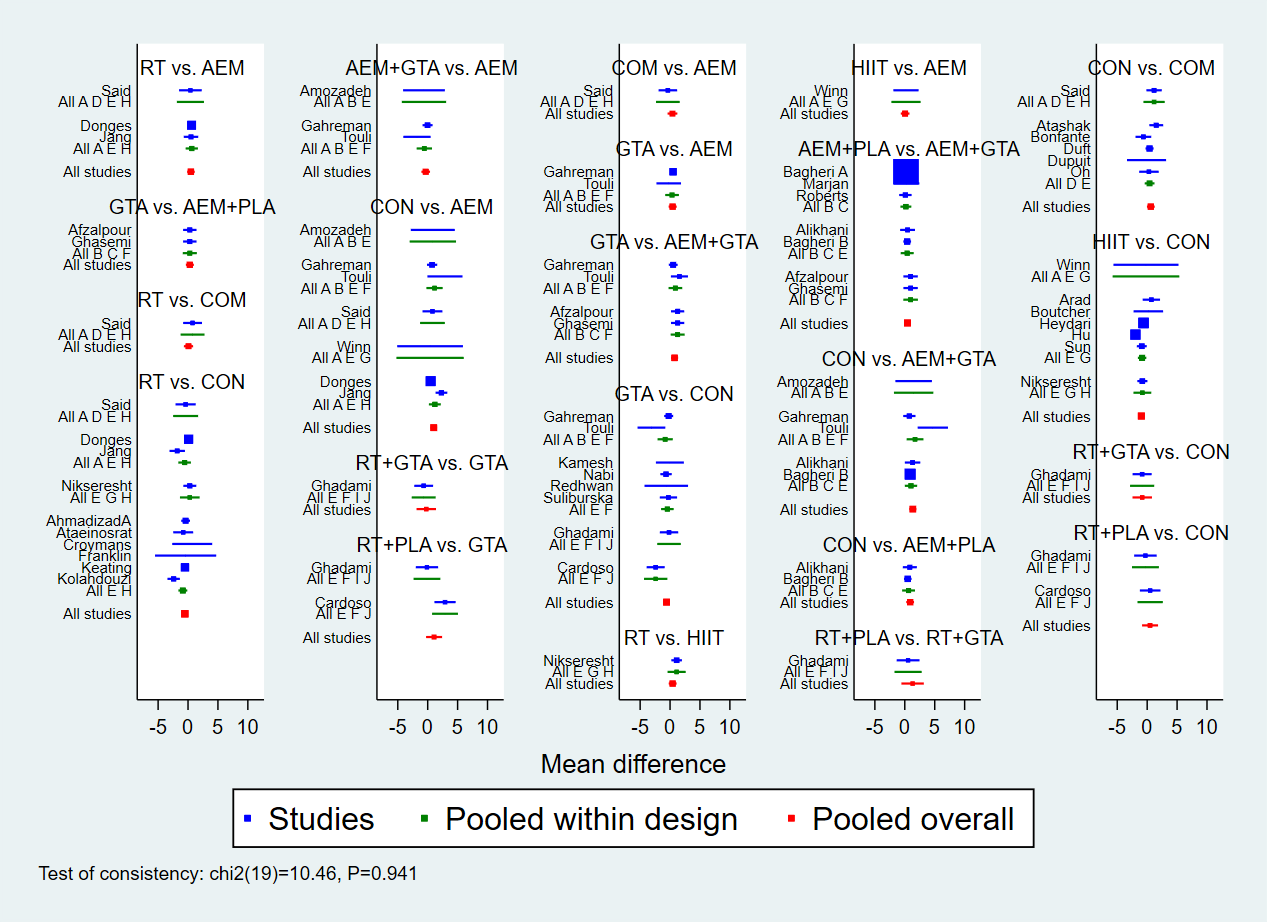

**Supplementary Figure 20. Network forest figure for BF%.**

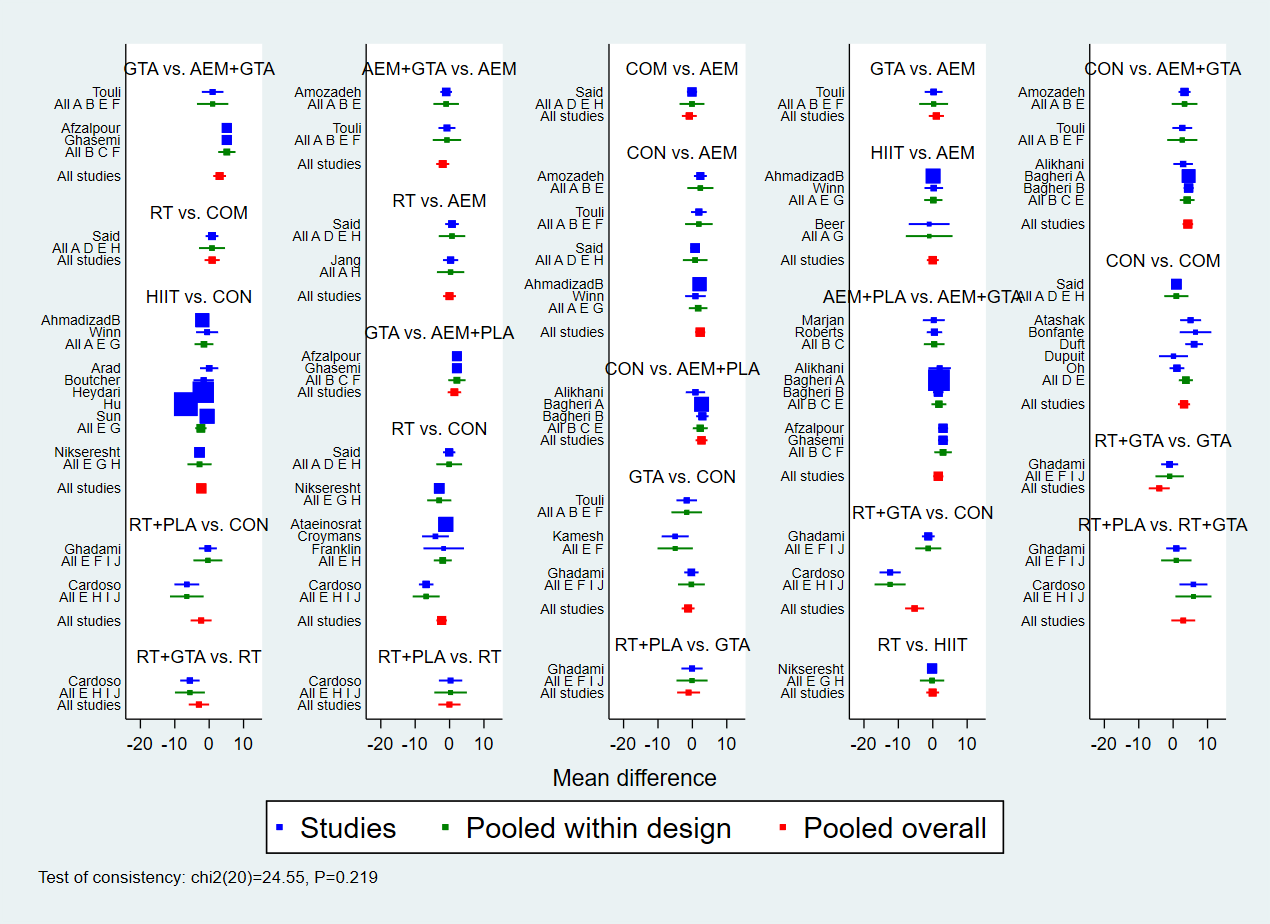

**Supplementary Figure 21. Network forest figure for WC.**

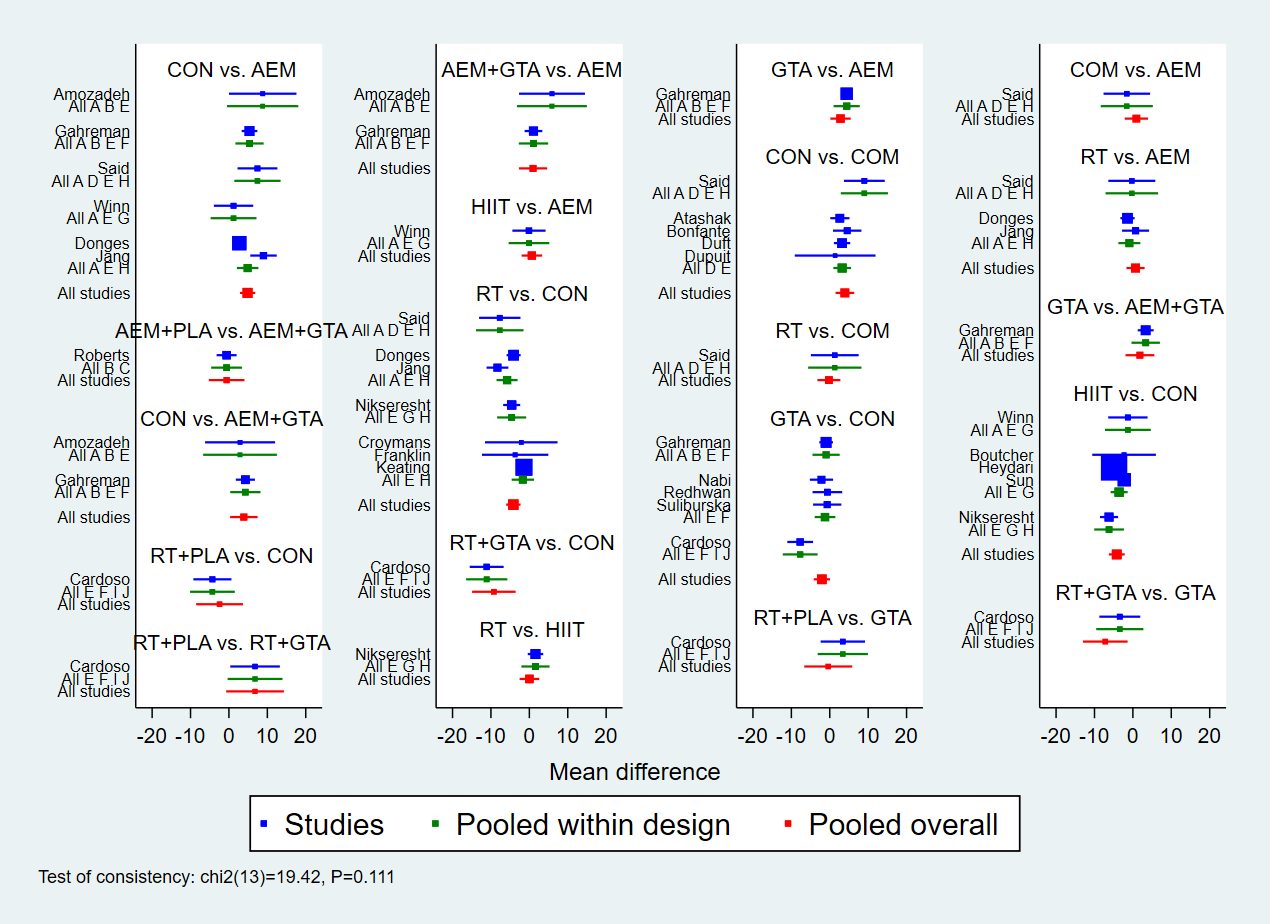

**Supplementary Figure 22. Network forest figure for TG.**

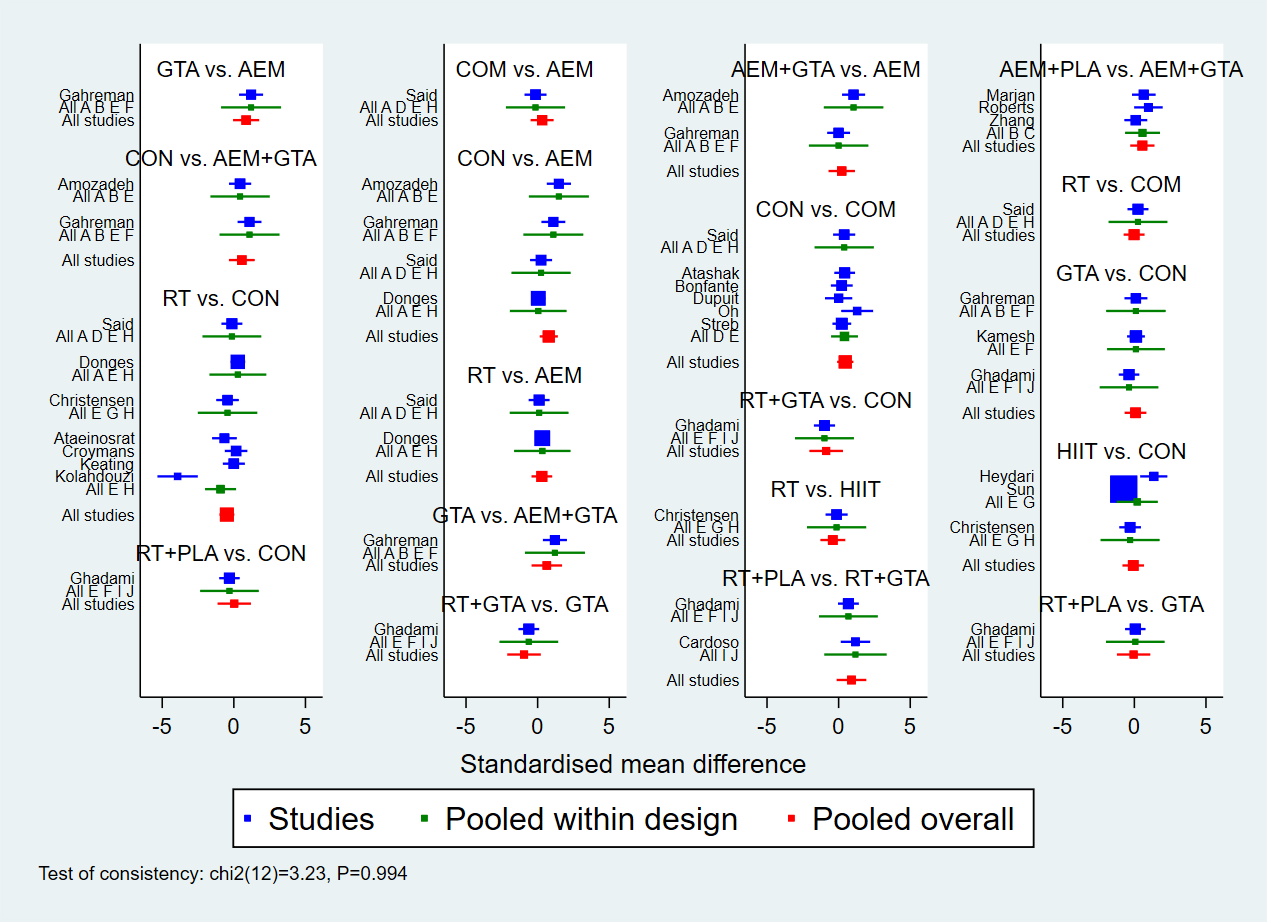

**Supplementary Figure 23. Network forest figure for TC.**

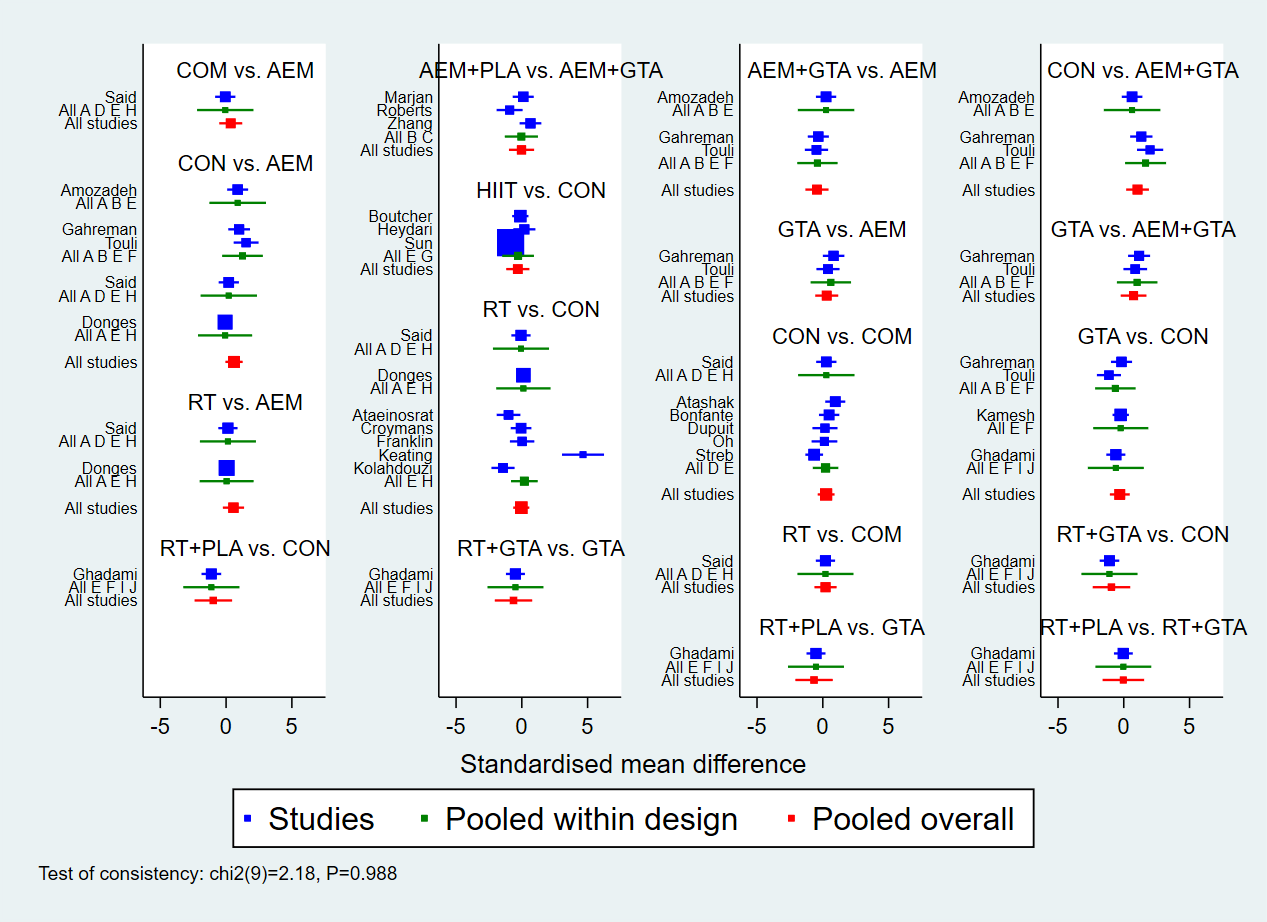

**Supplementary Figure 24. Network forest figure for LDL.**

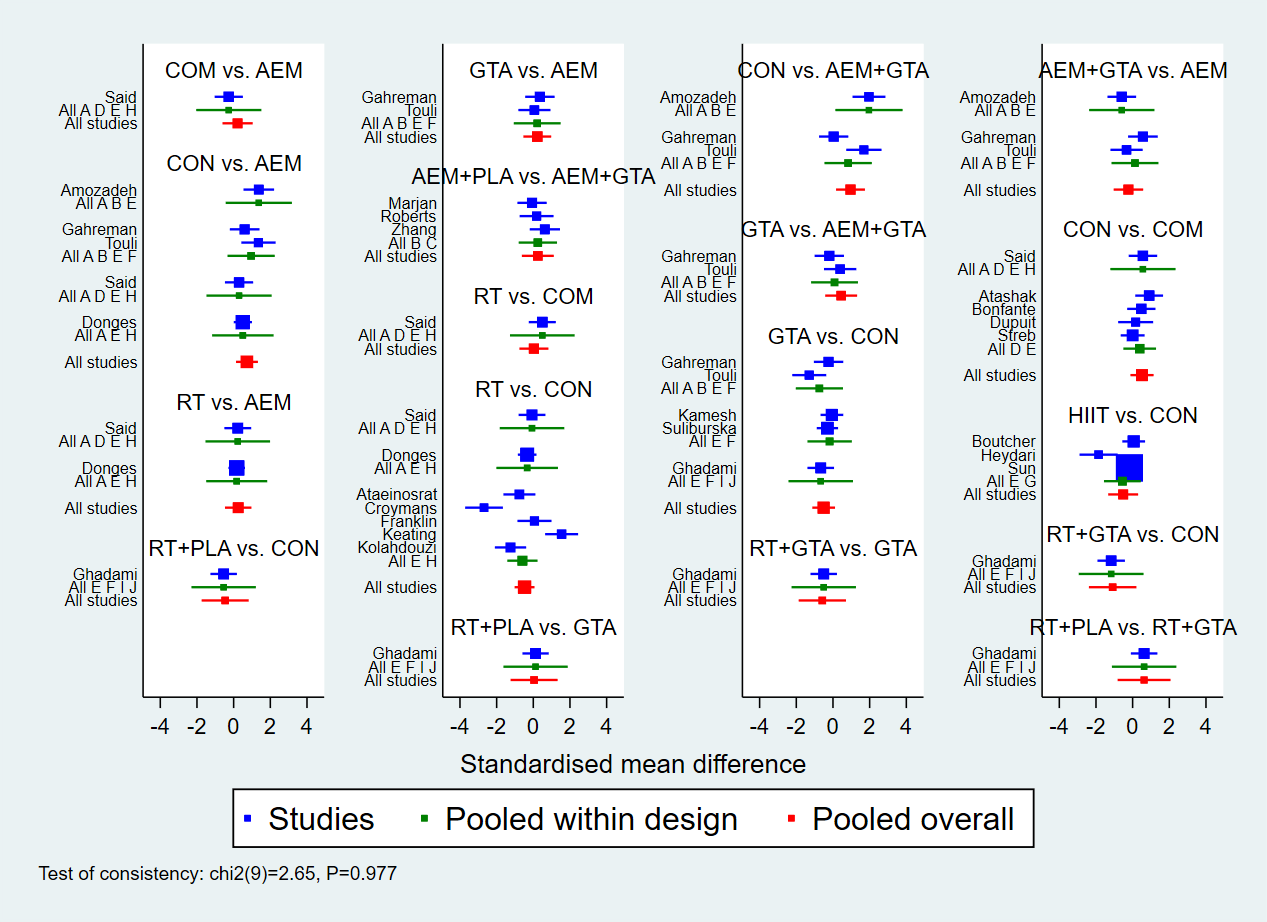

**Supplementary Figure 25. Network forest figure for HDL.**

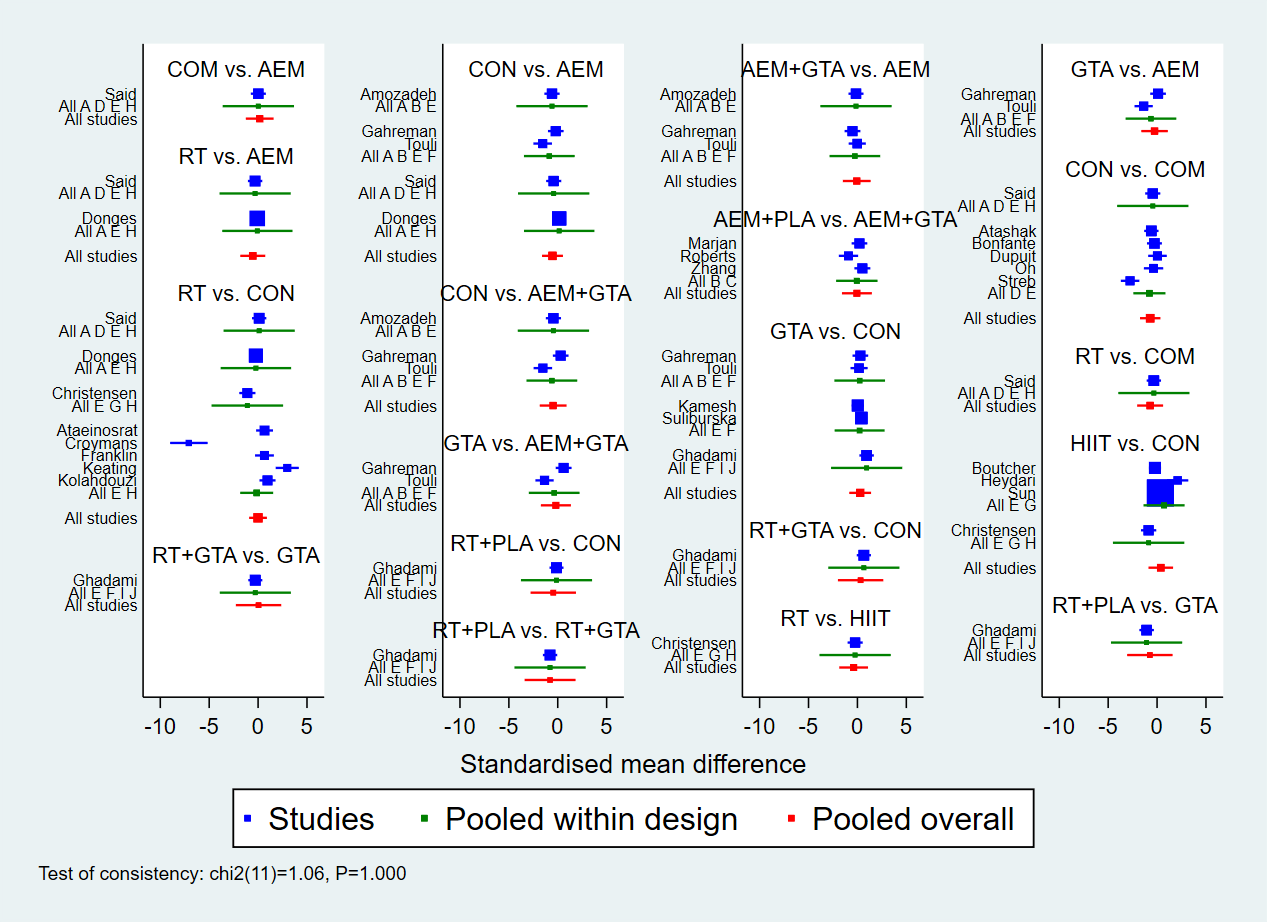

**Supplementary Figure 26. SUCRA figure for BW.**

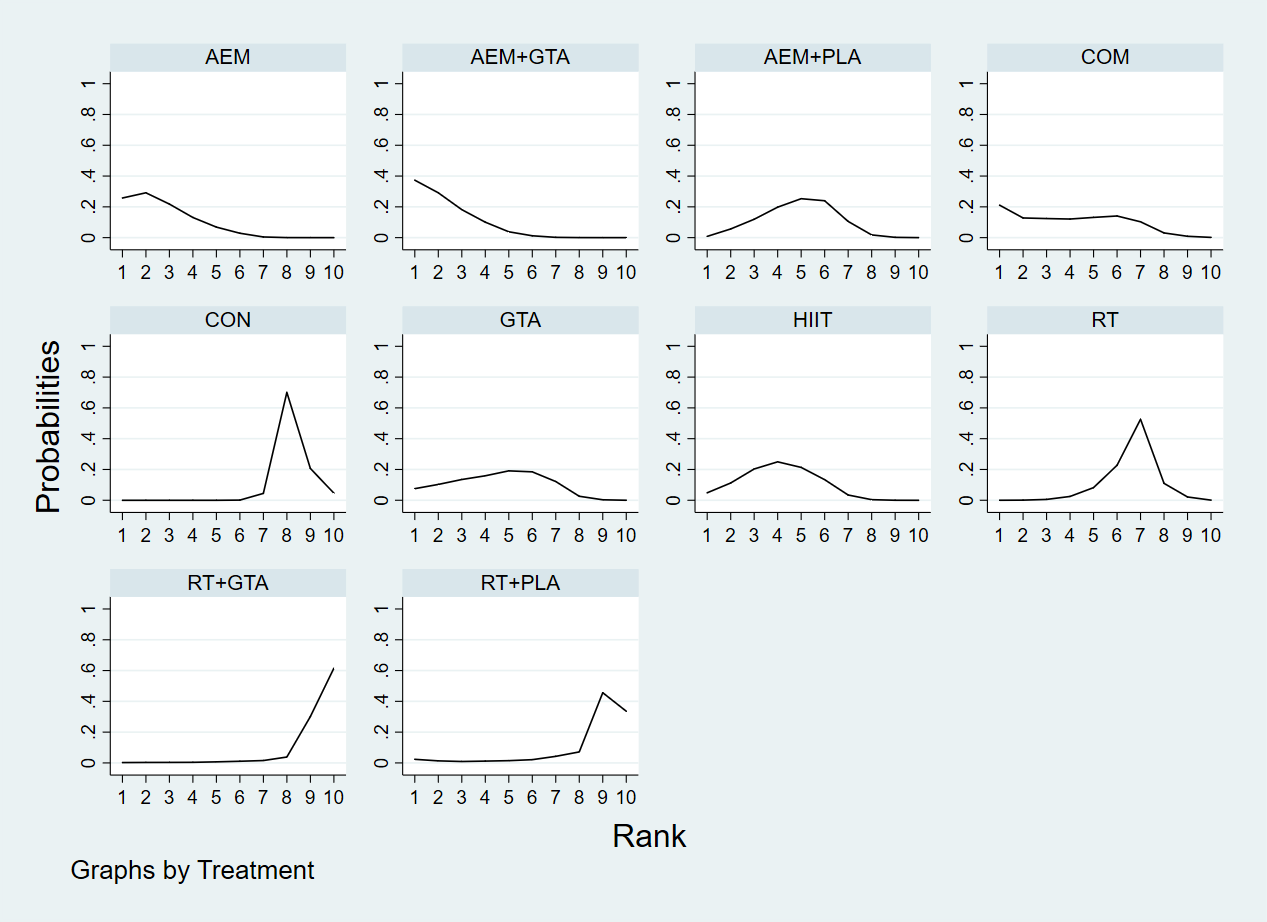

**Supplementary Figure 27. SUCRA figure for BMI.**

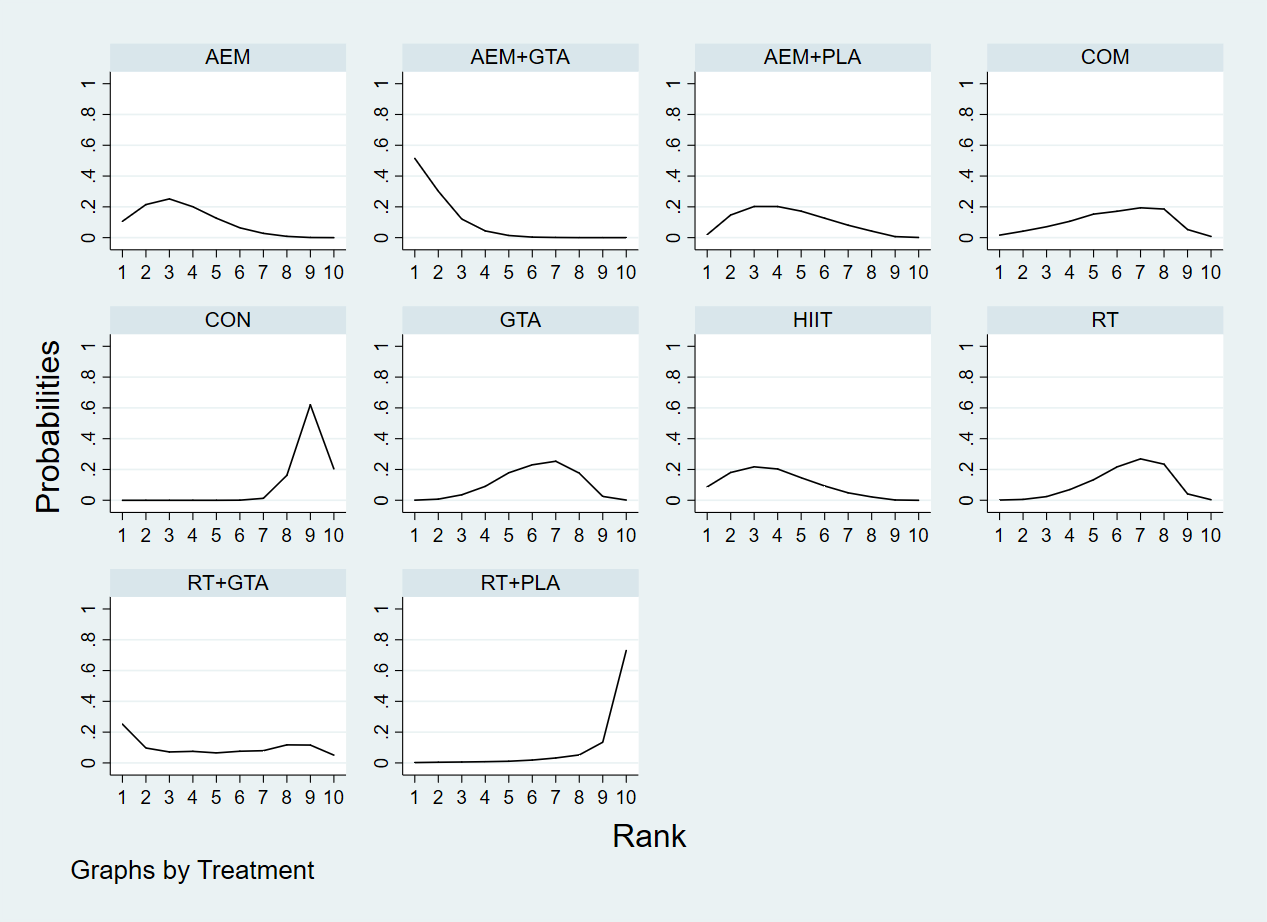

**Supplementary Figure 28. SUCRA figure for BF%.**

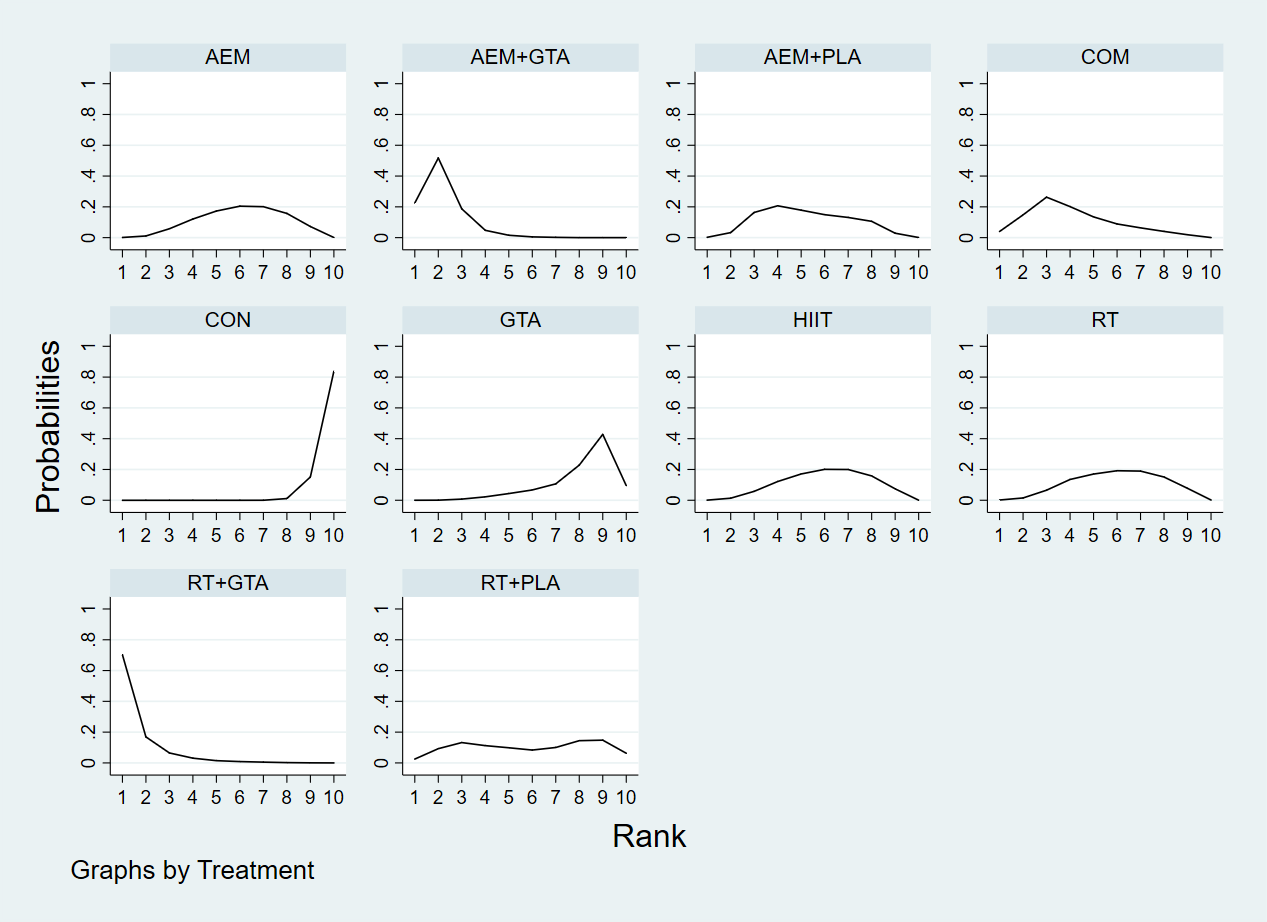

**Supplementary Figure 29. SUCRA figure for WC.**

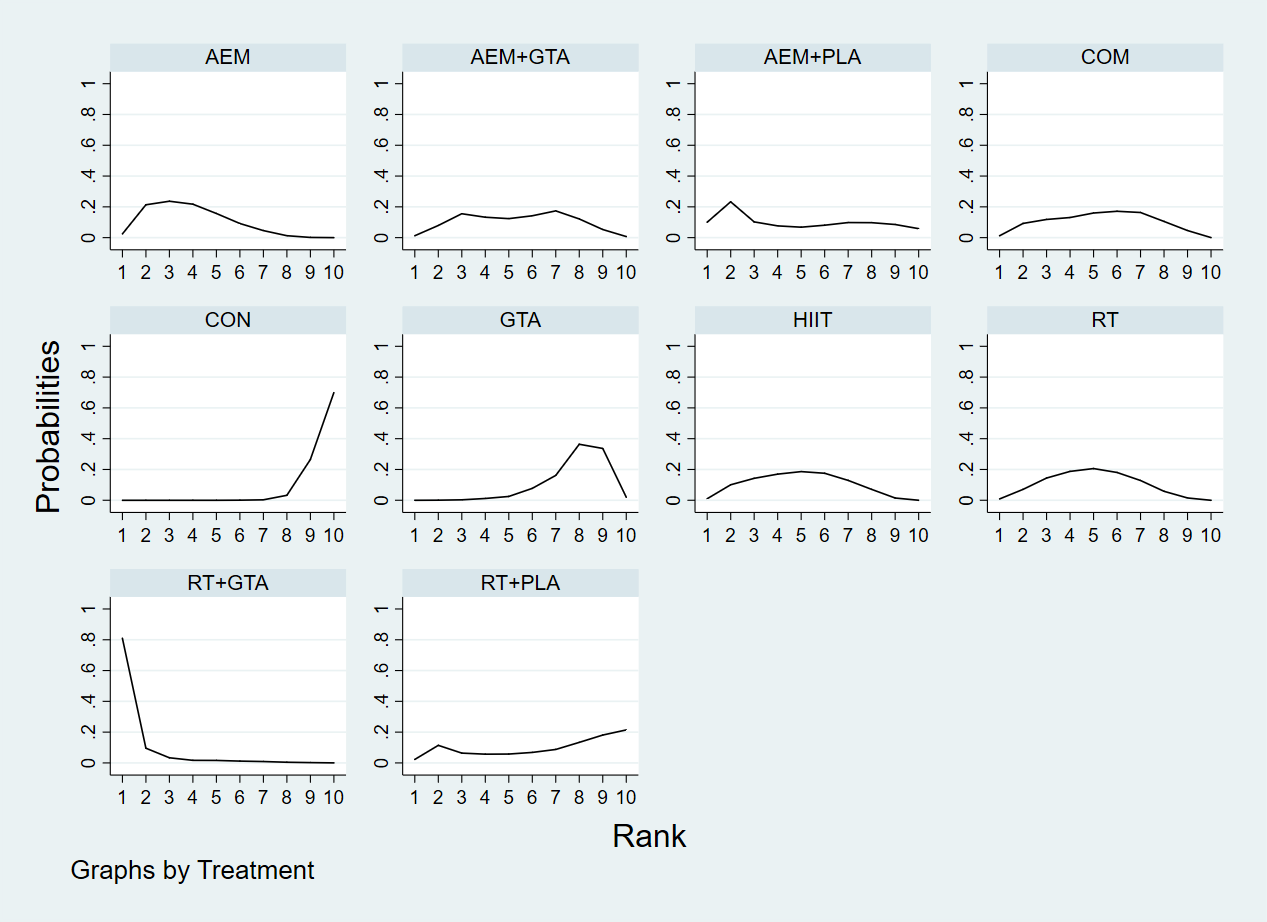

**Supplementary Figure 30. SUCRA figure for TG.**

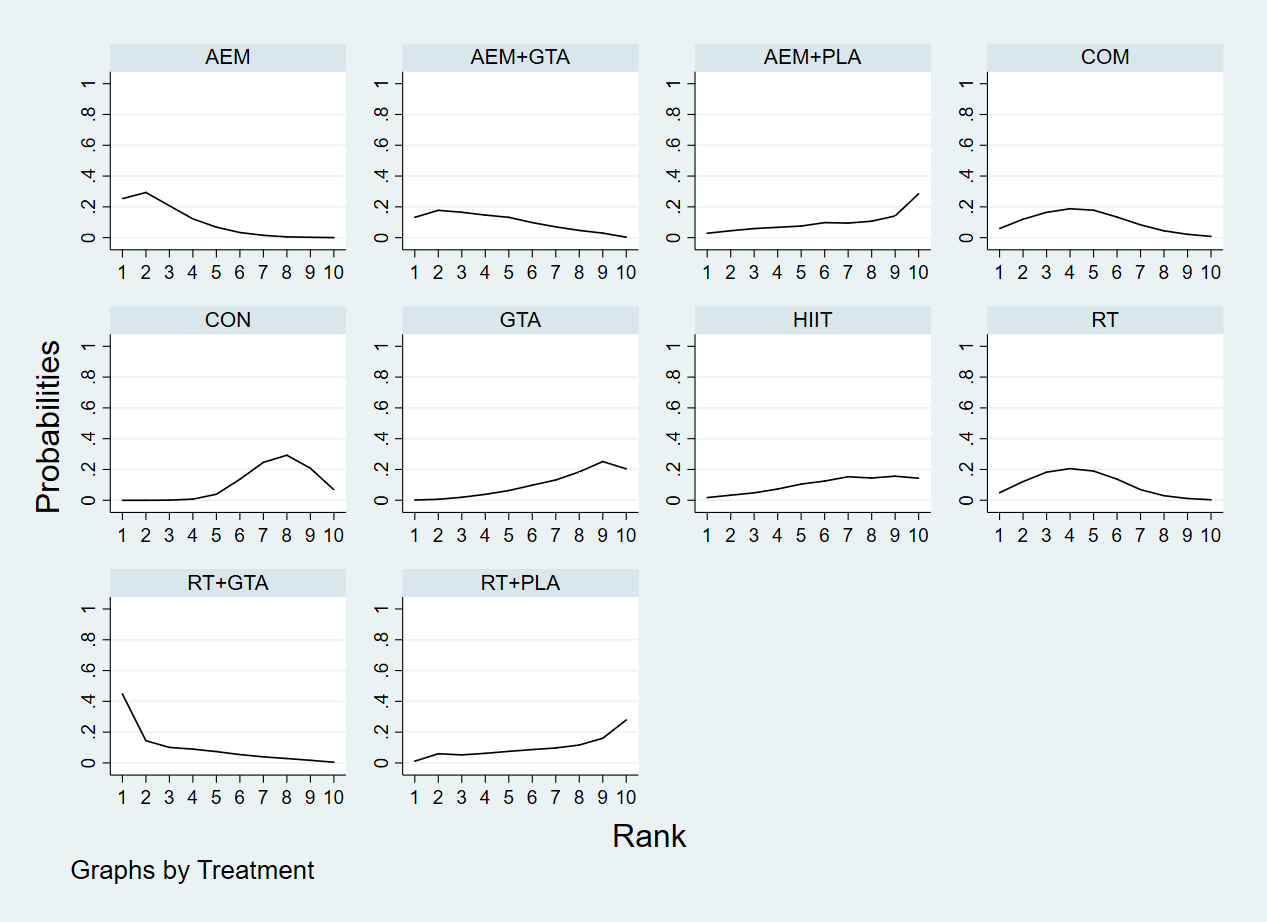

**Supplementary Figure 31. SUCRA figure for TC.**

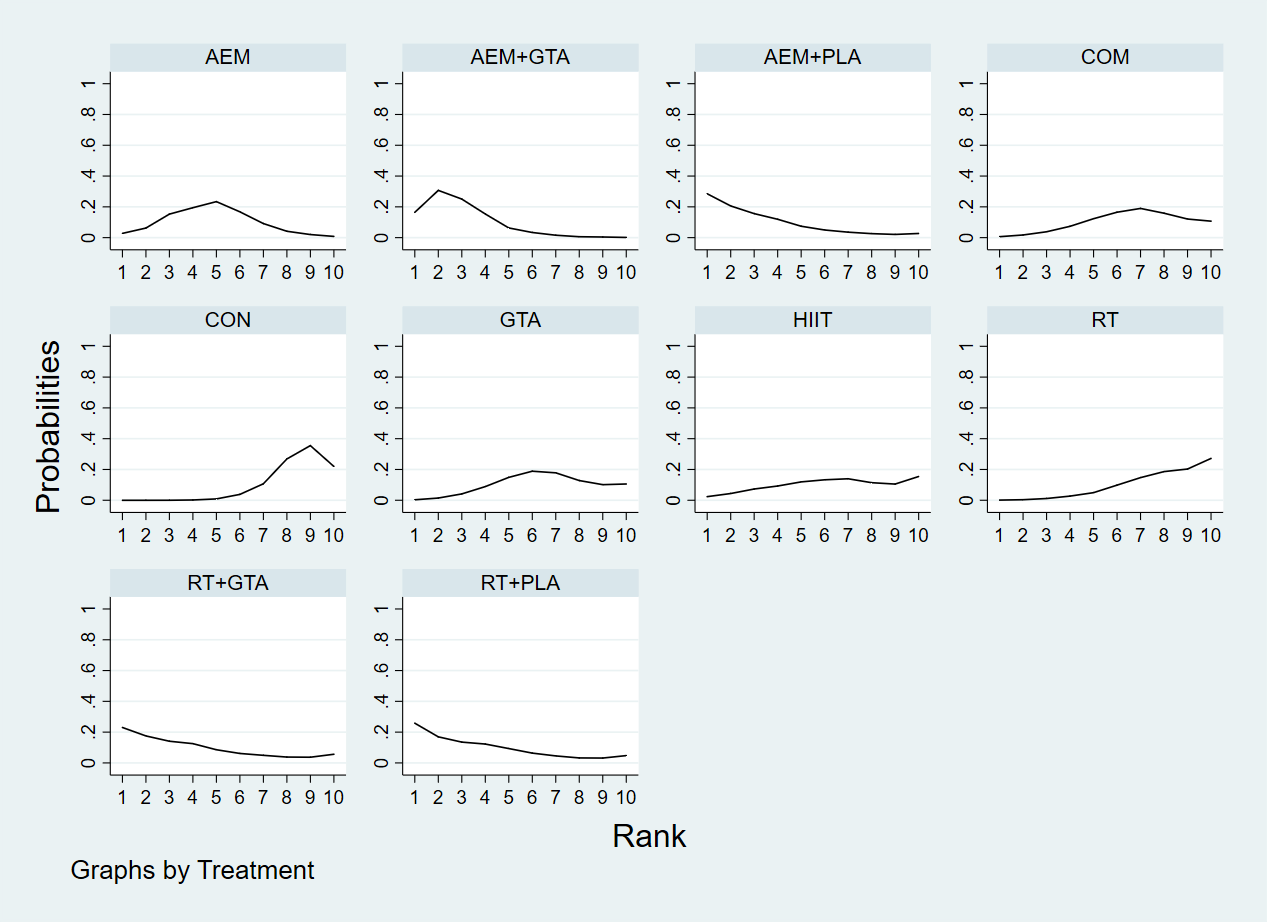

**Supplementary Figure 32. SUCRA figure for LDL.**

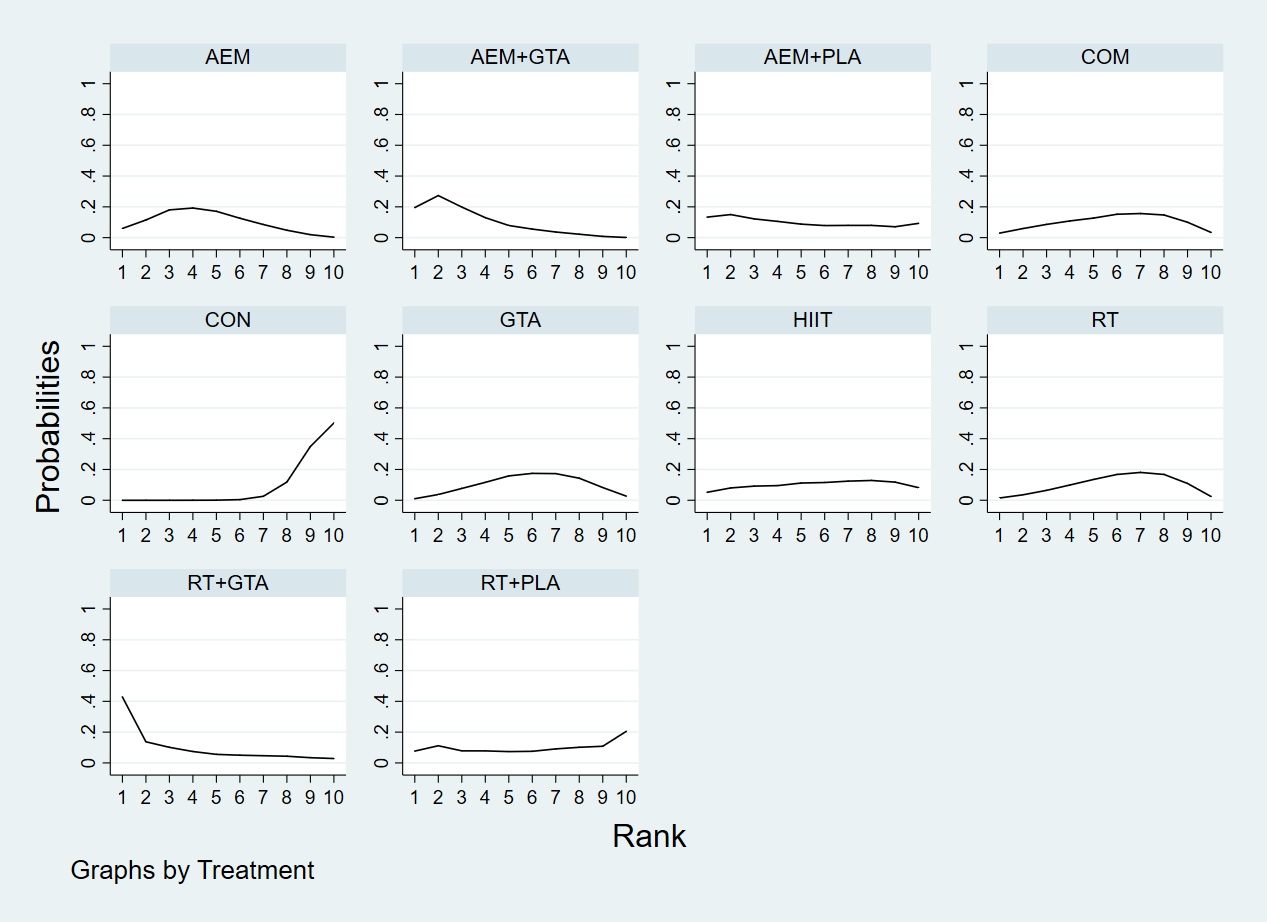

**Supplementary Figure 33. SUCRA figure for HDL.**

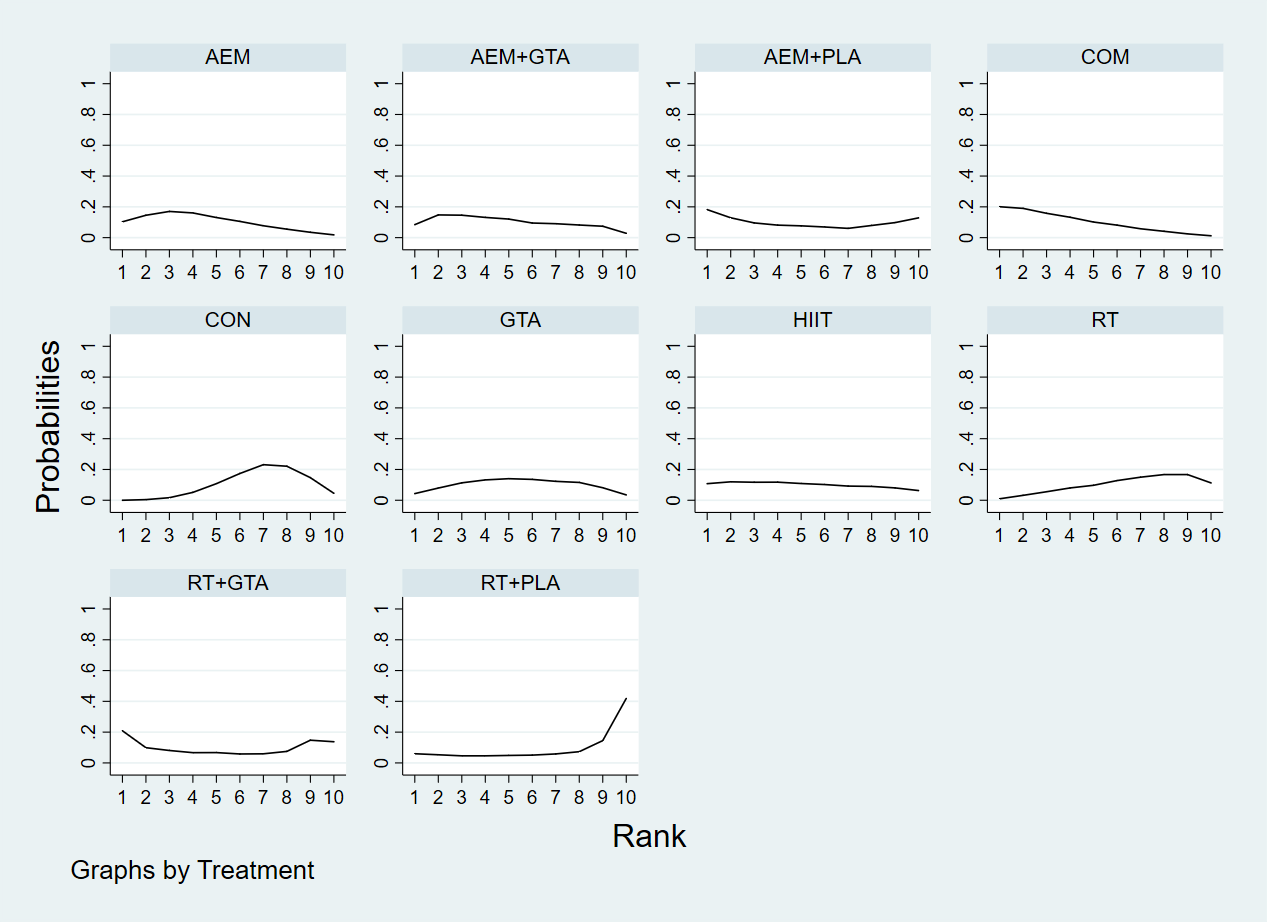

PRISMA CHECKLIST

| Section and Topic | Item # | Checklist item | Location where item is reported |
| --- | --- | --- | --- |
| Title |  |  |  |
| Title | 1 | Green Tea in Conjunction with Improved Physical Activity Is A Better Suggested Alternative Supplement Therapy for Obesity in Non-Aging Individuals—A Systematic Review and Network Meta-Analysis | Page 1 |
| abstract |  |  |  |
| Abstract | 2 | Increasing physical activity and drinking green tea are alternate non-surgical or pharmaceutical complementary treatments that many individuals opt to employ to manage obesity, and a number of recent clinical randomized controlled studies (RCTs) have shown interest in combining the two. The effectiveness of these three therapies in treating obesity has not been summarized, compared, or interpreted. A reticulated meta-analysis is superior than a meta-analysis for comparing and rating the effectiveness of different therapies for a particular condition. Through a reticulated meta-analysis, we wanted to examine the efficacy of three alternative supplementary treatments for obesity, namely green tea, physical activity, and green tea coupled with physical activity, and to provide recommendations. A total of 44 RCTs assessing the effects of nine alternative supplementary treatments on body weight, BMI, body fat percentage, waist circumference, total cholesterol, triglycerides, low-density lipoprotein, and high-density lipoprotein were included. Green tea combined with aerobic exercise may be optimal for reducing body weight, BMI, TC, and LDL; green tea combined with resistance training may be optimal for reducing body fat percentage and waist circumference; aerobic exercise may be optimal for reducing TG; and resistance combined with aerobic exercise may be optimal for increasing HDL. Nevertheless, based on the NMA results for the aforementioned eight indicators, we believe that the combination of green tea and increased physical activity is preferable to green tea or increased physical activity alone for non-elderly adult populations seeking non-surgical or pharmacological treatment or management of obesity. | Page 1 |
| introduction |  |  |  |
| Rationale | 3 | The core of exercise as a prescription therapy for obesity is increasing physical activity, boosting calorie intake, and mobilizing more adipose tissue to participate in the functioning of the energy system in order to burn excess fat[8].  In addition, a globally popular beverage, tea, may be a potentially beneficial dietary supplement for the treatment of obesity. As an unfermented Camellia sinensis product, green tea is rich in catechins that have significant health benefits, particularly epigallocatechin gallate EGCG, the main component of catechins (catechins make up 30-42% of the dried tea leaves, and EGCG accounts for 60-65% of the catechin content and is therefore regarded as the substance with the most significant health role)[9]. Yang has established that green tea and its catechins control hunger and limit energy absorption in his study[10], Boschmann's research has demonstrated that green tea regulates fat metabolism and thermogenesis[11], and Chatree's research has demonstrated that green tea inhibits fat formation and accumulation[12]. Therefore, physicians and researchers are increasingly using green tea as an effective and easily available dietary supplement for the treatment of obesity, and this is becoming a trend[13,14].  The use of green tea or increased physical activity as a supplementary treatment for obesity has been acknowledged for a long time. Additionally, a portion of recent research has begun to focus on combining the two for the treatment of obesity, with multiple animal models demonstrating that exercise combined with green tea has a positive effect on obesity control in high-fat-fed rats and is more effective than exercise or green tea alone[15,16]. This seems to indicate a novel supplemental alternative medicine for the treatment of obesity. | 2 |
| Objectives | 4 | Numerous randomized controlled trials (RCTs) have investigated the efficacy of the three modalities described above (exercise, green tea, and green tea during exercise) as an alternative complementary treatment for obesity; however, the results of individual RCTs on these interventions for obesity are not uniform and there are differences in evidence-based meta-analyses, indicating that these interventions may have different effects on the treatment of obesity. To our knowledge, there is no research compared the efficacy and consequences of these therapies for obesity. We feel it is necessary to do a pooled analysis to assess and identify the best therapies using a net meta-analysis in order to give physicians and the obese population with recommendations. | 2 |
| Methods |  |  |  |
| Eligibility criteria | 5 | According to PICOS principles, the original research were evaluated and incorporated[17]. (P) individuals (>18 years) having a BMI more than or equal to 25 kg/m2, excluding secondary obesity or obesity in conjunction with other conditions; (I) treatments of increased exercise or green tea or increased exercise in conjunction with green tea; (C) controls were given a conventional diet, placebo or similar calorie food as controls, and no extra physical activity or health education beyond everyday routines. (O) Body weight (BM), body mass index (BMI), waist circumference (WC), body fat percentage (BF%), plasma total cholesterol (TC), triglycerides (TG), high-density lipoprotein levels (HDL), and low-density lipoprotein levels (LDL) were among the outcomes measured after the intervention; (S) randomised controlled trials were utilized (RCTs). In order to ensure the integrity of the search, we also excluded conference abstracts, correspondence, reviews of works, pre-experimental protocols, literature reviews, crossover trial designs, and literature with unpublished data (but not literature with author permission), but we did not restrict the language. | 3 |
| Information sources | 6 | We consequently checked five internet databases, including Web of Science, Pubmed, Cochrane, Sport Discus, and EMBASE. | 2 |
| Search strategy | 7 | The search term is roughly “green tea OR tea OR green tea extract OR EGCG OR epigallocatechin gallate OR catechins AND exercise OR physical activity OR exercise, physical OR isometric exercise OR aerobic exercise OR exercise training OR training OR resistance training AND obesity OR obese OR overweight”. Detailed search strategies can be found in the Supplementary Table 1. | 2 |
| Selection process | 8 | The two researchers used the program Zotero 5.0 (Corporation for Digital Scholarship, Vienna, VA, USA) to do a three-round screening of the whole body of literature. The procedure was the following: (1) Eliminate duplicate publications from various databases. (2) After reading the abstracts and titles, those that did not correspond to the sort of literature were discarded. (3) Searching for the complete text of the remaining literature and reading it, removing material once again, and then selecting the literature to be included[19]. | 4 |
| Data collection process | 9 | A word table including seven parts was used for data extraction: (1) author's name and year of publication; (2) gender characteristics of individuals (male and female, male only, female only) and sample size; (3) body mass index; (4) age; (5) interventions; and (7) sample size (experimental and control groups) (6) experiment length (7) Outcomes. | 4 |
| Data items | 10 | Study,year,n,trt,mean1,sd1,mean2,sd2 | 4 |
| Study risk of bias assessment | 11 | The Cochrane Handbook version 5.1.0 (Cochrane, London, UK) was used by two researchers to evaluate the quality of each chosen piece of literature[20]. The instrument featured seven potential risks: (1) generation of a random sequence, (2) concealment of treatment allocation, (3) participant and (4) staff blinding, (4) insufficient outcome data, (5) selective reporting, and (7) other types of bias. Based on the number of components for which high ROB was potentially present, trials were categorized into three degrees of ROB: high risk (five or more components), moderate risk (three or four components), and low risk (two or fewer components). | 4 |
| Effect measures | 12 | BW, BMI, BF%, WC, TG, TC, LDL, HDL | 2 |
| Synthesis methods | 13 | By definition, we retrieved the mean (Mean) and standard deviation (SD) of the number of individuals participated in each RCT before and after the intervention. As the post-intervention impact, the mean and standard deviation at baseline were deducted from the mean and standard deviation after the intervention[22]. In addition, the missing standard deviations in the report were extracted using the formula  According to the definition of outcome indicators, a negative mean indicates a beneficial impact of treating obesity for WB, BMI, WC, BF%, TG, TC, and LDL, while a positive mean indicates a negative effect for HDL. In addition, since BM (in kg), BF% (in %), WC (in cm), and BMI (in kg/m2) have consistent units of measurement, MD (mean difference) was used for computation, however SMD (standardized mean difference) was utilized for TG, TC, LDL, HDL, as their units are not statistical[24].  An essential assumption of the NMA is the consistency and transferability of interventions; thus, before calculating the NMA, we conducted consistency and transferability tests on the combined data. I2 was used to evaluate inconsistency between studies, with I2 > 50% indicating high heterogeneity between studies and I2 ＜50% indicating low heterogeneity between studies. P values were used to evaluate transferability between studies, with P > 0.05 indicating good transferability between studies and P 0.05 indicating poor transferability between studies[25]. A horizontal line parallel to the horizontal axis in a funnel plot indicates a decreased probability of publishing bias, and vice versa[26].  The 'network map' diagram in STATA is used to visualize the connection between all studies, where each dot represents an intervention and each line indicates a direct comparison between two locations, or an indirect comparison through transmission if no line exists[27].  The creation of SUCRA (surface under the cumulative ranking curve) plots and P-values ranging from 0 to 1 is an essential feature of the NMA. In a comparison of various treatments, a P-value closer to 0 indicates that an intervention is less successful, while a P-value closer to 1 indicates that the intervention is more effective[28].  On the basis of the NMA calculations, a league table (network meta-analysis matrix) was constructed between the various treatments, which gives a visual depiction of the differences between the two comparisons of the various interventions. The diagonal lines across the table represent the different interventions, with the best intervention in the upper left corner and the worst intervention in the lower right corner. Below the diagonal lines are the estimates from the pairwise meta-analysis, where a 95% confidence interval (95% CI) in the estimate that does not contain '0' indicates a statistically significant difference between the pairwise comparisons | 5 |
| Reporting bias assessment | 14 | Funnel plot | Supplementary table |
| Certainty assessment | 15 | NA | NA |
| Results |  |  |  |
| Study selection | 16 | The search approach described above yielded a total of 8899 original documents in the database, plus three more documents discovered by manual search. After deleting duplicates, 5734 documents remained; for this section of the literature, the abstract and title were read; after eliminating 5564 documents, 174 documents remained; for this section of the literature, the entire text was read; after eliminating 130 documents, 44 documents remained. | 5 |
| Study characteristics | 16 |  | Supplementary table |
| Risk of bias in studies | 17 | 18% (8 studies) of the studies were deemed high risk, 31% (14 studies) were deemed low risk, and some (51%) were deemed intermediate risk. We consider that despite the fact that moderate risk accounts for fifty percent of the total, it is acceptable since the difficulties of adopting a double-blind approach to participants for exercise as an intervention is intrinsic to it and not due to the experimental design. The Supplementary Table 3 will present the details. | Supplementary table |
| Results of individual studies | 18 |  | Supplementary table |
| Results of syntheses | 19 |  | 5-10 |
| Reporting biases | 20 | Visual inspection of the horizontal line in the middle of the funnel plot revealed no significant publication bias. The funnel plots of eight outcomes can find in the Supplementary Figure 10 to 17. | 10 |
| Certainty of evidence | 21 | Inconsistency tests and transferability tests | Supplementary table |
| Discussion |  |  |  |
| Discussion | 22 |  | Page 10-12 |
| OTHER INFORMATION |  |  |  |
| Registration and protocol | 23 | Pending review |  |
| Support | 24 | NA |  |
| Competing interests | 25 | NA |  |
| Availability of data, code and other materials | 26 |  | Supplementary table |
